# SARS-CoV-2 in schools: genome analysis shows that concurrent cases in the second and third wave were often unconnected

**DOI:** 10.1101/2022.01.26.22269824

**Authors:** Madlen Stange, Eva Wuerfel, Jelissa Katharina Peter, Helena Seth-Smith, Tim Roloff, Severin Gsponer, Alfredo Mari, Blanca Cabrera Gil, Aitana Lebrand, Fanny Wegner, Ulrich Heininger, Julia Bielicki, Sarah Tschudin Sutter, Tanja Stadler, Karoline Leuzinger, Hans H. Hirsch, Markus Ledergerber, Simon Fuchs, Adrian Egli

## Abstract

**Background:** The risk of SARS-CoV-2 (SCoV2) infection in schools and student households is typically assessed using classical epidemiology whereby transmission is based on time of symptom onset and contact tracing data. Using such methodologies may be imprecise regarding transmission events of different, simultaneous SCoV2 variants spreading with different rates and directions in a given population. We analysed with high resolution the transmission among different communities, social networks, and educational institutions and the extent of outbreaks using whole genome sequencing (WGS).

**Methods and Findings:** We combined WGS and contact tracing spanning two pandemic waves from October 2020 to May 2021 in the Canton of Basel-City, Switzerland and performed an in-depth analysis of 235 cases relating to 22 educational institutions. We describe the caseload in educational institutions and the public health measures taken and delineate the WGS-supported outbreak surveillance.

During the study period, 1,573 of 24,557 (6.4%) children and 410 of 3,726 (11%) staff members from educational institutions were reported SCoV2 positive. Thereof, WGS data from 83 children, 35 adult staff in 22 educational institutions and their 117 contacts (social network, families) was available and analysed. 353 contextual sequences from residents of the Canton of Basel-City sequenced through surveillance were identified to be related to cases in the educational institutions. In total, we identified 55 clusters and found that coinciding SCoV2-cases in individual educational institutions were mostly introduced from different sources such as social networks or the larger community. More transmission chains started in the community and were brought into the educational institutions than vice versa (31 vs. 13). Adolescents (12-19 years old) had the highest case prevalence over both waves compared to younger children or adults, especially for the emerging Alpha variant.

**Conclusions:** Introduction of SCoV2 into schools accounts for most events and reflects transmission closely related to social activity, whereby teenagers and young adults contribute to significant parallel activity. Combining WGS with contact tracing is pivotal to properly inform authorities about SCoV2 infection clusters and transmission directions in educational settings and the effectiveness of enacted public health measures. The gathered data showing more clusters to seed in the community than vice versa as well as few subsequent in-school transmissions indicate that the agilely employed health measures for educational institutions helped to prevent outbreaks among staff and children.

The clinical trial accession number is NCT04351503 (clinicaltrials.gov).

## Introduction

During the first wave of the SARS-CoV-2 (SCoV2) pandemic in 2020, schools and day-care centres for children were widely closed around the world as part of the extensive measures, in order to reduce community-spread of the pandemic virus. In the following months, data suggested that children were not the main drivers of the SCoV2 pandemic; although children do get infected and transmit the virus, they are less susceptible to infection than adults [1–3] and also at low risk of a severe clinical course [4]. On the other hand, negative side-effects of the global lockdowns were found to be particularly pronounced in children and adolescents. Children from socio-demographic less privileged families regularly suffered from educational and psychological constraints because of the school closures [5,6].

In Switzerland, as in many other countries worldwide, the experiences from the strict lockdown during the first wave of the pandemic led to a broad social agreement to avoid further school closures during the ongoing pandemic with all possible efforts [7–9]. However, whether SCoV2 would circulate uncontrolled in schools and childcare facilities was controversially discussed and difficult to address due to lack of experience with the new pandemic virus. Viewpoints changed over time with the emergence of new data, more transmissible SCoV2 lineages, and an increasingly vaccinated adult population.

The widespread school closures had hampered the ability to collect data on the transmission risk in these settings. Studies addressing the question of SCoV2 infection patterns in schools and childcare facilities most often inferred infection clusters and attack rates based on date of symptom onset or positive test result, and contact tracing. Those studies largely concluded low contribution of schools to the overall dynamics of the epidemic, evidenced by few child-to-child transmissions [10–13] and few child-to-household transmissions [14]. Transmission and outbreaks that failed to be contained were shown to be due to insufficiently executed mitigation measures [11]. Importantly, these studies were often conducted over a short time-frame of a few weeks and primarily relied on counting numbers of SCoV2 infected students and staff members in schools, assuming that concurrently occurring infections in schools equate to contagion within the school. The directionality of transmission was deducted via the chronology of onset of symptoms. However, this may be misleading, as cases may arise from transmissions outside school premises. The diversification of SCoV2 into lineages [15] can be discerned by the application of whole genome sequencing (WGS). WGS provides the highest typing resolution and even allows the separation of single viral genotypes (variants) within and between samples from infected individuals. Ultimately, this means that the above found child-to-child and child-to-household transmission events could be even less, if controlled for virus identity of the infected persons. Further, studies that focussed on outbreak analyses in educational settings using WGS did not consider the outbreaks in relation to infection events in the larger community context [16] and hence could not draw conclusions about whether genotypic variation in the observed outbreak was due to viral evolution or the simultaneous introduction of similar variants from the community. A profound knowledge about the routes and directions of transmission events between and within schools and communities would allow adaptation of measurement concepts to reduce transmission events.

In this study, we address the question, whether clusters of SCoV2 infections in children and adolescents within educational settings over two pandemic waves (October 2020 to May 2021) were acquired in schools or childcare facilities, or if these could rather be traced back to transmission events within the family, other social networks, or the broader community. We further investigated the prevalence of detected SCoV2 lineages across different age groups. Our work used a complementary approach combining detailed contact tracing data with WGS of viral isolates to reconstruct the routes of transmission at high resolution.

## Methods

### Study design

In this observational retrospective epidemiological study, we describe SCoV2 transmission dynamics over a period of eight months from October 2020 to May 2021 within educational institutions of the Canton of Basel-City, Switzerland. This time frame includes the second and third SCoV2 local epidemic waves in Switzerland. The first wave is not included in the study because schools were closed early during the first wave and testing capacities were limited to symptomatic people, which led to an incalculable underestimation of the total caseload among children [17,18].

All cases in schools and child care facilities were monitored by the Child and Adolescent Health Services of Basel-City. Here, case clusters with suspected in-school transmission and contacts were identified for WGS and subjected to a molecular epidemiological outbreak analysis. Potential clusters of cases in schools were defined as four or more cases among students and children and staff in the same educational group appearing within ten days between single cases. We excluded cases that did not fit the definition of potential clusters. For not previously sequenced samples via our surveillance program, the patients’ nasopharyngeal swabs or saliva samples were collected from the testing laboratories and sent to the University Hospital of Basel for WGS and lineage typing. Two datasets were used for the subsequent analyses. Firstly, WGS data of SCoV2 isolates from all inhabitants of the canton of Basel-City were combined with WGS and contact tracing data from case clusters to infer routes of transmission in educational institutions for the respective study period. Secondly, WGS data from all inhabitants of the canton of Basel-City with available age data were used to infer SCoV2 lineage-specific prevalence per age group, as well as the odds of certain SCoV2 lineages affecting younger age groups more than adults.

### Ethical statement

The study was approved by the local ethical committee (EKNZ number 2020-00769). All data was anonymized for the analysis of the transmission events.

### Description of the school landscape in the Canton of Basel-City

In the Canton of Basel-City, children start compulsory education at the age of four years with two years of kindergarten. This is followed by six years of primary school (children aged 6 to 12 years) and three years of secondary school level I (children aged 12 to 15 years). This compulsory education is followed by two to four years of secondary school level II (children aged 15 years and older), which ranges from vocational education to college education and prepares for tertiary education. In 2021, at the Canton of Basel-City primary schools and kindergarten count 12,814 students and 1,906 teachers; secondary schools level I count 4,343 students and 650 teachers; secondary schools level II count 7,400 students and 1,170 teachers (data provided by the Department of Education of Basel-City). From the age of 15 weeks, children can be supervised in childcare facilities. Some of them foster open concepts with open age structures (children aged from 3 months to 14 years), which allow for mixing across age groups. Childcare facilities are often attended by students from different primary schools and kindergartens.

### General case management

The diagnostic laboratories notify each SCoV2 negative or positive test result via the national, mandatory reporting system to the Department of Health Basel-City as the official place of residence. The Department of Health Basel-City established a contact tracing team at the beginning of the pandemic. The management of SCoV2 positive cases was adjusted over time according to case prevalence and circulation of new Variants of Concern (VOCs), such as the Alpha variant (B.1.1.7), Gamma (P.1), and Beta (B.1.351) at the time. Throughout the pandemic, all individuals tested positive for SCoV2 (index patient (IP)) were contacted by phone and were isolated for ten days. Isolation started from the day after the onset of first symptoms or the date of sampling in asymptomatic patients. All individuals tested positive by an antigen test were advised to confirm infection with a SCoV2 specific nucleic acid amplification test (NAT) assay. All close contact persons (CCP) were also sent into quarantine for ten days - beginning from the day of the last close contact to the IP. Close contact outside of school and childcare facilities was defined as contact with less than 1.5 metres distance for more than 15 minutes without sufficient protective measures (e.g. wearing of masks). CCP were advised to take a NAT assay during the ten-day quarantine. During the isolation or quarantine, personal and clinical information was registered in the cantonal COVID Care App-database. The health condition of all IPs and CCPs was monitored through a smartphone app where questionnaires were filled out regarding the current severity or absence of symptoms. Individuals without the app were followed-up by phone. All IPs and CCPs were contacted at least twice during their isolation or quarantine. From 22 January 2021 to 5 March 2021, during the emergence of the Alpha variant, the Department of Health Basel-City quarantined all individuals living in the same household as CCPs. Further, all second-degree contact persons (close contact persons of CCPs) were advised to take a SCoV2 specific NAT assay between the 5th and 10th day after the last close contact. Exemptions for CCP from the obligation to test and quarantine were granted to fully vaccinated individuals and individuals with a SCoV2 infection up to six months prior to the close contact (**Table S1**).

Within the Department of Health Basel-City, the team of “Child and Youth Health Service” recorded and managed all SCoV2 cases occurring in schools and childcare facilities, including infections of children, youth, and educational staff. The Child and Youth Health Service decided on additional measures in these facilities such as quarantines of entire school classes. During the study period, several parameters regarding prevention, detection, and management of SCoV2 cases were repeatedly adjusted according to the epidemiological situation. These included testing recommendations, management of detected cases (e.g., extent of isolation and contact quarantine), and protection concepts in schools and in public.

### Molecular surveillance via whole-genome sequencing

All laboratory-confirmed SCoV2 positive nasopharyngeal swabs or saliva samples tested at the University Hospital Basel (UHB) [17,18], and samples from external providers requested by the Department of Health Basel-City were subjected to WGS as previously described [19]. Briefly, we used the NextSeq500 platform (Illumina) and the Artic 3.0 protocol and sequences were quality controlled using our in-house developed pipeline COVGAP. We identified viral lineages based on PANGO nomenclature [15]. During the study period the UHB attempted to sequence 4,960 samples, of which 3,433 were of sufficient quality for further investigations. WGS data were shared via the Swiss Pathogen Surveillance Platform [20] (SPSP, www.spsp.ch) and subsequently deposited at GISAID (https://www.gisaid.org) and the European Nucleotide Archive (ENA, https://www.ebi.ac.uk/ena/browser/home) at EMBL-EBI. Regular data exports were generated from SPSP to the Swiss Federal Office of Public Health. Isolates that were not specifically sequenced for this analysis were used as community or contextual sequences. We could, in most instances, access meta information such as host sex, age, place of residence and, travel history for the whole-genome sequenced samples originating from UHB, whereas semi-public and public data retrieved from external sources such as GISAID or ENA lack this information. Filtering data from UHB and additional samples by other providers from ENA for those originating from the Canton of Basel-City residents only between 1 October 2020 to 31 May 2021 yielded 4,674 sequences that were subjected to analyses of transmission direction (**Table S2**; 3,433 from UHB, 1,240 from ETHZ, 1 from HUG). A subset of 1,693 sequences contained information on age and were subjected to the analysis of lineage-specific infections in children and adolescents (**Table S2**; 1,589 from UHB, 4 from ETHZ).

### Assessment of infection risk of children for specific lineages from cases of residents of the Canton of Basel-City

We calculated census-normalised ratios of sequenced and reported cases in children in order to infer whether children and youth in different education institutions were more at risk to be infected with certain viral lineages than adults. The ratios were then scaled to 100,000 inhabitants. With the available metadata for this particular analysis, we cannot know which educational institution the infected person visited, so we inferred the educational institution using age as a proxy. We equated zero-to three-year-olds to “childcare”, four-to five-year-olds to “kindergarten” (i.e., mandatory pre-school); six-to eleven-year-olds to “primary school”, 12-to 15-year-olds to “secondary school Level I”, and 16-to 19-year-olds to “secondary school Level II. People aged 20 or older were assigned as “adults”. Case numbers can be found in **Table 2**.

This analysis was done for three significant time frames which differed by major health measure in the schools as outlined in **Figure 1**. The first time frame from 1 October 2020 to 31 December 2020, or second pandemic wave before the occurrence of VOCs; the second time frame from 1 January 2021 to 28 February 2021, or second pandemic wave with the Alpha variant emerging; and the third time frame from 1 March 2021 to 31 May 2021, or the third pandemic wave with the Alpha variant dominating.

**Figure 1.**
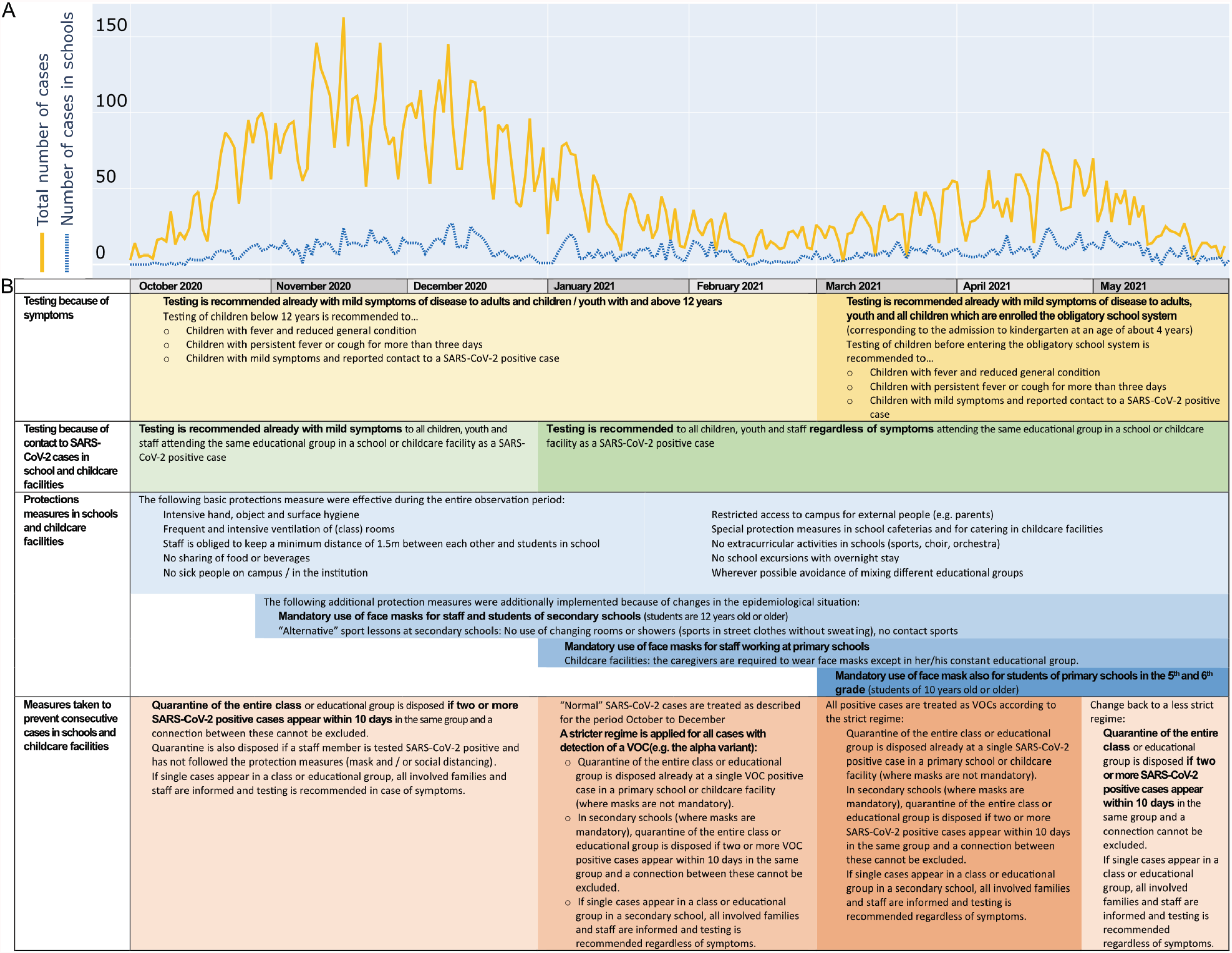
Case detection, prevention and protection concepts in schools in the Canton of Basel-City from 1 October 2020 to 31 May 2021. **(A)** Comparison of SCoV2 cases detected in the general population and in schools (including students and teachers) of the Canton of Basel-City. **(B)** Overview of testing policy, case management, and protection measures in schools and childcare facilities.

To quantify the risk of children in different institutions according to their age being infected with a certain lineage, we applying logistic regression using age groups as co-variate and extracted odds ratios and p-values for the exponential model coefficients. Confidence intervals are calculated as exponential 2.5% and 97.5% confidence intervals from the fitted model (point estimate of the coefficient +/-97.5% symmetric interval for a standard normal distribution x standard error of the coefficient).

### Detection of transmission chains in suspected outbreak events

Within the context of targeted resolution of suspected outbreaks in schools or childcare facilities, we analysed 235 high-quality SCoV2 genomes from 83 children and students (5.3% of 1,573 totally infected), 22 teachers and 13 educational caregivers (8.5% of totally infected staff), and 117 household contacts. These were from four *Kindertagestätten* (childcare facilities), two *Tagesstrukturen* (childcare facilities), one *Kindergarten* (i.e., pre-school), 13 primary schools, and two secondary schools level I. The percentage of included children and students is low as most cases did not fit our inclusion criteria (at least 5 clustered cases within 10 days) and due to a lower success rate in sequencing due to potential pre-analytical issues with samples from these cohorts. In contrast to the above-mentioned analysis, we know from contact-tracing, which educational institutions were attended by each IP and which persons were CCPs. We discerned the source of infection between (i) community-acquired (publicly-available contextual data on ENA (**Table S2**) from genomic surveillance from residents of the Canton of Basel-City), (ii) social-network-acquired from known contacts, or (iii) school-acquired. Transmission chains were defined as genomes that are connected by hierarchical clustering to a seed consisting of a 0 single nucleotide polymorphism (SNP) distance cluster. In detail, a three-step method was used to identify transmission chains. Firstly, sequences at 0 SNP distance (including missing regions) were grouped to obtain strict, mutually-exclusive, clusters, without applying a time window for cluster delimitation, as we were interested in the progression of transmission chains. At this point, if a SNP difference occurred in a missing region of an otherwise identical sequence, this was counted as a SNP difference of 1 and hence, the sequence with the missing region was placed in a separate cluster. To overcome this limitation and also to account for the fact that the virus can mutate within a transmission chain the initial 0 SNP clusters were expanded by adding sequences at 1 SNP distance from each member of the cluster in a second step. Finally, to avoid having too many clusters that contain several overlapping sequences, the expanded clusters were merged using hierarchical clustering based on the sequences belonging to each expanded cluster. The resulting merged superclusters allowed the identification of transmission chains based on nucleotide level mutations from the whole genome sequence. The code used for transmission chains is freely available (https://gitlab.sib.swiss/SPSP/sarscov2/cluster-detection).

### Tools used for data wrangling, statistical analysis, and visualisation of results

Data processing was done either in R version 4.0.3 [21] or Python v3.8.9 [22]. Odds ratios were calculated using questionr v0.7.4 [23]. The following packages were used for data wrangling and visualisation: tidyr v1.1.3 [24], ggplot2 v3.3.3 [25], pivottabler v1.5.2 [26], tibble v3.1.0 [27], hrbrthemes v0.8.0 [28], ggpubr v0.4.0 [29], pandas v1.2.4 [30], and plotly v4.14.3 [31].

## Results

### Overall case burden in the Canton of Basel-City between October 2020 and May 2021

Between 1 October 2020 and 31 May 2021, a total of 11,476 individuals (IP) with residency in the Canton of Basel-City tested positive for SARS-CoV-2, of which 693 had to be hospitalised and 150 died of COVID-19. The median age at diagnosis was 39 years (interquartile range [IQR] 25-55). Among all confirmed cases, 19% were aged ≤ 20 years, 68% were aged 20 - 65 years, and 13% were aged ≥ 65 years. Around 52% were female, 48% were male, 0.01% had no sex indicated. The 11,476 IPs in the observed time frame generated a total of 18,055 CCPs. The median age of CCPs was 22 years (IQR 9-44), of which 48% were aged ≤ 20 years, 42% were aged 20-65 years, and 10% were aged ≥ 65 years. 50.3% of CCP were female, 49.6% were male, 0.1% had no sex indicated.

### Prevention and protection concepts in schools

Figure 1. provides a detailed description of the adjustments of prevention and protection concepts during the study period. By the beginning of the study period in fall 2020, SCoV2 testing was well established, with sufficient testing capacities and easy access in testing centres and medical facilities throughout the city (**Figure 1A**). Initially, low-threshold testing was recommended to all people aged 12 years and older showing mild symptoms of respiratory disease. Testing of children younger than 12 years was recommended only in case of fever and reduced general condition, persistent fever or cough for more than three days, or mild symptoms after close contact with a confirmed SCoV2 infected patient. In schools and childcare facilities, general protection measurements included intensive hand, object, and surface hygiene and frequent ventilation of (class) rooms (**Figure 1B**). Staff in all educational facilities and students attending schools beyond the compulsory school system were required to keep 1.5 m distance to each other, as well as to younger students and children. Mandatory use of face masks was introduced for staff and students (aged 12 years and older) in secondary schools from the beginning of the second wave in November 2020.

From October to December 2020, group quarantines of entire classes or educational groups were only imposed if two or more SCoV2 positive cases appeared within ten days in the same group and a connection between the cases could not be excluded. When a single case appeared in a class or educational group, all involved families and staff were informed and testing was recommended in case of symptoms.

In January 2021, when the new Alpha variant of concern reached the Canton of Basel-City, contact tracing policies were intensified to slow down its spread. The mandatory use of face masks was extended to the staff of primary schools. As evidence pointed towards increased infections in children caused by the Alpha variant, face masks also became mandatory for students of 5th and 6th grade in primary schools (children of 10 years and older). Group quarantines were now directed at single cases of infections with verified Alpha variant in schools and childcare facilities in all groups of children or students which did not wear facemasks. The Alpha variant was detected through variant-specific NAT established as of mid-January 2021 [32]. In settings with mandatory use of face masks, test recommendations were given to all students and staff of the same educational group. By March 2021, the Alpha variant had become dominant in Switzerland and contact tracing in schools no longer distinguished between different viral lineages, but all cases were managed with the regime described above. In May 2021, the epidemiological situation shifted again with decreasing case numbers in Switzerland. By then, people at risk largely had access to vaccination [33]. In schools and childcare facilities, the policies in the Canton of Basel-City were switched back to a less rigid regime. Group quarantines were only directed with two consecutive cases in the same educational group, regardless of the mandatory use of face masks in the group. However, test recommendations were still given with a low threshold to all people of the same educational group in every single case in a school and childcare facility.

### Cases in schools and childcare facilities by cantonal contact tracing

During the study period, a total of 1,983 SCoV2 positive cases among children and staff were recorded in schools and childcare facilities. The total number of SCoV2 positive cases separated by education level is shown in **Table 1**. In childcare facilities for ages between zero and six years, we saw a predominance of positive cases among staff (61.2%), whereas only 38.8% of reported cases were in children. In higher educational levels with older children, we observed a higher percentage of cases among children and adolescents compared to staff members (**Table 1**). In the primary level (kindergarten and primary school), 4.9% (623/12,814) of students and 12.4% (236/1,906) of staff tested positive, in secondary school level I 6.9% (298/4,343) of students and 6.2% (40/650) of staff tested positive, in secondary school level II 7.9% (586/7,400) of students and 2.6% (30/1,170) of staff tested positive. Hence from primary level to secondary level II the proportion of positive tested individuals increased for students and decreased for staff.

**Table 1.**
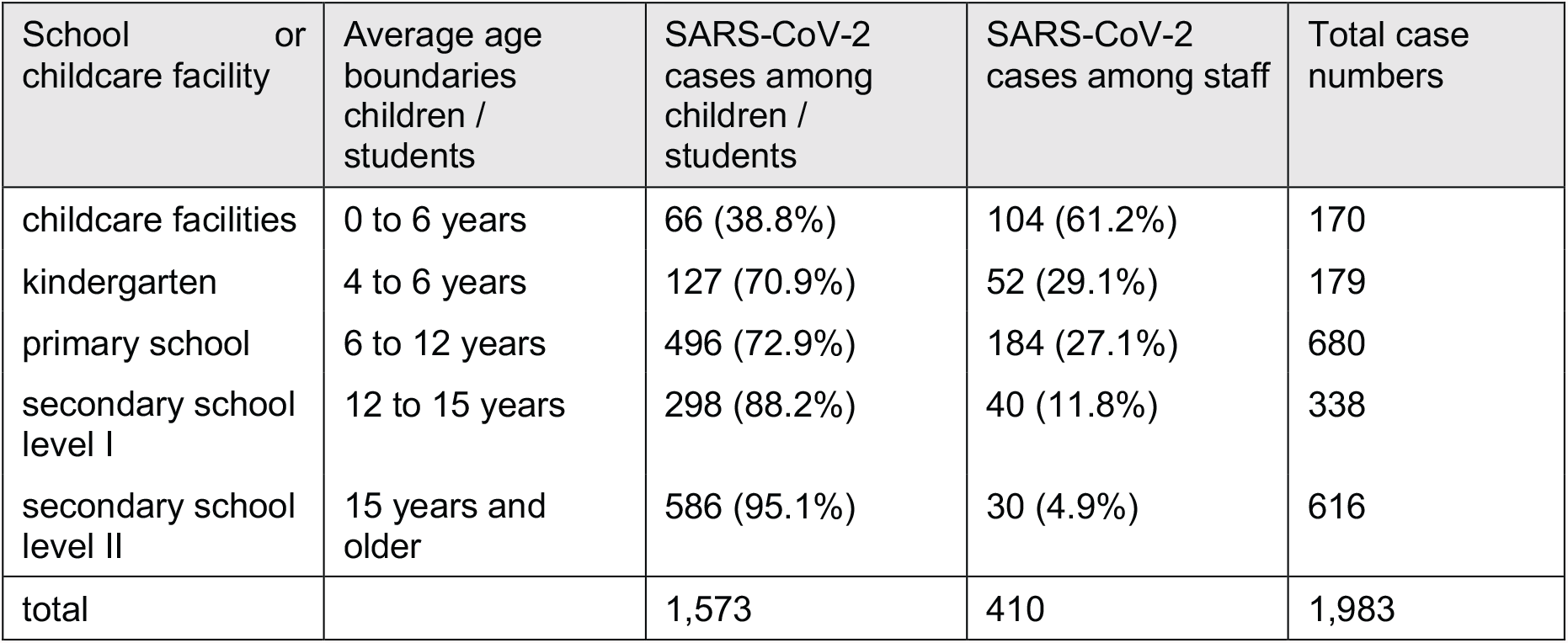
Number of SCoV2 positive cases during the study period from October 2020 to May 2021 in schools or childcare facilities, without contact persons, categorised by education level and resolved by children or students and staff registered by the Department of Health Basel-City. The percentage of children and students or staff in relation to the total number of cases in the educational level is given in brackets. Age boundaries for school and childcare facilities overlap as these were determined based on the actual knowledge of which facility was visited by each infected case.

The temporal change of relative proportions of teacher/student infections are shown in **Figure 2**. Interestingly, the distribution patterns changed around January 2021 with a higher case load predominantly in younger age groups covering childcare facilities, kindergarten and primary schools. In contrast, the distribution of cases among students versus staff in secondary school was consistent and relatively low during the entire study period.

**Figure 2.**
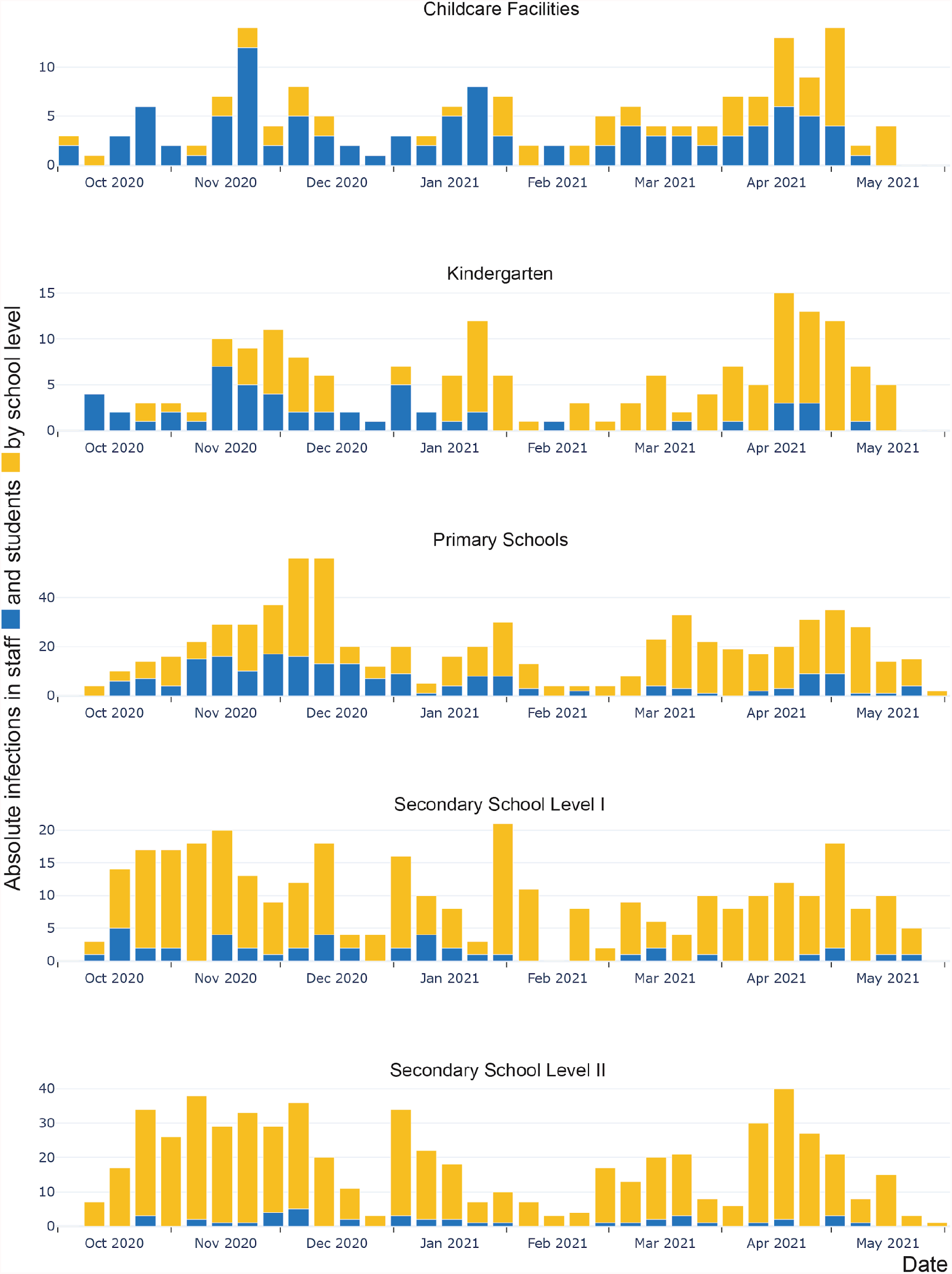
Temporal resolution in weeks of absolute positive SCoV2 cases in teachers and caregivers (staff; in blue) and students (yellow) resolved by school level during the study period (1 October 2020 to 31 May 2021).

### Contact tracing-informed molecular cluster identification in schools and childcare facilities

Based on the contact tracing information available to the Department of Health Basel-City, we sought data and samples from cases linked to 22 educational facilities, resulting in sequencing data for 83 infected students (primary to secondary level) and children (below primary level but in childcare), 35 staff, 117 household contacts, to compare to surveillance data for residents of the Canton of Basel-City. Based on this data, we attempted to define clusters of infections and to discern the source and direction of transmissions.

The cluster analysis identified another 353 contextual sequences from residents of the Canton of Basel-City that were putatively connected to the school cases. Across this dataset, we identified 55 clusters with a total of 566 cases and a size of two to 63 cases per cluster (**Figure 3**). The largest identified cluster (no. 37 in **Figure 3**) belongs to the Alpha variant and contains samples from 8 January 2021 to 10 March 2021 with 63 cases. This particular cluster consisted of seven students and two teachers, from two schools in the same city quarter, as well as seven contacts identified via contact tracing, and 47 additionally identified contextual sequences from the community. The earliest cases from January 2021 were contextual samples and only later, in early February 2021, cases were traced in the school environment. When focusing on individual schools (colour of symbols), we observed that temporally clustered cases per school frequently belong to different transmission chains, e.g., schools A, C, O, M and others (**Figure 3**). In addition, among the identified clusters, some contain cases from different educational institutions, e.g., clusters no. 0 and no. 2 (**Figure 3**), which were likely caused by cross-institutional visits such as primary school in the morning and a childcare facility in the afternoon.

**Fig. 3.**
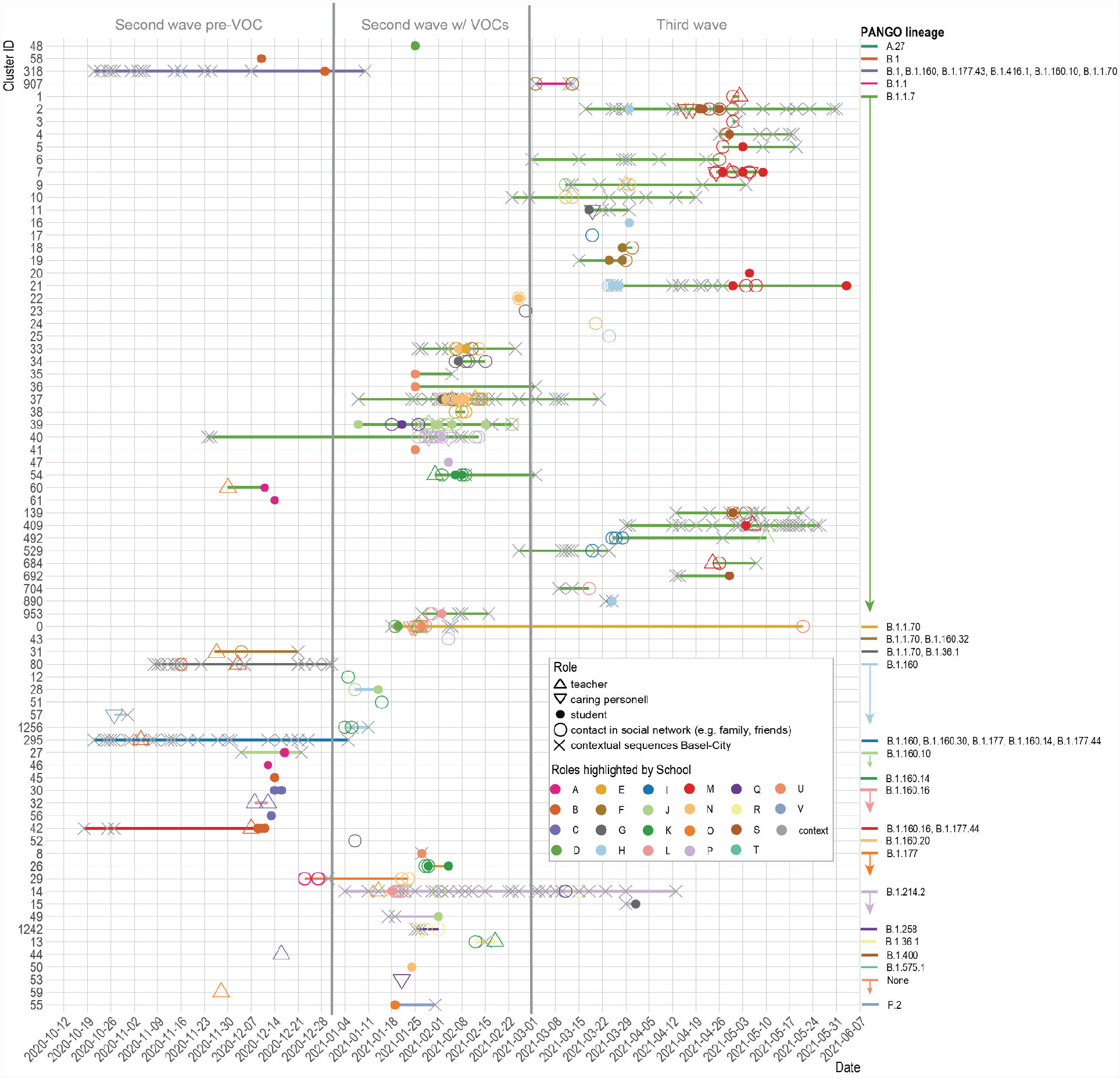
Temporal resolution of SCoV2 transmission chains in 55 identified clusters and 22 single cases including 83 students, 22 teachers,13 caregivers and 117 by contact tracing identified contacts from social networks (family, friends), plus another 353 sequences from residents of the Canton of Basel-City identified via the WGS-aided clustering approach. Each horizontal line represents clustered (genetically related) sequences coloured by PANGO lineage and vertically sorted by PANGO lineage in alphabetical order; symbols represent origin per detected case (school setting or contextual case from the Canton of Basel-City); colour of symbols represent origin per school.

In summary, taking the temporal succession of cases by sampling date in each cluster into account, the following plausible transmission scenarios unfold: we putatively identified 19 community-to-school transmissions (chains that start with “x”) without within-school transmissions and 12 with subsequent in-school transmissions (**Figure 3**); ten school-to-community transmissions without within-school transmission and three with within-school transmissions (5 of which showed the Alpha variant). In transmission clusters with in-school transmissions, we putatively identified seven with teacher-to-student, three with teacher-to-teacher, seven with student-to-student, and five with student-to-teacher transmissions (**Figure 3**).

Most outbreaks, in terms of numbers of involved cases, occurred during the second wave with the introduction of VOCs: here the Alpha variant had the largest impact. During the equally long two time periods before and after that (second wave pre-VOC and third wave), 17 and 28 clusters or single cases were counted, respectively (**Figure 3**).

We also identified 22 single cases (11 students, two teachers, one caregiver, eight contacts) from 28 November 2020 to 5 May 2021 that did not cluster with any other sequence registered in the Canton of Basel-City (**Figure 3**). By the study-inclusion criteria only clustered cases were subjected to contact tracing-informed molecular cluster identification. Hence the identified single cases were either connected to cases that failed sequencing or were true single cases. The lack of other related sequences in the community dataset indicates successful mitigation of broader transmission.

**Table 2.**
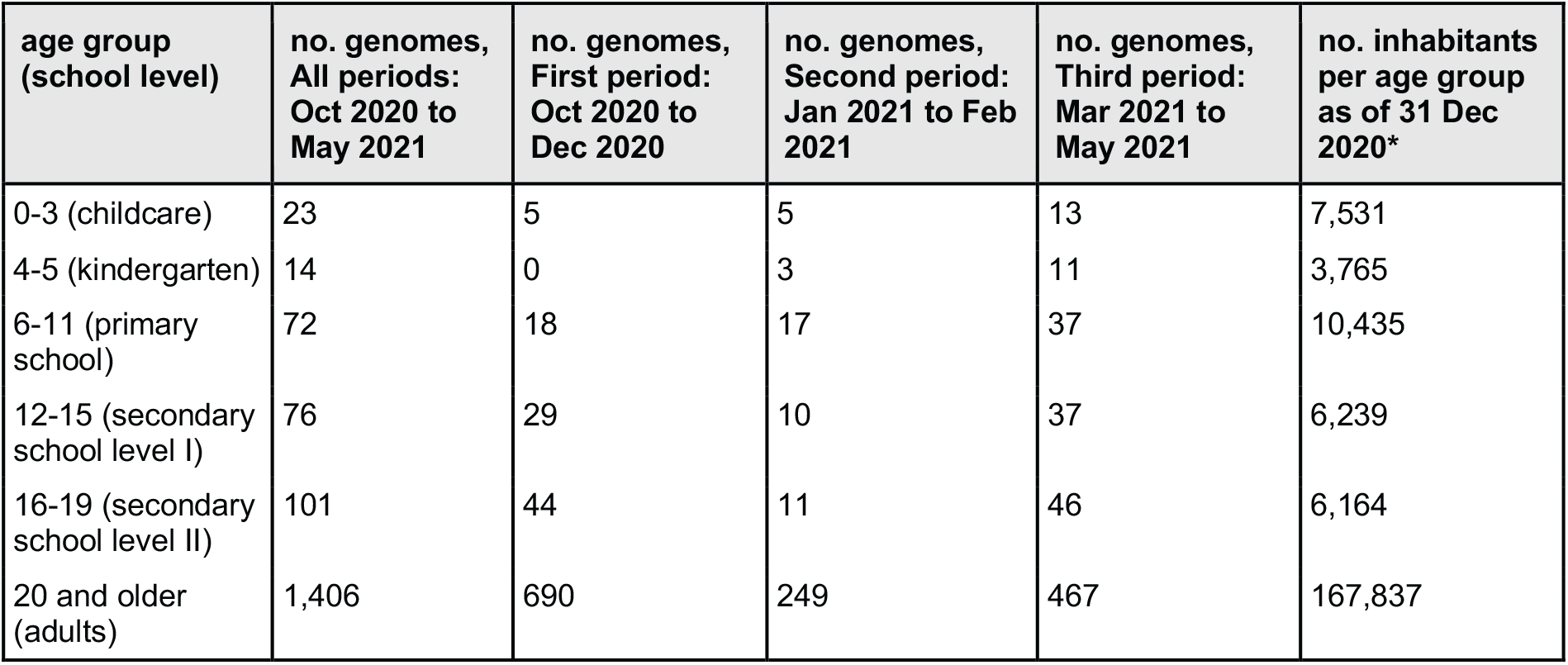
Number of available SCoV2 genomes from October 2020 to May 2021 and per subdivided time frame as per major health care measures, as well as census data for each age category from the Canton of Basel-City as of 31 December 2020 [34]* in age groups and their corresponding school level. School level was deduced solely based on age, as no information on school level per case was available.

### Temporal dynamics of lineage-specific infection risk of different age groups

Based on the SCoV2 PANGO lineage identity of detected cases in education institutions and infected residents of the Canton of Basel-City, we explored whether children of different age groups as compared to adults were of greater risk to be infected with certain SCoV2 lineages. In particular, we identified the most prevalent lineages and VOCs compared to other SCoV2 lineages circulating during the same time period. Generally, we observed an increasing number of infections with increasing age (**Table 2**), which could reflect a sampling bias. For this reason, we normalized the SCoV2 lineage per infection in each age category. This shows that SCoV2 lineage distribution in younger age groups is mirrored by the dynamic changes observed in adults (**Figure 4A-D, Figure S1**). Age- and lineage-specific prevalence demonstrates that during the second wave, before the occurrence of the Alpha variant, adjusted for their proportion of the population of the Canton of Basel-City, youths aged 16 to 19 years were more infected than adults (**Figure 4B**). During the third wave, driven by the Alpha variant, children aged 12 to 19 years were most affected (**Figure 4D**). Over both epidemic waves the Alpha variant had the highest prevalence among children of school age (aged 6 to 19 years) when compared to other age groups (**Figure 4A**). In terms of the odds to be infected with a certain lineage compared to adults, during the entire study period, children in the primary level and younger had 3 to 4 times the odds (p < 0.03) of an adult to be infected with the Alpha variant and only 0.3 times the odds (p = 0.027) to be infected with B.1.177 (**Table 3, Table S3**). During the first study period (second wave pre-VOCs, October to December 2020) children aged 0 to 3 years old had 9 times (p=0.02) the odds of an adult getting B.1.258 (**Table 3, Table S4**). Yet during this time the diversity of SCoV2 lineages (N = 57) and hence viral competition for hosts was large (**Table S8**). During the second study period (second wave with VOCs, January to February 2021) 31 SCoV2 lineages were detected in the Canton of Basel-City (**Table S9**). During that time children aged 6 to 11 years had 4 times (p=0.009) the odds of an adult to be infected with the Alpha variant (**Table 3, Table S5**). During the third study period (third wave, March to May 2021) 19 SCoV2 lineages were still circulating (**Table S10**) of which the Alpha variant was dominating the infection events. Accordingly, no differences in odds to be infected with a specific SCoV2 variant among age groups were found (**Table 3, Table S6**).

**Table 3.**
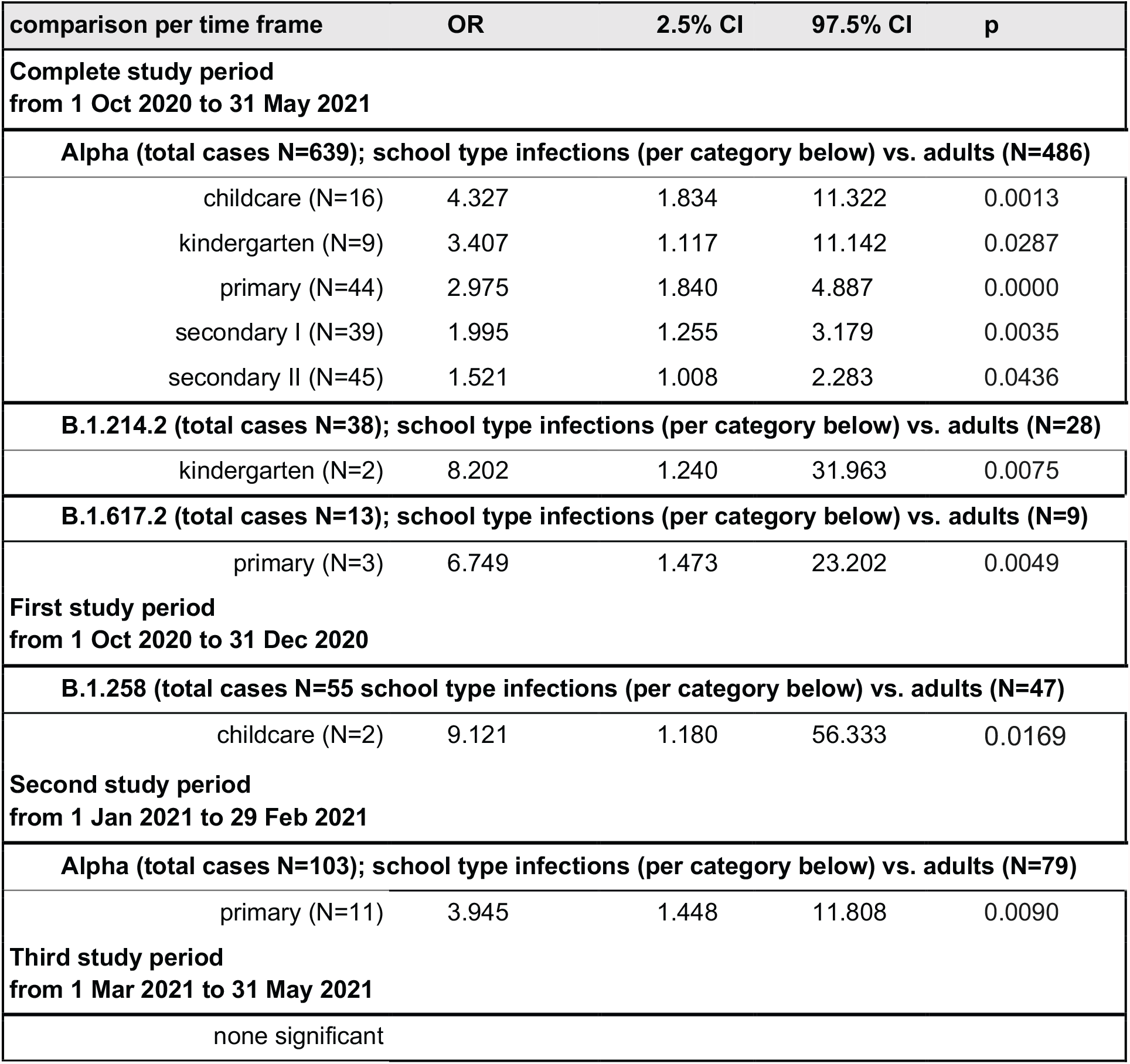
Summary table of odds ratios, with significant results and total case number per PANGO lineage larger than 10 only, in defined educational institutions (childcare [zero to three years old], kindergarten [four to five years old], primary school [six to eleven years old], secondary school level I [twelve to fifteen years old], secondary school level II [sixteen to nineteen years old]) compared to adults (older than 19 years) for prevalent lineages for the entire study period and selected time frames according to major health measures. Total case numbers used to calculate odds can be found in **Table 2**. All results to be found in **Tables S3-S6**. Number of cases per PANGO lineage in **Tables S7-S10**. “Secondary I” corresponds to Secondary school Level I, “Secondary II” to Secondary school Level II. OR: point estimate for odds ratio, CI: confidence interval, p: p-value, N: sample size.

**Figure 4.**
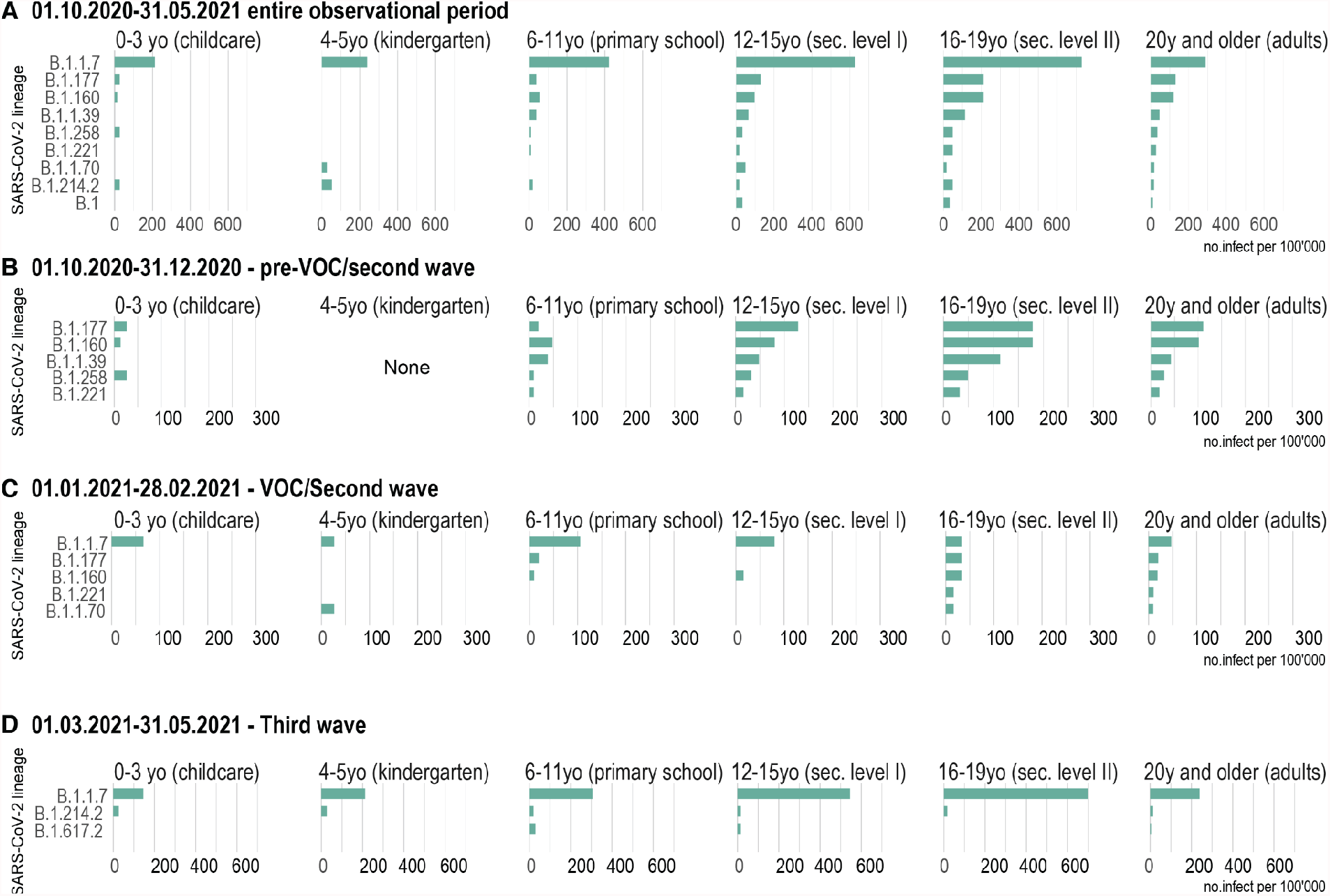
Population normalised SCoV2 lineage-specific prevalence per 100,000 inhabitants per age category (corresponding school levels shown in brackets) shown for the entire study period and per major time window as defined by major health measures in each period (for reference see **Figure 1**). Only the most prevalent lineages are shown here; the complete figures for the entire study period and the subperiods can be found in **Figures S1-S4**.

## Discussion

This study adds important evidence on the transmission dynamics of SCoV2 in schools and childcare facilities during two waves of the current COVID-19 pandemic, combining high-resolution WGS and contextual contact tracing data in a unique way.

The Canton of Basel-City sequenced 40.6% of its positive cases, which corresponds to a very high sequencing frequency per capita compared to other countries (https://www.covid19dataportal.org/statistics). In terms of total sequences deposited at GISAID per capita, Switzerland ranks 7th globally in a current estimation (Dec 2021). Hence, the evolution and distribution of viral lineages and variants (to the SNP-level) was extremely well monitored during the course of the epidemic, rendering a comprehensive picture of the virus transmission in the community as “background data” for the investigation of transmissions in schools and childcare facilities. Contact tracing of positive cases in schools and childcare facilities was operated centrally at the Child and Youth Health Services for the Canton of Basel-City, ensuring that all available information about such cases was collected and analysed continuously. This, to our knowledge, unique approach combining contact tracing and WGS data beyond the lineage identity from outbreaks in educational settings allowed us to analyse and interrelate to WGS data of other cases in the general community.

The observational investigation of transmission clusters in school settings showed that more transmission chains started in the community and were brought into the educational institutions than vice versa (31 vs. 13). Also, we observed fewer transmission chains resulting in in-school than outside-school transmissions (15 vs. 29), independent of the source. When in-school transmission occurred, staff commenced transmission chains to children more often than the other way around (7 vs. 5). These findings support previous studies relying on contact tracing data only, which also found a low rate of in-school transmissions, especially in younger children [10–12,35,36]. Our results add more granularity and the fact that the community largely seeded infection events within schools, which is important when evaluating protective measures. Staff-to-staff and student-to-student transmissions were 3 vs. 7, which is comparable to a study conducted in the UK where only about 10% of assumed transmission events were between students themselves [13]. This is remarkable taking into account that more than six times more students than teachers attend school, and given that especially young children interact closer to each other and are less conscious of hygienic measures. This observation may be biased by the higher test frequency and therefore higher detection rate in adults compared to children during the study period. The recommendation to test differed for children and adults, resulting in a higher threshold to test children especially below the age of 6 years old. However, the discrepancy might have been balanced out to a certain extent since children were symptomatic more often due to several other viruses spreading in childcare facilities and schools. Children therefore met test criteria frequently despite the high threshold imposed. Notably, wearing face masks was mandatory as of 5th grade for children, but not for children in the lower grades. We expected to observe shifts in the distribution of cases in childcare facilities and in the primary school level towards a higher proportion of affected students after January 2021. This shift is concomitant to the establishment of the Alpha variant in the Canton of Basel-City and the implementation of the mandatory use of face masks among staff in childcare facilities, kindergarten and primary school, as well as students of 5th and 6th grade in primary school. We hypothesize that the few transmissions among staff can be attributed to the implemented health measures, i.e., wearing of masks, distances, hygiene, and free testing even for mildly symptomatic individuals. Yet although transmission in schools were low, most lineage-specific SCoV2 prevalences were higher in children than in adults in both epidemic waves particularly in 12-to 19-year-olds, meaning adolescents had a higher relative case burden. Our analysis yielded an age-dependent infectiousness for the Alpha variant, with younger children being more likely to be infected than adults. Contrasting evidence is found in the current literature, from no differential affection of children and adults [37] to preliminary not yet peer-reviewed findings that support higher case burden the younger the children are [38].

Importantly, we were able to demonstrate that suspected clusters with temporally co-occurring cases were often caused by simultaneous introductions and sometimes transmissions of different SCoV2 variants. Our study is unique in the sense that WGS data allowed us to prove at highest resolution that many co-occurring cases were not connected. Previous studies on school outbreaks assumed viral transmission when several cases co-occurred in a classroom or childcare group within a certain time frame. In fact, we noted that during the second wave before the occurrence of VOCs, almost all apparent school clusters could be unravelled to stem from the random simultaneous appearance of different virus variants in the same classroom. These findings also influenced public health decisions and measures in the Canton of Basel-City, for instance challenging the assumptions previously leading to group quarantines.

Over the course of the SCoV2 epidemic, public health measures and contact tracing were adapted agilely, largely driven by the development of case numbers and the shift of the circulating dominant virus variants. However, apart from the initial lockdown, the Canton of Basel-City avoided closing schools and childcare facilities and did not implement strict online distance learning. Our data justify this management, showing that schools and childcare facilities were not high-risk venues driving virus transmission. The viral activity in educational institutions mirrored what was happening in local communities. Keeping schools and childcare facilities open was accompanied by a large number of protection measures, which were implemented in order to reduce the risk of virus transmissions. It is difficult to determine whether these measures had a significant influence in the dynamics of virus transmission, even after our detailed time-resolved analysis. However, we did observe that after the introduction of the mandatory use of face masks for teachers and caregivers in primary school, kindergarten and child care facilities and the increasing number of vaccinated adults, the ratio of affected adults compared to children was clearly reduced. This finding points towards a protective effect of mask wearing in educational institutions. This is concordant with other studies, showing that the use of face masks and the COVID-19 vaccine are one of the most efficient protection measures in the SCoV2 pandemic [39].

This study bears important limitations caused by missing data due to unsequenced cases. We found 22 single cases which had - derived from contact tracing - been related to clusters in schools or childcare facilities, for which we failed to detect other school or community cases despite our extensive SCoV2 database for the Canton of Basel-City. Some of these might indeed be “single cases” within the Canton of Basel-City and might have been imported from other cantons or countries. However, particularly after the establishment of VOCs, WGS more often failed because of pre-analytical quality or low viral load in the collected sample. This was probably due to changes in contact tracing policies, now advising also asymptomatic contact persons, as well as people with mild symptoms to undergo testing. Another important aspect is the sampling bias due to changing recommendations. We tried to sequence as many cases as possible to generate a comprehensive picture.

The SCoV2 pandemic is highly dynamic and therefore scientific results are in many ways short-lived. After completion of this study, the Delta variant (B.1.617.2) became the dominant virus variant as of July 2021 in the Canton of Basel-City. It caused the fourth epidemic wave for about three months, whereafter the first Omicron variant (B.1.1.529) cases were reported in our region in December 2021. During the fourth wave, we registered an increasing number of apparent infection clusters in schools and childcare facilities. It will be interesting to repeat the analyses depicted in this study including and comparing the wave caused by the more infectious Delta variant and the Omicron wave that is to come, especially since NAT pool-testing was established in the primary level as of June 2021.

## Conclusions

Analysing SCoV2 case clusters with WGS, this study shows that in the Canton of Basel-City from October 2020 to May 2021 cases of apparent transmission clusters in schools or childcare facilities were often not connected, but caused by different SCoV2 variants introduced from the community. In-school transmissions were infrequent and often involved adults. Our findings confirm that schools and childcare facilities were not hotspots for SCoV2 transmission during the Alpha wave and mostly reflected contagion in the community. During both waves, teenagers had higher caseloads than younger children or adults, which may be due to different behaviours. With the establishment of the Alpha variant as of January 2021, children were more likely to be infected than adults. Our data add additional arguments to the effectiveness of health measures, such as mandatory face masks and low testing thresholds.

## Data Availability

The SARS-CoV-2 genomes used in this study are accessible at the European Nucleotide Archive (ENA). Accession numbers are listed in Table S2.

## Acknowledgements

SARS-CoV-2 genome assemblies were performed at sciCORE (http://scicore.unibas.ch/) scientific computing centre at University of Basel, the support from the sciCORE team for the analysis is greatly appreciated. We also thank Daniel Walther from the development team of the Swiss Pathogen Surveillance Platform (www.spsp.ch), all authors who have shared their genomic data on ENA, and Gilles Dutilh from Clinical Trial Unit of the Department Clinical Research, University of Basel.

## Funding statement

The author(s) received no specific funding for this work.

## Ethics

The study was conducted according to good laboratory practice and in accordance with the Declaration of Helsinki and national and institutional standards. The study was assessed and approved by the local ethical committee (Ethikkommission Nordwest und Zentralschweiz, www.eknz.ch; ID number: 2020–00769). No signature was required according to the ethical assessment. All data from study participants (pediatric and adult patients) were reported in an anonymous fashion. The clinical trial accession number is NCT04351503 (clinicaltrials.gov).

## Notes

### Competing Interest Statement

The authors have declared no competing interest.

### Author Declarations

The study was assessed and approved by the local ethical committee (Ethikkommission Nordwest und Zentralschweiz, www.eknz.ch; ID number: 202000769). No signature was required according to the ethical assessment. All data from study participants (pediatric and adult patients) were reported in an anonymous fashion. The clinical trial accession number is NCT04351503 (clinicaltrials.gov)

