## Supplementary material for "SARS-CoV-2 in schools: genome analysis shows that concurrent cases in the second and third wave were often unconnected": Table S1

Prof. Adrian Egli, MD PhD

Division of Clinical Bacteriology and Mycology

University Hospital Basel

Petersgraben 4

4031 Basel, Switzerland

¶ These authors contributed equally to this work.

### Figures

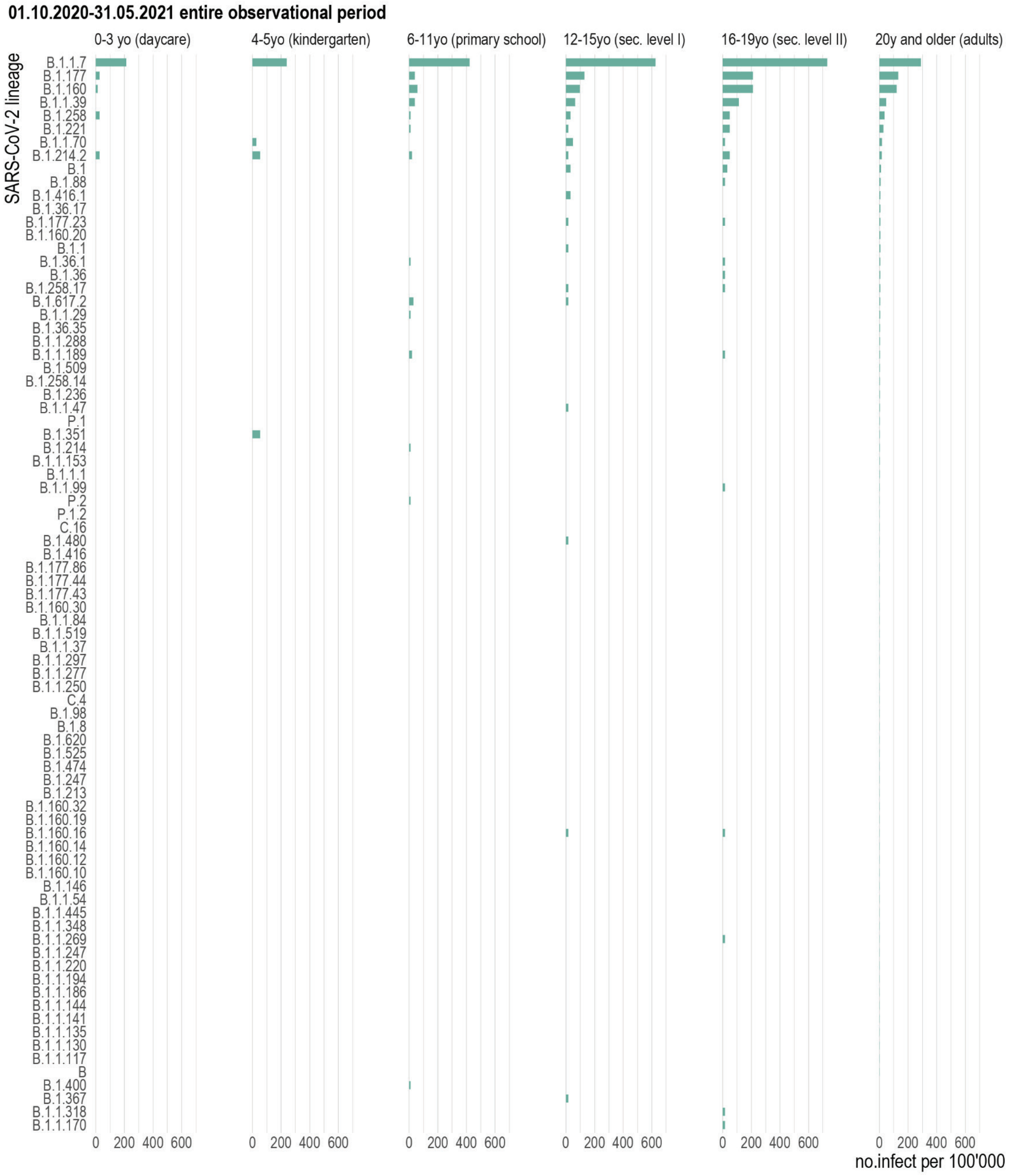


**Fig. S1**. Population normalised SARS-CoV-2 lineage-specific prevalence per 100’000 inhabitants per age category (corresponding school levels shown in brackets) shown for the entire observational period.

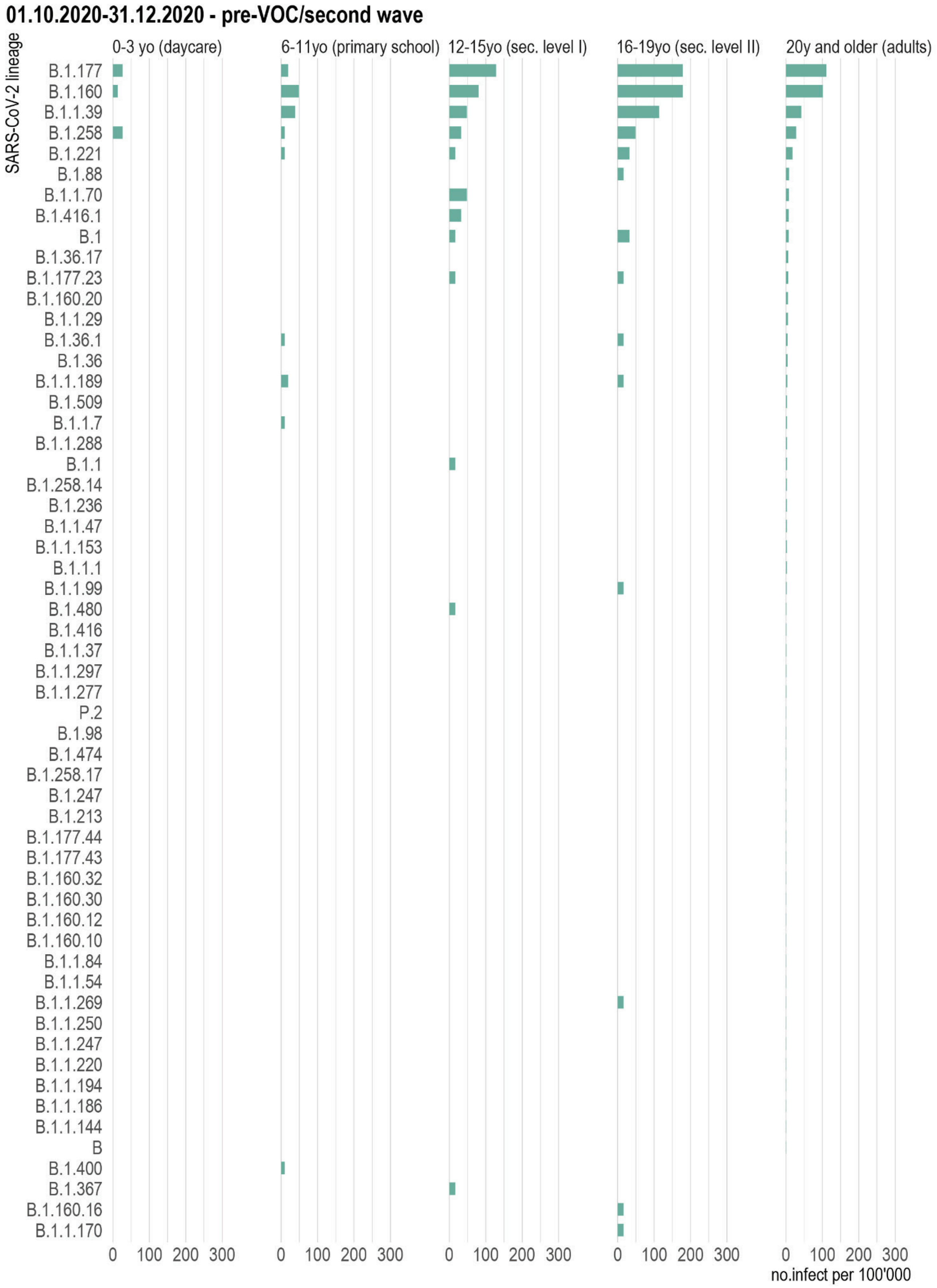


**Fig. S2**. Population normalised SARS-CoV-2 lineage-specific prevalence per 100'000 inhabitants per age category (corresponding school levels shown in brackets) in the canton of Basel-City, shown for the second wave before the establishment of the alpha variant, from 1 October 2020 until 31 December 2020. Note, that during this period no cases among children of 4 and 5 years (kindergarten age) were recorded.

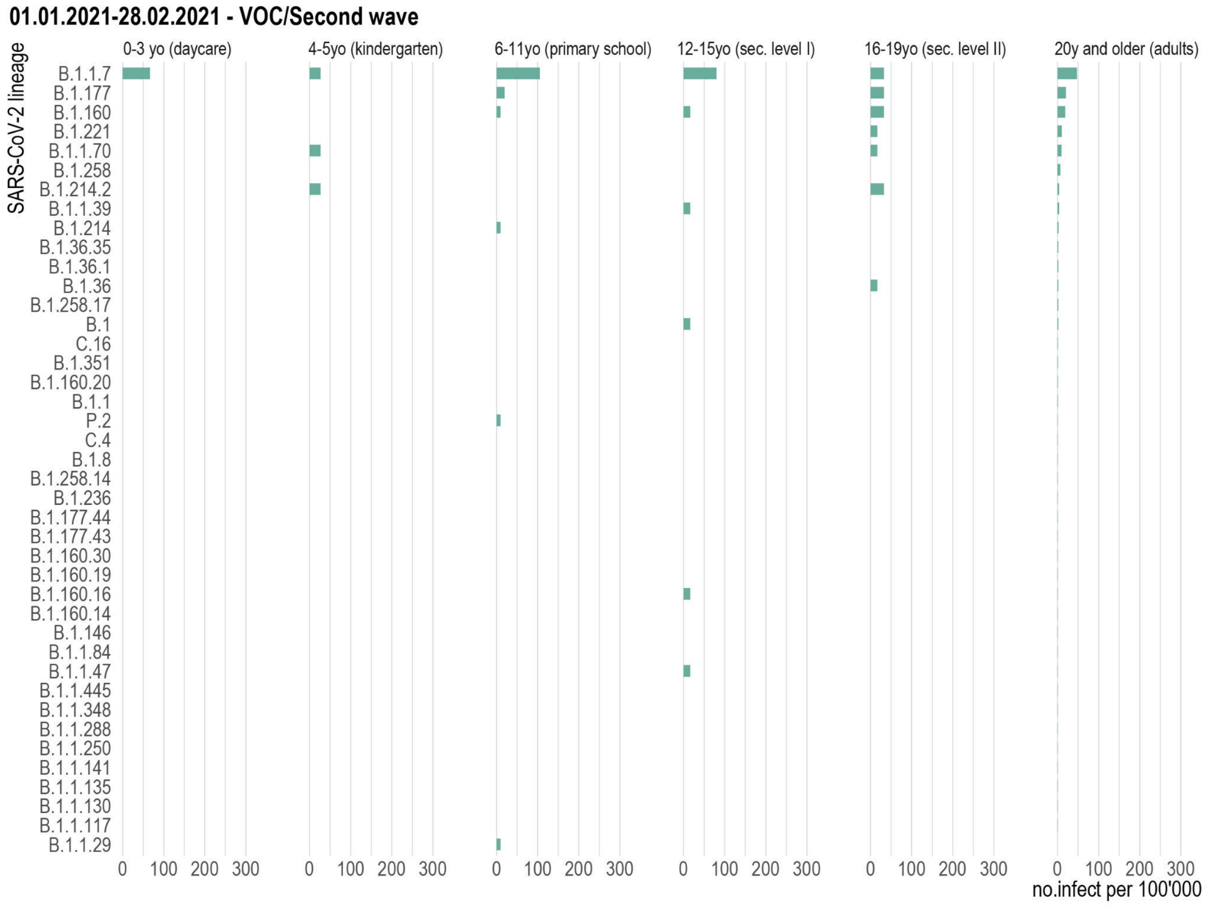


**Fig. S3**. Population normalised SARS-CoV-2 lineage-specific prevalence per 100’000 inhabitants per age category (corresponding school levels shown in brackets) in the canton of Basel-City, shown for the second wave with the establishment of the alpha variant, from 1 January 2021 to 28 February 2021.

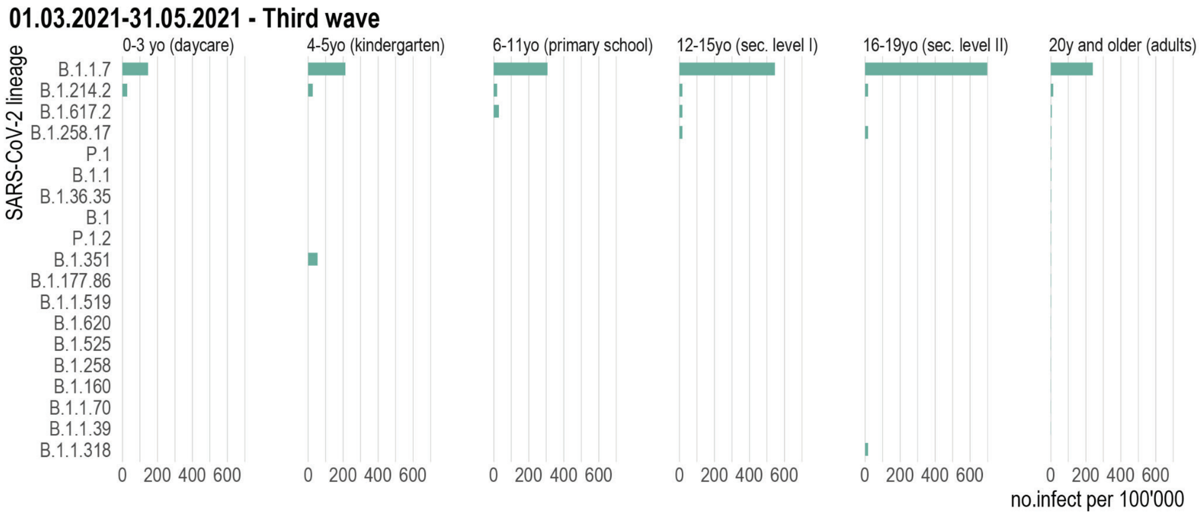


**Fig. S4**. Population normalised SARS-CoV-2 lineage-specific prevalence per 100’000 inhabitants per age category (corresponding school levels shown in brackets) in the canton of Basel-City, shown for the third wave, from 1 March 2021 to 31 May 2021.

### Tables

**Table S1.** Case definition, legitimacy and applied measures during study period.

| Case | Legitimacy | Measure |
|---|---|---|
| **Index Person (IP)** | Positive PCR test in symptomatic or asymptomatic individuals or positive antigen test in symptomatic individuals | 10 day isolation beginning from the day of first symptoms or the day of sampling in asymptomatic individuals |
| **Close Contact Person (CCP)** | Contact with less than 1.5 meters distance for more than 15 minutes without sufficient protective measures (e.g., wearing of masks) | 10 day quarantine beginning from the day of last close contact |
| **Household Member of CCP** | Individual living in the same household as CCP | From January 22 2021 to March 5 2021: 10 day quarantine with CCP |
| **Second Degree Close Contact Person** | Close contact person of CCP (Contact with less than 1.5 meters distance for more than 15 minutes without sufficient protective measures) | From January 22 2021 to March 5 2021: Recommendation to test (PCR) between the 5th and 10th day after close contact to CCP |
| **CCP not obliged for quarantine** | Fully vaccinated individuals and individuals with a SCoV2 infection up to six months prior to the close contact | None |

**Table S2**. GISAID identifiers for SARS-CoV-2 genomes from Basel-City residents with test dates between 01.10.2020 and 31.05.201. Identifiers in bold and italics with known age (not publicly disclosed) used to infer whether children in specific age classes are more at risk to be infected with specific SARS-CoV-2 lineages; normal and italics used for the analysis of transmission direction.

| | | |
|---|---|---|
| **EPI_ISL_1036070** | **EPI_ISL_1036127** | **EPI_ISL_1233691** |
| **EPI_ISL_1036130** | **EPI_ISL_830729** | **EPI_ISL_1233663** |
| **EPI_ISL_830730** | **EPI_ISL_830731** | **EPI_ISL_896081** |
| **EPI_ISL_2610950** | **EPI_ISL_1036084** | **EPI_ISL_896083** |
| **EPI_ISL_896080** | **EPI_ISL_830732** | **EPI_ISL_1036069** |
| **EPI_ISL_1036135** | **EPI_ISL_1233629** | **EPI_ISL_830734** |
| **EPI_ISL_1233628** | **EPI_ISL_861857** | **EPI_ISL_1036102** |
| **EPI_ISL_896108** | **EPI_ISL_1233692** | **EPI_ISL_896071** |
| | **EPI_ISL_830727** | **EPI_ISL_830733** |

| |
|---|
| **EPI_ISL_896115** |
| **EPI_ISL_2361631** |
| **EPI_ISL_1036116** |
| **EPI_ISL_1233630** |
| **EPI_ISL_896092** |
| **EPI_ISL_1233667** |
| **EPI_ISL_861858** |
| **EPI_ISL_2180925** |
| **EPI_ISL_861856** |
| **EPI_ISL_896097** |
| **EPI_ISL_896106** |
| **EPI_ISL_2420665** |
| **EPI_ISL_1036100** |
| **EPI_ISL_1036117** |
| **EPI_ISL_896077** |
| **EPI_ISL_930950** |
| **EPI_ISL_896079** |
| **EPI_ISL_896082** |
| **EPI_ISL_830735** |

| |
|---|
| **EPI_ISL_930878** |
| **EPI_ISL_1233664** |
| **EPI_ISL_896096** |
| **EPI_ISL_2361625** |
| **EPI_ISL_1233637** |
| **EPI_ISL_1014705** |
| **EPI_ISL_2420702** |
| **EPI_ISL_1036126** |
| **EPI_ISL_3344073** |
| **EPI_ISL_1036137** |
| **EPI_ISL_896073** |
| **EPI_ISL_930937** |
| **EPI_ISL_1014704** |
| *EPI_ISL_930976* |
| *EPI_ISL_2484550* |
| *EPI_ISL_2484562* |
| *EPI_ISL_2484231* |
| *EPI_ISL_2484372* |
| *EPI_ISL_2484461* |

| |
|---|
| *EPI_ISL_830823* |
| *EPI_ISL_830824* |
| *EPI_ISL_830825* |
| *EPI_ISL_830826* |
| *EPI_ISL_830827* |
| *EPI_ISL_830828* |
| *EPI_ISL_830829* |
| *EPI_ISL_830830* |
| *EPI_ISL_830831* |
| *EPI_ISL_830832* |
| *EPI_ISL_830833* |
| *EPI_ISL_830834* |
| *EPI_ISL_830835* |
| *EPI_ISL_830836* |
| *EPI_ISL_830837* |
| *EPI_ISL_830838* |
| *EPI_ISL_830839* |
| *EPI_ISL_830840* |
| *EPI_ISL_830841* |

| |
|---|
| *EPI_ISL_830842* |
| *EPI_ISL_830843* |
| *EPI_ISL_842649* |
| *EPI_ISL_830844* |
| *EPI_ISL_830845* |
| *EPI_ISL_830846* |
| *EPI_ISL_830847* |
| *EPI_ISL_830848* |
| *EPI_ISL_830849* |
| *EPI_ISL_830850* |
| *EPI_ISL_830851* |
| *EPI_ISL_830852* |
| *EPI_ISL_830853* |
| *EPI_ISL_830854* |
| *EPI_ISL_830855* |
| *EPI_ISL_830856* |
| *EPI_ISL_830857* |
| *EPI_ISL_830858* |
| *EPI_ISL_830859* |

| |
|---|
| *EPI_ISL_830860* |
| *EPI_ISL_830861* |
| *EPI_ISL_830862* |
| *EPI_ISL_830863* |
| *EPI_ISL_830864* |
| *EPI_ISL_830865* |
| *EPI_ISL_830866* |
| *EPI_ISL_830867* |
| *EPI_ISL_830868* |
| *EPI_ISL_830869* |
| *EPI_ISL_830984* |
| *EPI_ISL_830870* |
| *EPI_ISL_830871* |
| *EPI_ISL_830872* |
| *EPI_ISL_842650* |
| *EPI_ISL_830873* |
| *EPI_ISL_830874* |
| *EPI_ISL_830875* |
| *EPI_ISL_830876* |

| |
|---|
| *EPI_ISL_830877* |
| *EPI_ISL_830878* |
| *EPI_ISL_830879* |
| *EPI_ISL_830880* |
| *EPI_ISL_830881* |
| *EPI_ISL_830882* |
| *EPI_ISL_830883* |
| *EPI_ISL_830884* |
| *EPI_ISL_830885* |
| *EPI_ISL_830886* |
| *EPI_ISL_830990* |
| *EPI_ISL_830887* |
| *EPI_ISL_830888* |
| *EPI_ISL_830889* |
| *EPI_ISL_830890* |
| *EPI_ISL_830991* |
| *EPI_ISL_830891* |
| *EPI_ISL_830892* |
| *EPI_ISL_830893* |

| |
|---|
| EPI_ISL_830894 |
| EPI_ISL_830895 |
| EPI_ISL_830896 |
| EPI_ISL_830897 |
| EPI_ISL_830898 |
| EPI_ISL_830899 |
| EPI_ISL_830900 |
| EPI_ISL_830901 |
| EPI_ISL_830902 |
| EPI_ISL_830903 |
| EPI_ISL_830904 |
| EPI_ISL_830905 |
| EPI_ISL_830906 |
| EPI_ISL_830907 |
| EPI_ISL_830908 |
| EPI_ISL_830909 |
| EPI_ISL_830910 |
| EPI_ISL_830911 |
| EPI_ISL_830912 |

| |
|---|
| EPI_ISL_830913 |
| EPI_ISL_830914 |
| EPI_ISL_830915 |
| EPI_ISL_830916 |
| EPI_ISL_830992 |
| EPI_ISL_830993 |
| EPI_ISL_830777 |
| EPI_ISL_830917 |
| EPI_ISL_830918 |
| EPI_ISL_830919 |
| EPI_ISL_830920 |
| EPI_ISL_830921 |
| EPI_ISL_830922 |
| EPI_ISL_830923 |
| EPI_ISL_830778 |
| EPI_ISL_830924 |
| EPI_ISL_830994 |
| EPI_ISL_830925 |
| EPI_ISL_830926 |

| |
|---|
| EPI_ISL_830927 |
| EPI_ISL_830928 |
| EPI_ISL_830929 |
| EPI_ISL_830930 |
| EPI_ISL_830931 |
| EPI_ISL_830932 |
| EPI_ISL_830995 |
| EPI_ISL_830933 |
| EPI_ISL_830934 |
| EPI_ISL_830935 |
| EPI_ISL_830936 |
| EPI_ISL_830937 |
| EPI_ISL_830938 |
| EPI_ISL_830939 |
| EPI_ISL_830996 |
| EPI_ISL_830940 |
| EPI_ISL_830997 |
| EPI_ISL_830941 |
| EPI_ISL_830942 |

| |
|---|
| *EPI_ISL_830943* |
| *EPI_ISL_830944* |
| *EPI_ISL_830945* |
| *EPI_ISL_830946* |
| *EPI_ISL_830947* |
| *EPI_ISL_830998* |
| *EPI_ISL_830948* |
| *EPI_ISL_830949* |
| *EPI_ISL_830950* |
| *EPI_ISL_830951* |
| *EPI_ISL_830952* |
| *EPI_ISL_830953* |
| *EPI_ISL_830954* |
| *EPI_ISL_830955* |
| *EPI_ISL_830999* |
| *EPI_ISL_830956* |
| *EPI_ISL_830957* |
| *EPI_ISL_830958* |
| *EPI_ISL_830959* |

| |
|---|
| *EPI_ISL_830960* |
| *EPI_ISL_830961* |
| *EPI_ISL_831000* |
| *EPI_ISL_830962* |
| *EPI_ISL_830963* |
| *EPI_ISL_830964* |
| *EPI_ISL_830965* |
| *EPI_ISL_830966* |
| *EPI_ISL_830967* |
| *EPI_ISL_830968* |
| *EPI_ISL_830969* |
| *EPI_ISL_830970* |
| *EPI_ISL_830971* |
| *EPI_ISL_830972* |
| *EPI_ISL_830973* |
| *EPI_ISL_830974* |
| *EPI_ISL_830975* |
| *EPI_ISL_830976* |
| *EPI_ISL_830977* |

| |
|---|
| *EPI_ISL_930861* |
| *EPI_ISL_930864* |
| *EPI_ISL_930865* |
| *EPI_ISL_930868* |
| *EPI_ISL_930872* |
| *EPI_ISL_930873* |
| *EPI_ISL_930874* |
| *EPI_ISL_930875* |
| *EPI_ISL_930876* |
| *EPI_ISL_930877* |
| *EPI_ISL_930879* |
| *EPI_ISL_930880* |
| *EPI_ISL_930882* |
| *EPI_ISL_930883* |
| *EPI_ISL_930885* |
| *EPI_ISL_930886* |
| *EPI_ISL_930887* |
| *EPI_ISL_930888* |
| *EPI_ISL_930909* |

| |
|---|
| *EPI_ISL_930889* |
| *EPI_ISL_930890* |
| *EPI_ISL_930894* |
| *EPI_ISL_930895* |
| *EPI_ISL_930910* |
| *EPI_ISL_930896* |
| *EPI_ISL_930898* |
| *EPI_ISL_930911* |
| *EPI_ISL_930912* |
| *EPI_ISL_930913* |
| *EPI_ISL_930900* |
| *EPI_ISL_930914* |
| *EPI_ISL_930915* |
| *EPI_ISL_930917* |
| *EPI_ISL_930901* |
| *EPI_ISL_930902* |
| *EPI_ISL_930904* |
| *EPI_ISL_930918* |
| *EPI_ISL_930920* |

| |
|---|
| *EPI_ISL_930923* |
| *EPI_ISL_930924* |
| *EPI_ISL_930925* |
| *EPI_ISL_930926* |
| *EPI_ISL_930927* |
| *EPI_ISL_930928* |
| *EPI_ISL_930969* |
| *EPI_ISL_930929* |
| *EPI_ISL_930934* |
| *EPI_ISL_930935* |
| *EPI_ISL_930936* |
| *EPI_ISL_930970* |
| *EPI_ISL_930938* |
| *EPI_ISL_930939* |
| *EPI_ISL_930940* |
| *EPI_ISL_930971* |
| *EPI_ISL_930972* |
| *EPI_ISL_930973* |
| *EPI_ISL_930943* |

| |
|---|
| *EPI_ISL_930944* |
| *EPI_ISL_930945* |
| *EPI_ISL_930946* |
| *EPI_ISL_930947* |
| *EPI_ISL_930948* |
| *EPI_ISL_930949* |
| *EPI_ISL_930951* |
| *EPI_ISL_930952* |
| *EPI_ISL_930953* |
| *EPI_ISL_930955* |
| *EPI_ISL_930956* |
| *EPI_ISL_930957* |
| *EPI_ISL_930958* |
| *EPI_ISL_930959* |
| *EPI_ISL_931396* |
| *EPI_ISL_831001* |
| *EPI_ISL_1014691* |
| *EPI_ISL_930977* |
| *EPI_ISL_930978* |

| |
|---|
| *EPI_ISL_930979* |
| *EPI_ISL_930980* |
| *EPI_ISL_930983* |
| *EPI_ISL_930984* |
| *EPI_ISL_930985* |
| *EPI_ISL_930992* |
| *EPI_ISL_930993* |
| *EPI_ISL_930994* |
| *EPI_ISL_930995* |
| *EPI_ISL_930996* |
| *EPI_ISL_930997* |
| *EPI_ISL_930998* |
| *EPI_ISL_931001* |
| *EPI_ISL_931002* |
| *EPI_ISL_931003* |
| *EPI_ISL_931005* |
| *EPI_ISL_931006* |
| *EPI_ISL_931007* |
| *EPI_ISL_931008* |

| |
|---|
| *EPI_ISL_931009* |
| *EPI_ISL_931010* |
| *EPI_ISL_931011* |
| *EPI_ISL_931013* |
| *EPI_ISL_931014* |
| *EPI_ISL_931016* |
| *EPI_ISL_931019* |
| *EPI_ISL_931020* |
| *EPI_ISL_931021* |
| *EPI_ISL_931022* |
| *EPI_ISL_931023* |
| *EPI_ISL_931024* |
| *EPI_ISL_931026* |
| *EPI_ISL_931027* |
| *EPI_ISL_931029* |
| *EPI_ISL_931036* |
| *EPI_ISL_931030* |
| *EPI_ISL_931031* |
| *EPI_ISL_931038* |

| |
|---|
| *EPI_ISL_931040* |
| *EPI_ISL_931041* |
| *EPI_ISL_931042* |
| *EPI_ISL_930905* |
| *EPI_ISL_931044* |
| *EPI_ISL_931046* |
| *EPI_ISL_931047* |
| *EPI_ISL_931048* |
| *EPI_ISL_931049* |
| *EPI_ISL_931050* |
| *EPI_ISL_931052* |
| *EPI_ISL_931053* |
| *EPI_ISL_931054* |
| *EPI_ISL_931055* |
| *EPI_ISL_931056* |
| *EPI_ISL_931057* |
| *EPI_ISL_931058* |
| *EPI_ISL_931060* |
| *EPI_ISL_931062* |

| |
|---|
| *EPI_ISL_931063* |
| *EPI_ISL_931064* |
| *EPI_ISL_931065* |
| *EPI_ISL_931066* |
| *EPI_ISL_931068* |
| *EPI_ISL_931084* |
| *EPI_ISL_931070* |
| *EPI_ISL_931085* |
| *EPI_ISL_931086* |
| *EPI_ISL_931071* |
| *EPI_ISL_931087* |
| *EPI_ISL_931088* |
| *EPI_ISL_931398* |
| *EPI_ISL_931089* |
| *EPI_ISL_931090* |
| *EPI_ISL_931091* |
| *EPI_ISL_931399* |
| *EPI_ISL_931092* |
| *EPI_ISL_931093* |

| |
|---|
| *EPI_ISL_931094* |
| *EPI_ISL_931096* |
| *EPI_ISL_931097* |
| *EPI_ISL_931098* |
| *EPI_ISL_931099* |
| *EPI_ISL_931101* |
| *EPI_ISL_931102* |
| *EPI_ISL_931103* |
| *EPI_ISL_931104* |
| *EPI_ISL_931105* |
| *EPI_ISL_931106* |
| *EPI_ISL_931107* |
| *EPI_ISL_931108* |
| *EPI_ISL_830978* |
| *EPI_ISL_931111* |
| *EPI_ISL_931112* |
| *EPI_ISL_931116* |
| *EPI_ISL_931117* |
| *EPI_ISL_931118* |

| |
|---|
| *EPI_ISL_931120* |
| *EPI_ISL_931121* |
| *EPI_ISL_931122* |
| *EPI_ISL_931123* |
| *EPI_ISL_930906* |
| *EPI_ISL_931124* |
| *EPI_ISL_931128* |
| *EPI_ISL_931130* |
| *EPI_ISL_931132* |
| *EPI_ISL_931133* |
| *EPI_ISL_931134* |
| *EPI_ISL_931137* |
| *EPI_ISL_931138* |
| *EPI_ISL_931140* |
| *EPI_ISL_931141* |
| *EPI_ISL_931148* |
| *EPI_ISL_931149* |
| *EPI_ISL_931152* |
| *EPI_ISL_931153* |

| |
|---|
| EPI_ISL_931155 |
| EPI_ISL_931156 |
| EPI_ISL_931158 |
| EPI_ISL_931160 |
| EPI_ISL_931161 |
| EPI_ISL_931162 |
| EPI_ISL_931163 |
| EPI_ISL_931164 |
| EPI_ISL_931165 |
| EPI_ISL_931166 |
| EPI_ISL_931168 |
| EPI_ISL_931170 |
| EPI_ISL_931172 |
| EPI_ISL_931173 |
| EPI_ISL_931199 |
| EPI_ISL_931174 |
| EPI_ISL_931175 |
| EPI_ISL_931176 |
| EPI_ISL_931177 |

| |
|---|
| EPI_ISL_931179 |
| EPI_ISL_931180 |
| EPI_ISL_931181 |
| EPI_ISL_931182 |
| EPI_ISL_931183 |
| EPI_ISL_931200 |
| EPI_ISL_931184 |
| EPI_ISL_931185 |
| EPI_ISL_931186 |
| EPI_ISL_931187 |
| EPI_ISL_931201 |
| EPI_ISL_931189 |
| EPI_ISL_931190 |
| EPI_ISL_931202 |
| EPI_ISL_931191 |
| EPI_ISL_931192 |
| EPI_ISL_931193 |
| EPI_ISL_931203 |
| EPI_ISL_931194 |

| |
|---|
| EPI_ISL_931195 |
| EPI_ISL_931204 |
| EPI_ISL_931205 |
| EPI_ISL_931206 |
| EPI_ISL_931197 |
| EPI_ISL_931208 |
| EPI_ISL_931209 |
| EPI_ISL_931210 |
| EPI_ISL_931211 |
| EPI_ISL_931212 |
| EPI_ISL_931213 |
| EPI_ISL_931214 |
| EPI_ISL_931215 |
| EPI_ISL_931216 |
| EPI_ISL_931217 |
| EPI_ISL_931218 |
| EPI_ISL_931219 |
| EPI_ISL_931220 |
| EPI_ISL_931221 |

| | | |
|---|---|---|
| *EPI_ISL_931222* | *EPI_ISL_931251* | *EPI_ISL_931276* |
| *EPI_ISL_931223* | *EPI_ISL_931253* | *EPI_ISL_931279* |
| *EPI_ISL_931224* | *EPI_ISL_931254* | *EPI_ISL_931280* |
| *EPI_ISL_931225* | *EPI_ISL_931256* | *EPI_ISL_931281* |
| *EPI_ISL_931226* | *EPI_ISL_931257* | *EPI_ISL_931284* |
| *EPI_ISL_931227* | *EPI_ISL_931258* | *EPI_ISL_931285* |
| *EPI_ISL_931228* | *EPI_ISL_931259* | *EPI_ISL_931286* |
| *EPI_ISL_931229* | *EPI_ISL_931261* | *EPI_ISL_931287* |
| *EPI_ISL_931230* | *EPI_ISL_931262* | *EPI_ISL_931288* |
| *EPI_ISL_931231* | *EPI_ISL_931263* | *EPI_ISL_931289* |
| *EPI_ISL_931232* | *EPI_ISL_931264* | *EPI_ISL_931290* |
| *EPI_ISL_931233* | *EPI_ISL_931265* | *EPI_ISL_931291* |
| *EPI_ISL_931246* | *EPI_ISL_931267* | *EPI_ISL_931292* |
| *EPI_ISL_931234* | *EPI_ISL_931268* | *EPI_ISL_931293* |
| *EPI_ISL_931235* | *EPI_ISL_931269* | *EPI_ISL_931294* |
| *EPI_ISL_931236* | *EPI_ISL_931270* | *EPI_ISL_931295* |
| *EPI_ISL_931247* | *EPI_ISL_931272* | *EPI_ISL_931296* |
| *EPI_ISL_931248* | *EPI_ISL_931273* | *EPI_ISL_931297* |
| *EPI_ISL_931249* | *EPI_ISL_931274* | *EPI_ISL_931301* |

| |
|---|
| *EPI_ISL_931302* |
| *EPI_ISL_931303* |
| *EPI_ISL_931305* |
| *EPI_ISL_931306* |
| *EPI_ISL_931307* |
| *EPI_ISL_931309* |
| *EPI_ISL_931310* |
| *EPI_ISL_931312* |
| *EPI_ISL_931313* |
| *EPI_ISL_931376* |
| *EPI_ISL_931314* |
| *EPI_ISL_931400* |
| *EPI_ISL_931379* |
| *EPI_ISL_931315* |
| *EPI_ISL_931316* |
| *EPI_ISL_931381* |
| *EPI_ISL_931319* |
| *EPI_ISL_931320* |
| *EPI_ISL_931321* |

| |
|---|
| *EPI_ISL_931322* |
| *EPI_ISL_931324* |
| *EPI_ISL_931327* |
| *EPI_ISL_931328* |
| *EPI_ISL_931382* |
| *EPI_ISL_931329* |
| *EPI_ISL_931330* |
| *EPI_ISL_931331* |
| *EPI_ISL_931333* |
| *EPI_ISL_931335* |
| *EPI_ISL_931336* |
| *EPI_ISL_931383* |
| *EPI_ISL_931384* |
| *EPI_ISL_931338* |
| *EPI_ISL_931339* |
| *EPI_ISL_931386* |
| *EPI_ISL_931387* |
| *EPI_ISL_931388* |
| *EPI_ISL_931402* |

| |
|---|
| *EPI_ISL_931403* |
| *EPI_ISL_931404* |
| *EPI_ISL_931405* |
| *EPI_ISL_931406* |
| *EPI_ISL_931408* |
| *EPI_ISL_931409* |
| *EPI_ISL_931411* |
| *EPI_ISL_931412* |
| *EPI_ISL_931413* |
| *EPI_ISL_931414* |
| *EPI_ISL_931415* |
| *EPI_ISL_931416* |
| *EPI_ISL_931417* |
| *EPI_ISL_931418* |
| *EPI_ISL_931419* |
| *EPI_ISL_931423* |
| *EPI_ISL_931425* |
| *EPI_ISL_931426* |
| *EPI_ISL_931430* |

| |
|---|
| EPI_ISL_931431 |
| EPI_ISL_931432 |
| EPI_ISL_931435 |
| EPI_ISL_931444 |
| EPI_ISL_931446 |
| EPI_ISL_931447 |
| EPI_ISL_931072 |
| EPI_ISL_931073 |
| EPI_ISL_931075 |
| EPI_ISL_931076 |
| EPI_ISL_931077 |
| EPI_ISL_931078 |
| EPI_ISL_931342 |
| EPI_ISL_931081 |
| EPI_ISL_2610978 |
| EPI_ISL_2610979 |
| EPI_ISL_2610980 |
| EPI_ISL_2610982 |
| EPI_ISL_2610983 |

| |
|---|
| EPI_ISL_2610985 |
| EPI_ISL_2610986 |
| EPI_ISL_2610987 |
| EPI_ISL_2610989 |
| EPI_ISL_931343 |
| EPI_ISL_931344 |
| EPI_ISL_931345 |
| EPI_ISL_931346 |
| EPI_ISL_931347 |
| EPI_ISL_931348 |
| EPI_ISL_931350 |
| EPI_ISL_931351 |
| EPI_ISL_931352 |
| EPI_ISL_931353 |
| EPI_ISL_931354 |
| EPI_ISL_931355 |
| EPI_ISL_931356 |
| EPI_ISL_931357 |
| EPI_ISL_931358 |

| |
|---|
| EPI_ISL_931359 |
| EPI_ISL_1273465 |
| EPI_ISL_931361 |
| EPI_ISL_931362 |
| EPI_ISL_931389 |
| EPI_ISL_1273466 |
| EPI_ISL_931363 |
| EPI_ISL_1273469 |
| EPI_ISL_1273470 |
| EPI_ISL_1273471 |
| EPI_ISL_1273472 |
| EPI_ISL_1273675 |
| EPI_ISL_1273473 |
| EPI_ISL_1273474 |
| EPI_ISL_931364 |
| EPI_ISL_2611007 |
| EPI_ISL_2611009 |
| EPI_ISL_2611010 |
| EPI_ISL_2611011 |

| |
|---|
| EPI_ISL_1273464 |
| EPI_ISL_1273676 |
| EPI_ISL_2611014 |
| EPI_ISL_2611015 |
| EPI_ISL_2611016 |
| EPI_ISL_1273710 |
| EPI_ISL_1273480 |
| EPI_ISL_1273481 |
| EPI_ISL_1273482 |
| EPI_ISL_931366 |
| EPI_ISL_2367210 |
| EPI_ISL_2367211 |
| EPI_ISL_1273483 |
| EPI_ISL_2610946 |
| EPI_ISL_2610947 |
| EPI_ISL_2610948 |
| EPI_ISL_2610949 |
| EPI_ISL_931368 |
| EPI_ISL_931369 |

| |
|---|
| EPI_ISL_2610951 |
| EPI_ISL_1273485 |
| EPI_ISL_1273486 |
| EPI_ISL_1273740 |
| EPI_ISL_1273711 |
| EPI_ISL_1273487 |
| EPI_ISL_1273488 |
| EPI_ISL_1273489 |
| EPI_ISL_2367214 |
| EPI_ISL_2367215 |
| EPI_ISL_2367216 |
| EPI_ISL_1747647 |
| EPI_ISL_931448 |
| EPI_ISL_2408423 |
| EPI_ISL_1273741 |
| EPI_ISL_1273742 |
| EPI_ISL_1273491 |
| EPI_ISL_1273712 |
| EPI_ISL_1273492 |

| |
|---|
| EPI_ISL_1273493 |
| EPI_ISL_1273494 |
| EPI_ISL_1273708 |
| EPI_ISL_1296819 |
| EPI_ISL_1273495 |
| EPI_ISL_1273496 |
| EPI_ISL_1273497 |
| EPI_ISL_1273498 |
| EPI_ISL_1273499 |
| EPI_ISL_1273500 |
| EPI_ISL_1273501 |
| EPI_ISL_1273503 |
| EPI_ISL_1273504 |
| EPI_ISL_1273505 |
| EPI_ISL_1273506 |
| EPI_ISL_1273507 |
| EPI_ISL_1273508 |
| EPI_ISL_1273509 |
| EPI_ISL_1273510 |

| |
|---|
| *EPI_ISL_1273511* |
| *EPI_ISL_1273512* |
| *EPI_ISL_1273513* |
| *EPI_ISL_1273743* |
| *EPI_ISL_1273744* |
| *EPI_ISL_1273514* |
| *EPI_ISL_1273518* |
| *EPI_ISL_1273519* |
| *EPI_ISL_1273520* |
| *EPI_ISL_1273521* |
| *EPI_ISL_1273522* |
| *EPI_ISL_1273523* |
| *EPI_ISL_1273524* |
| *EPI_ISL_1273525* |
| *EPI_ISL_1273526* |
| *EPI_ISL_1273527* |
| *EPI_ISL_1273528* |
| *EPI_ISL_1273745* |
| *EPI_ISL_1273529* |

| |
|---|
| *EPI_ISL_1273746* |
| *EPI_ISL_1273530* |
| *EPI_ISL_1273531* |
| *EPI_ISL_1273532* |
| *EPI_ISL_1273533* |
| *EPI_ISL_1273534* |
| *EPI_ISL_1273535* |
| *EPI_ISL_1273536* |
| *EPI_ISL_1273537* |
| *EPI_ISL_1273538* |
| *EPI_ISL_1273539* |
| *EPI_ISL_2180924* |
| *EPI_ISL_1273540* |
| *EPI_ISL_1273541* |
| *EPI_ISL_1273542* |
| *EPI_ISL_1273543* |
| *EPI_ISL_1273544* |
| *EPI_ISL_1273545* |
| *EPI_ISL_1273546* |

| |
|---|
| *EPI_ISL_1273747* |
| *EPI_ISL_1273748* |
| *EPI_ISL_1273547* |
| *EPI_ISL_1273548* |
| *EPI_ISL_1273549* |
| *EPI_ISL_1273550* |
| *EPI_ISL_1273764* |
| *EPI_ISL_1273720* |
| *EPI_ISL_1273551* |
| *EPI_ISL_1273552* |
| *EPI_ISL_1273703* |
| *EPI_ISL_1273553* |
| *EPI_ISL_1273554* |
| *EPI_ISL_1296821* |
| *EPI_ISL_1273749* |
| *EPI_ISL_1273555* |
| *EPI_ISL_1273556* |
| *EPI_ISL_1273557* |
| *EPI_ISL_1273558* |

| |
|---|
| *EPI_ISL_1273559* |
| *EPI_ISL_2941526* |
| *EPI_ISL_2941527* |
| *EPI_ISL_3040150* |
| *EPI_ISL_2941528* |
| *EPI_ISL_2941531* |
| *EPI_ISL_3086259* |
| *EPI_ISL_3530748* |
| *EPI_ISL_3425783* |
| *EPI_ISL_3425784* |
| *EPI_ISL_3425785* |
| *EPI_ISL_3030091* |
| *EPI_ISL_3425787* |
| *EPI_ISL_3030092* |
| *EPI_ISL_3030093* |
| *EPI_ISL_3030094* |
| *EPI_ISL_3030095* |
| *EPI_ISL_3030096* |
| *EPI_ISL_3030097* |

| |
|---|
| *EPI_ISL_3030098* |
| *EPI_ISL_3030100* |
| *EPI_ISL_3030101* |
| *EPI_ISL_3086262* |
| *EPI_ISL_2367218* |
| *EPI_ISL_2367219* |
| *EPI_ISL_1273560* |
| *EPI_ISL_1273561* |
| *EPI_ISL_1273562* |
| *EPI_ISL_1273563* |
| *EPI_ISL_1273564* |
| *EPI_ISL_1273565* |
| *EPI_ISL_1273566* |
| *EPI_ISL_2180927* |
| *EPI_ISL_1273567* |
| *EPI_ISL_1273568* |
| *EPI_ISL_1273569* |
| *EPI_ISL_1273570* |
| *EPI_ISL_1273571* |

| |
|---|
| *EPI_ISL_1273572* |
| *EPI_ISL_1296822* |
| *EPI_ISL_1273573* |
| *EPI_ISL_1273574* |
| *EPI_ISL_1273575* |
| *EPI_ISL_1273576* |
| *EPI_ISL_2521383* |
| *EPI_ISL_1273577* |
| *EPI_ISL_1273578* |
| *EPI_ISL_2180929* |
| *EPI_ISL_1273579* |
| *EPI_ISL_1273580* |
| *EPI_ISL_1273750* |
| *EPI_ISL_1273581* |
| *EPI_ISL_1273582* |
| *EPI_ISL_1273583* |
| *EPI_ISL_1273584* |
| *EPI_ISL_1273585* |
| *EPI_ISL_1273586* |

| |
|---|
| EPI_ISL_1273587 |
| EPI_ISL_1273588 |
| EPI_ISL_1296824 |
| EPI_ISL_1273589 |
| EPI_ISL_1273590 |
| EPI_ISL_1273728 |
| EPI_ISL_1273591 |
| EPI_ISL_1273592 |
| EPI_ISL_1273593 |
| EPI_ISL_1273751 |
| EPI_ISL_1273762 |
| EPI_ISL_1273763 |
| EPI_ISL_1273594 |
| EPI_ISL_1273595 |
| EPI_ISL_1296825 |
| EPI_ISL_1273596 |
| EPI_ISL_1273752 |
| EPI_ISL_1273597 |
| EPI_ISL_2408424 |

| |
|---|
| EPI_ISL_2408425 |
| EPI_ISL_1296826 |
| EPI_ISL_1273598 |
| EPI_ISL_1273599 |
| EPI_ISL_1296828 |
| EPI_ISL_1273600 |
| EPI_ISL_1273601 |
| EPI_ISL_1273602 |
| EPI_ISL_1273603 |
| EPI_ISL_1273604 |
| EPI_ISL_1273605 |
| EPI_ISL_1273606 |
| EPI_ISL_1273607 |
| EPI_ISL_1273608 |
| EPI_ISL_1296829 |
| EPI_ISL_1993811 |
| EPI_ISL_1296830 |
| EPI_ISL_1273610 |
| EPI_ISL_1273611 |

| |
|---|
| EPI_ISL_1273612 |
| EPI_ISL_1273613 |
| EPI_ISL_1993812 |
| EPI_ISL_1273765 |
| EPI_ISL_1273766 |
| EPI_ISL_1296832 |
| EPI_ISL_1273753 |
| EPI_ISL_1273768 |
| EPI_ISL_1273614 |
| EPI_ISL_1273615 |
| EPI_ISL_1273616 |
| EPI_ISL_1273617 |
| EPI_ISL_1273618 |
| EPI_ISL_1296833 |
| EPI_ISL_1273619 |
| EPI_ISL_1273715 |
| EPI_ISL_1273756 |
| EPI_ISL_1273620 |
| EPI_ISL_1296834 |

| |
|---|
| *EPI_ISL_1273622* |
| *EPI_ISL_1273623* |
| *EPI_ISL_1273624* |
| *EPI_ISL_1273625* |
| *EPI_ISL_1273626* |
| *EPI_ISL_1273627* |
| *EPI_ISL_1273628* |
| *EPI_ISL_1273629* |
| *EPI_ISL_1273630* |
| *EPI_ISL_1014698* |
| *EPI_ISL_1014701* |
| *EPI_ISL_1273632* |
| *EPI_ISL_1273633* |
| *EPI_ISL_1273634* |
| *EPI_ISL_1296836* |
| *EPI_ISL_1273635* |
| *EPI_ISL_1273636* |
| *EPI_ISL_1273637* |
| *EPI_ISL_1273638* |

| |
|---|
| *EPI_ISL_1273639* |
| *EPI_ISL_1273640* |
| *EPI_ISL_1273709* |
| *EPI_ISL_1273641* |
| *EPI_ISL_1273642* |
| *EPI_ISL_1273643* |
| *EPI_ISL_1273644* |
| *EPI_ISL_1273645* |
| *EPI_ISL_1273646* |
| *EPI_ISL_1273647* |
| *EPI_ISL_1273648* |
| *EPI_ISL_1273649* |
| *EPI_ISL_1273654* |
| *EPI_ISL_1273655* |
| *EPI_ISL_1273656* |
| *EPI_ISL_1296837* |
| *EPI_ISL_1296838* |
| *EPI_ISL_1273657* |
| *EPI_ISL_1273658* |

| |
|---|
| *EPI_ISL_1273659* |
| *EPI_ISL_1273660* |
| *EPI_ISL_1296841* |
| *EPI_ISL_1273661* |
| *EPI_ISL_1273662* |
| *EPI_ISL_1273665* |
| *EPI_ISL_1273666* |
| *EPI_ISL_1273667* |
| *EPI_ISL_1273668* |
| *EPI_ISL_1273757* |
| *EPI_ISL_1273669* |
| *EPI_ISL_1273670* |
| *EPI_ISL_1014699* |
| *EPI_ISL_1014702* |
| *EPI_ISL_1014697* |
| *EPI_ISL_2466552* |
| *EPI_ISL_2466555* |
| *EPI_ISL_2466560* |
| *EPI_ISL_2466561* |

| |
|---|
| EPI_ISL_2484511 |
| EPI_ISL_2484388 |
| EPI_ISL_2484289 |
| EPI_ISL_2484279 |
| EPI_ISL_2484312 |
| EPI_ISL_2484281 |
| EPI_ISL_2484287 |
| EPI_ISL_2484374 |
| EPI_ISL_2484316 |
| EPI_ISL_2484488 |
| EPI_ISL_2484232 |
| EPI_ISL_2484341 |
| EPI_ISL_2484240 |
| EPI_ISL_2484348 |
| EPI_ISL_2484513 |
| EPI_ISL_2611018 |
| EPI_ISL_2611019 |
| EPI_ISL_2611020 |
| EPI_ISL_2611021 |

| |
|---|
| EPI_ISL_2611022 |
| EPI_ISL_2611023 |
| EPI_ISL_2611024 |
| EPI_ISL_2608292 |
| EPI_ISL_2611025 |
| EPI_ISL_2608293 |
| EPI_ISL_2608294 |
| EPI_ISL_2521386 |
| EPI_ISL_2521388 |
| EPI_ISL_2608295 |
| EPI_ISL_2608296 |
| EPI_ISL_2608297 |
| EPI_ISL_2608298 |
| EPI_ISL_2608299 |
| EPI_ISL_2608300 |
| EPI_ISL_1993813 |
| EPI_ISL_1993814 |
| EPI_ISL_1993815 |
| EPI_ISL_1993816 |

| |
|---|
| EPI_ISL_1993817 |
| EPI_ISL_1993818 |
| EPI_ISL_1993819 |
| EPI_ISL_1993820 |
| EPI_ISL_1993823 |
| EPI_ISL_1993824 |
| EPI_ISL_1993825 |
| EPI_ISL_1993826 |
| EPI_ISL_1993827 |
| EPI_ISL_1993830 |
| EPI_ISL_1993831 |
| EPI_ISL_1993876 |
| EPI_ISL_1993877 |
| EPI_ISL_1993879 |
| EPI_ISL_1993880 |
| EPI_ISL_1993882 |
| EPI_ISL_1914605 |
| EPI_ISL_1914606 |
| EPI_ISL_1747689 |

| | | |
|---|---|---|
| *EPI_ISL_1747772* | *EPI_ISL_2484375* | *EPI_ISL_1747619* |
| *EPI_ISL_1747661* | *EPI_ISL_1747651* | *EPI_ISL_2484507* |
| *EPI_ISL_1747725* | *EPI_ISL_1747645* | *EPI_ISL_2484408* |
| *EPI_ISL_1747993* | *EPI_ISL_1747635* | *EPI_ISL_2484317* |
| *EPI_ISL_1747833* | *EPI_ISL_1747631* | *EPI_ISL_2484469* |
| *EPI_ISL_1747926* | *EPI_ISL_1747638* | *EPI_ISL_2484352* |
| *EPI_ISL_1748194* | *EPI_ISL_1747637* | *EPI_ISL_2484349* |
| *EPI_ISL_1747709* | *EPI_ISL_1747621* | *EPI_ISL_2484416* |
| *EPI_ISL_1747960* | *EPI_ISL_1747642* | *EPI_ISL_2484308* |
| *EPI_ISL_1747665* | *EPI_ISL_2484397* | *EPI_ISL_2484487* |
| *EPI_ISL_1747662* | *EPI_ISL_2484275* | *EPI_ISL_2484252* |
| *EPI_ISL_1747784* | *EPI_ISL_2484517* | *EPI_ISL_2484504* |
| *EPI_ISL_1747684* | *EPI_ISL_2484370* | *EPI_ISL_2484336* |
| *EPI_ISL_1747756* | *EPI_ISL_1747628* | *EPI_ISL_2484347* |
| *EPI_ISL_1747658* | *EPI_ISL_1747649* | *EPI_ISL_2484339* |
| *EPI_ISL_1747776* | *EPI_ISL_1747646* | *EPI_ISL_2484360* |
| *EPI_ISL_1747714* | *EPI_ISL_1747629* | *EPI_ISL_2484514* |
| *EPI_ISL_1747740* | *EPI_ISL_1747650* | *EPI_ISL_2484266* |
| *EPI_ISL_2484250* | *EPI_ISL_2484439* | *EPI_ISL_2484268* |

| |
|---|
| EPI_ISL_2484342 |
| EPI_ISL_2484415 |
| EPI_ISL_2484315 |
| EPI_ISL_2484391 |
| EPI_ISL_2484277 |
| EPI_ISL_2484366 |
| EPI_ISL_2484321 |
| EPI_ISL_2484454 |
| EPI_ISL_2484313 |
| EPI_ISL_2102071 |
| EPI_ISL_2484390 |
| EPI_ISL_2521391 |
| EPI_ISL_2521392 |
| EPI_ISL_2521393 |
| EPI_ISL_2521394 |
| EPI_ISL_2521395 |
| EPI_ISL_2521396 |
| EPI_ISL_2102073 |
| EPI_ISL_2521397 |

| |
|---|
| EPI_ISL_2102074 |
| EPI_ISL_2102076 |
| EPI_ISL_2102077 |
| EPI_ISL_2102078 |
| EPI_ISL_2102079 |
| EPI_ISL_2102080 |
| EPI_ISL_2102081 |
| EPI_ISL_2102082 |
| EPI_ISL_2102083 |
| EPI_ISL_2102084 |
| EPI_ISL_2102085 |
| EPI_ISL_2102087 |
| EPI_ISL_2102088 |
| EPI_ISL_2180931 |
| EPI_ISL_2102089 |
| EPI_ISL_2180932 |
| EPI_ISL_2180934 |
| EPI_ISL_2180935 |
| EPI_ISL_2180937 |

| |
|---|
| EPI_ISL_2180940 |
| EPI_ISL_2180942 |
| EPI_ISL_1747625 |
| EPI_ISL_1747814 |
| EPI_ISL_1747853 |
| EPI_ISL_1747826 |
| EPI_ISL_1747899 |
| EPI_ISL_1747892 |
| EPI_ISL_1747641 |
| EPI_ISL_1747622 |
| EPI_ISL_1747636 |
| EPI_ISL_1748287 |
| EPI_ISL_1748278 |
| EPI_ISL_1748256 |
| EPI_ISL_1748266 |
| EPI_ISL_1748319 |
| EPI_ISL_1748302 |
| EPI_ISL_1748310 |
| EPI_ISL_1748269 |

| |
|---|
| *EPI_ISL_1748026* |
| *EPI_ISL_1747998* |
| *EPI_ISL_1748002* |
| *EPI_ISL_1748042* |
| *EPI_ISL_1748137* |
| *EPI_ISL_1748080* |
| *EPI_ISL_1748037* |
| *EPI_ISL_1748088* |
| *EPI_ISL_1748164* |
| *EPI_ISL_1748060* |
| *EPI_ISL_1747995* |
| *EPI_ISL_1748366* |
| *EPI_ISL_1748351* |
| *EPI_ISL_1748338* |
| *EPI_ISL_1748419* |
| *EPI_ISL_1748387* |
| *EPI_ISL_1748412* |
| *EPI_ISL_1748359* |
| *EPI_ISL_1748343* |

| |
|---|
| *EPI_ISL_1748409* |
| *EPI_ISL_1748364* |
| *EPI_ISL_1748336* |
| *EPI_ISL_1748382* |
| *EPI_ISL_1748384* |
| *EPI_ISL_1748368* |
| *EPI_ISL_1748348* |
| *EPI_ISL_1748371* |
| *EPI_ISL_1748362* |
| *EPI_ISL_1748353* |
| *EPI_ISL_1748395* |
| *EPI_ISL_1748332* |
| *EPI_ISL_1748483* |
| *EPI_ISL_1748490* |
| *EPI_ISL_1748430* |
| *EPI_ISL_1748493* |
| *EPI_ISL_1748468* |
| *EPI_ISL_1748452* |
| *EPI_ISL_1748378* |

| |
|---|
| *EPI_ISL_1748341* |
| *EPI_ISL_1748391* |
| *EPI_ISL_1748437* |
| *EPI_ISL_1748495* |
| *EPI_ISL_1748441* |
| *EPI_ISL_1748474* |
| *EPI_ISL_1748459* |
| *EPI_ISL_1748422* |
| *EPI_ISL_1748427* |
| *EPI_ISL_1748486* |
| *EPI_ISL_1748649* |
| *EPI_ISL_1748527* |
| *EPI_ISL_1748660* |
| *EPI_ISL_1748509* |
| *EPI_ISL_1748677* |
| *EPI_ISL_1748570* |
| *EPI_ISL_1748642* |
| *EPI_ISL_1748587* |
| *EPI_ISL_1748725* |

| |
|---|
| *EPI_ISL_1748614* |
| *EPI_ISL_1748694* |
| *EPI_ISL_1748732* |
| *EPI_ISL_1748543* |
| *EPI_ISL_1748522* |
| *EPI_ISL_1748604* |
| *EPI_ISL_1748644* |
| *EPI_ISL_1748668* |
| *EPI_ISL_1748685* |
| *EPI_ISL_1748568* |
| *EPI_ISL_1748640* |
| *EPI_ISL_1748516* |
| *EPI_ISL_1748618* |
| *EPI_ISL_1748734* |
| *EPI_ISL_1748730* |
| *EPI_ISL_1748714* |
| *EPI_ISL_1748539* |
| *EPI_ISL_1748681* |
| *EPI_ISL_1748655* |

| |
|---|
| *EPI_ISL_1748534* |
| *EPI_ISL_1748703* |
| *EPI_ISL_1748673* |
| *EPI_ISL_1748634* |
| *EPI_ISL_1748580* |
| *EPI_ISL_1748563* |
| *EPI_ISL_1748518* |
| *EPI_ISL_1748711* |
| *EPI_ISL_1748647* |
| *EPI_ISL_1747859* |
| *EPI_ISL_1748499* |
| *EPI_ISL_1748463* |
| *EPI_ISL_1833641* |
| *EPI_ISL_1833649* |
| *EPI_ISL_1833652* |
| *EPI_ISL_1833656* |
| *EPI_ISL_1833661* |
| *EPI_ISL_1833663* |
| *EPI_ISL_1833668* |

| |
|---|
| *EPI_ISL_1833673* |
| *EPI_ISL_1833676* |
| *EPI_ISL_1833679* |
| *EPI_ISL_1833682* |
| *EPI_ISL_1833684* |
| *EPI_ISL_1833687* |
| *EPI_ISL_1833690* |
| *EPI_ISL_1833692* |
| *EPI_ISL_1833695* |
| *EPI_ISL_1833698* |
| *EPI_ISL_1833701* |
| *EPI_ISL_1833704* |
| *EPI_ISL_1973557* |
| *EPI_ISL_1973558* |
| *EPI_ISL_1973559* |
| *EPI_ISL_1973562* |
| *EPI_ISL_1973563* |
| *EPI_ISL_1973564* |
| *EPI_ISL_1973565* |

| |
|---|
| EPI_ISL_1833709 |
| EPI_ISL_1833711 |
| EPI_ISL_1993832 |
| EPI_ISL_1993833 |
| EPI_ISL_1993883 |
| EPI_ISL_1833714 |
| EPI_ISL_1833718 |
| EPI_ISL_1833721 |
| EPI_ISL_1993884 |
| EPI_ISL_1993885 |
| EPI_ISL_1993886 |
| EPI_ISL_1993887 |
| EPI_ISL_1993888 |
| EPI_ISL_1993890 |
| EPI_ISL_1993891 |
| EPI_ISL_1993892 |
| EPI_ISL_1993893 |
| EPI_ISL_1993834 |
| EPI_ISL_1833726 |

| |
|---|
| EPI_ISL_1833729 |
| EPI_ISL_1973566 |
| EPI_ISL_1993835 |
| EPI_ISL_1833731 |
| EPI_ISL_1993894 |
| EPI_ISL_1993895 |
| EPI_ISL_1993836 |
| EPI_ISL_1993896 |
| EPI_ISL_1993897 |
| EPI_ISL_1993837 |
| EPI_ISL_1993898 |
| EPI_ISL_1993899 |
| EPI_ISL_1993838 |
| EPI_ISL_1993900 |
| EPI_ISL_1993901 |
| EPI_ISL_1993902 |
| EPI_ISL_1993904 |
| EPI_ISL_1993839 |
| EPI_ISL_2622118 |

| |
|---|
| EPI_ISL_2622119 |
| EPI_ISL_2622120 |
| EPI_ISL_1833741 |
| EPI_ISL_2622121 |
| EPI_ISL_2622122 |
| EPI_ISL_2622123 |
| EPI_ISL_2622124 |
| EPI_ISL_2622125 |
| EPI_ISL_1993906 |
| EPI_ISL_2622126 |
| EPI_ISL_2622127 |
| EPI_ISL_2622128 |
| EPI_ISL_2622129 |
| EPI_ISL_2622130 |
| EPI_ISL_2622131 |
| EPI_ISL_1993840 |
| EPI_ISL_2622132 |
| EPI_ISL_1993908 |
| EPI_ISL_1993841 |

| |
|---|
| *EPI_ISL_1993842* |
| *EPI_ISL_1993843* |
| *EPI_ISL_1993844* |
| *EPI_ISL_1993845* |
| *EPI_ISL_1993846* |
| *EPI_ISL_1993847* |
| *EPI_ISL_1993909* |
| *EPI_ISL_1993848* |
| *EPI_ISL_1993849* |
| *EPI_ISL_1993850* |
| *EPI_ISL_1993910* |
| *EPI_ISL_1993912* |
| *EPI_ISL_1993913* |
| *EPI_ISL_1993851* |
| *EPI_ISL_1993852* |
| *EPI_ISL_1993914* |
| *EPI_ISL_1993915* |
| *EPI_ISL_1993853* |
| *EPI_ISL_1993916* |

| |
|---|
| *EPI_ISL_1993854* |
| *EPI_ISL_1993855* |
| *EPI_ISL_1993856* |
| *EPI_ISL_1993857* |
| *EPI_ISL_1993859* |
| *EPI_ISL_1993860* |
| *EPI_ISL_2622133* |
| *EPI_ISL_1993917* |
| *EPI_ISL_1993918* |
| *EPI_ISL_2102091* |
| *EPI_ISL_2622134* |
| *EPI_ISL_2102092* |
| *EPI_ISL_2102093* |
| *EPI_ISL_2102094* |
| *EPI_ISL_2102095* |
| *EPI_ISL_2180946* |
| *EPI_ISL_2102096* |
| *EPI_ISL_2102097* |
| *EPI_ISL_2102099* |

| |
|---|
| *EPI_ISL_2102100* |
| *EPI_ISL_2102101* |
| *EPI_ISL_2180947* |
| *EPI_ISL_2102102* |
| *EPI_ISL_2180949* |
| *EPI_ISL_2102103* |
| *EPI_ISL_2180950* |
| *EPI_ISL_2102104* |
| *EPI_ISL_2180952* |
| *EPI_ISL_2180954* |
| *EPI_ISL_2180955* |
| *EPI_ISL_2102105* |
| *EPI_ISL_2180957* |
| *EPI_ISL_2180959* |
| *EPI_ISL_2102106* |
| *EPI_ISL_2102108* |
| *EPI_ISL_2102109* |
| *EPI_ISL_2180960* |
| *EPI_ISL_2180962* |

| |
|---|
| EPI_ISL_2367220 |
| EPI_ISL_2367221 |
| EPI_ISL_2367223 |
| EPI_ISL_2180963 |
| EPI_ISL_2367224 |
| EPI_ISL_2180965 |
| EPI_ISL_2180967 |
| EPI_ISL_2180968 |
| EPI_ISL_2180970 |
| EPI_ISL_2180972 |
| EPI_ISL_2180973 |
| EPI_ISL_2180975 |
| EPI_ISL_2180977 |
| EPI_ISL_2180978 |
| EPI_ISL_2180980 |
| EPI_ISL_2180981 |
| EPI_ISL_2180983 |
| EPI_ISL_2180985 |
| EPI_ISL_2180986 |

| |
|---|
| EPI_ISL_2270038 |
| EPI_ISL_2270040 |
| EPI_ISL_2270041 |
| EPI_ISL_2270043 |
| EPI_ISL_2270045 |
| EPI_ISL_2270046 |
| EPI_ISL_2270048 |
| EPI_ISL_2270049 |
| EPI_ISL_2270051 |
| EPI_ISL_2270053 |
| EPI_ISL_2270054 |
| EPI_ISL_2270057 |
| EPI_ISL_2270059 |
| EPI_ISL_2270060 |
| EPI_ISL_2270062 |
| EPI_ISL_2270064 |
| EPI_ISL_2270065 |
| EPI_ISL_2180990 |
| EPI_ISL_2270068 |

| |
|---|
| EPI_ISL_2180992 |
| EPI_ISL_2180994 |
| EPI_ISL_2367225 |
| EPI_ISL_2180995 |
| EPI_ISL_2180997 |
| EPI_ISL_2367226 |
| EPI_ISL_2270112 |
| EPI_ISL_2270113 |
| EPI_ISL_2270115 |
| EPI_ISL_2270117 |
| EPI_ISL_2270118 |
| EPI_ISL_2367227 |
| EPI_ISL_2270120 |
| EPI_ISL_2270121 |
| EPI_ISL_2270123 |
| EPI_ISL_2270125 |
| EPI_ISL_2270126 |
| EPI_ISL_2270128 |
| EPI_ISL_2270129 |

| |
|---|
| EPI_ISL_2270131 |
| EPI_ISL_2270133 |
| EPI_ISL_2270134 |
| EPI_ISL_2270136 |
| EPI_ISL_2270137 |
| EPI_ISL_2270139 |
| EPI_ISL_2270141 |
| EPI_ISL_2270142 |
| EPI_ISL_2270144 |
| EPI_ISL_2270145 |
| EPI_ISL_2270147 |
| EPI_ISL_2270149 |
| EPI_ISL_2270150 |
| EPI_ISL_2270152 |
| EPI_ISL_2270153 |
| EPI_ISL_2270155 |
| EPI_ISL_2270157 |
| EPI_ISL_2270070 |
| EPI_ISL_2270077 |

| |
|---|
| EPI_ISL_2270079 |
| EPI_ISL_2270081 |
| EPI_ISL_2270084 |
| EPI_ISL_2270165 |
| EPI_ISL_2270085 |
| EPI_ISL_2270086 |
| EPI_ISL_2408427 |
| EPI_ISL_2367228 |
| EPI_ISL_2367229 |
| EPI_ISL_2270095 |
| EPI_ISL_2270096 |
| EPI_ISL_2270098 |
| EPI_ISL_2610954 |
| EPI_ISL_2367230 |
| EPI_ISL_2610955 |
| EPI_ISL_2367231 |
| EPI_ISL_2408428 |
| EPI_ISL_2367232 |
| EPI_ISL_2408429 |

| |
|---|
| EPI_ISL_2367233 |
| EPI_ISL_2408430 |
| EPI_ISL_2367234 |
| EPI_ISL_2408431 |
| EPI_ISL_2367235 |
| EPI_ISL_2367236 |
| EPI_ISL_2367237 |
| EPI_ISL_2466563 |
| EPI_ISL_2367238 |
| EPI_ISL_2408432 |
| EPI_ISL_2367239 |
| EPI_ISL_2408433 |
| EPI_ISL_2367240 |
| EPI_ISL_2367241 |
| EPI_ISL_2466564 |
| EPI_ISL_2367242 |
| EPI_ISL_2367243 |
| EPI_ISL_2367245 |
| EPI_ISL_2367246 |

| |
|---|
| *EPI_ISL_2367247* |
| *EPI_ISL_2367248* |
| *EPI_ISL_2408434* |
| *EPI_ISL_2367249* |
| *EPI_ISL_2408435* |
| *EPI_ISL_2367250* |
| *EPI_ISL_2466565* |
| *EPI_ISL_2367252* |
| *EPI_ISL_2408437* |
| *EPI_ISL_2367253* |
| *EPI_ISL_2408438* |
| *EPI_ISL_2408440* |
| *EPI_ISL_2367255* |
| *EPI_ISL_2408441* |
| *EPI_ISL_2367257* |
| *EPI_ISL_2408442* |
| *EPI_ISL_2408443* |
| *EPI_ISL_2466566* |
| *EPI_ISL_2408445* |

| |
|---|
| *EPI_ISL_2408446* |
| *EPI_ISL_2466567* |
| *EPI_ISL_2466568* |
| *EPI_ISL_2408448* |
| *EPI_ISL_2408449* |
| *EPI_ISL_2466569* |
| *EPI_ISL_2408450* |
| *EPI_ISL_2408451* |
| *EPI_ISL_2408452* |
| *EPI_ISL_2408453* |
| *EPI_ISL_2408454* |
| *EPI_ISL_2408455* |
| *EPI_ISL_2408456* |
| *EPI_ISL_2408457* |
| *EPI_ISL_2408458* |
| *EPI_ISL_2408459* |
| *EPI_ISL_2408460* |
| *EPI_ISL_2408461* |
| *EPI_ISL_2408462* |

| |
|---|
| *EPI_ISL_2408463* |
| *EPI_ISL_2408464* |
| *EPI_ISL_2466570* |
| *EPI_ISL_2466571* |
| *EPI_ISL_2466572* |
| *EPI_ISL_2466573* |
| *EPI_ISL_2466574* |
| *EPI_ISL_2466575* |
| *EPI_ISL_2466576* |
| *EPI_ISL_2408470* |
| *EPI_ISL_2466577* |
| *EPI_ISL_2466578* |
| *EPI_ISL_2408471* |
| *EPI_ISL_2466592* |
| *EPI_ISL_2610956* |
| *EPI_ISL_2466593* |
| *EPI_ISL_2610957* |
| *EPI_ISL_2466594* |
| *EPI_ISL_2466595* |

| |
|---|
| *EPI_ISL_2466596* |
| *EPI_ISL_2466597* |
| *EPI_ISL_2611026* |
| *EPI_ISL_2466583* |
| *EPI_ISL_2610958* |
| *EPI_ISL_2466600* |
| *EPI_ISL_2466601* |
| *EPI_ISL_2466602* |
| *EPI_ISL_2466603* |
| *EPI_ISL_2610961* |
| *EPI_ISL_2610962* |
| *EPI_ISL_2466604* |
| *EPI_ISL_2466605* |
| *EPI_ISL_2466606* |
| *EPI_ISL_2610963* |
| *EPI_ISL_2610964* |
| *EPI_ISL_2610965* |
| *EPI_ISL_2610990* |
| *EPI_ISL_2610966* |

| |
|---|
| *EPI_ISL_2610967* |
| *EPI_ISL_2622136* |
| *EPI_ISL_2610968* |
| *EPI_ISL_2610969* |
| *EPI_ISL_2611035* |
| *EPI_ISL_830979* |
| *EPI_ISL_830980* |
| *EPI_ISL_830981* |
| *EPI_ISL_830982* |
| *EPI_ISL_830983* |
| *EPI_ISL_930908* |
| *EPI_ISL_930965* |
| *EPI_ISL_930966* |
| *EPI_ISL_930968* |
| *EPI_ISL_931032* |
| *EPI_ISL_931083* |
| *EPI_ISL_931198* |
| *EPI_ISL_931243* |
| *EPI_ISL_931244* |

| |
|---|
| *EPI_ISL_931298* |
| *EPI_ISL_931299* |
| *EPI_ISL_931371* |
| *EPI_ISL_931372* |
| *EPI_ISL_931373* |
| *EPI_ISL_931374* |
| *EPI_ISL_3344128* |
| *EPI_ISL_1273467* |
| *EPI_ISL_1273468* |
| *EPI_ISL_2611046* |
| *EPI_ISL_2611047* |
| *EPI_ISL_1273475* |
| *EPI_ISL_2611048* |
| *EPI_ISL_1273476* |
| *EPI_ISL_1273477* |
| *EPI_ISL_1273478* |
| *EPI_ISL_1273479* |
| *EPI_ISL_1273484* |
| *EPI_ISL_1273490* |

| |
|---|
| *EPI_ISL_931449* |
| *EPI_ISL_931450* |
| *EPI_ISL_1273502* |
| *EPI_ISL_1273515* |
| *EPI_ISL_2367258* |
| *EPI_ISL_1273516* |
| *EPI_ISL_1273517* |
| *EPI_ISL_1273609* |
| *EPI_ISL_942515* |
| *EPI_ISL_1273621* |
| *EPI_ISL_1014696* |
| *EPI_ISL_1273631* |
| *EPI_ISL_2521398* |
| *EPI_ISL_2626178* |
| *EPI_ISL_1273650* |
| *EPI_ISL_1273651* |
| *EPI_ISL_1273652* |
| *EPI_ISL_1273754* |
| *EPI_ISL_1273663* |

| |
|---|
| *EPI_ISL_1273664* |
| *EPI_ISL_1273671* |
| *EPI_ISL_1296840* |
| *EPI_ISL_1273672* |
| *EPI_ISL_1273673* |
| *EPI_ISL_1014694* |
| *EPI_ISL_1014695* |
| *EPI_ISL_1034152* |
| *EPI_ISL_1014700* |
| *EPI_ISL_1993861* |
| *EPI_ISL_1993862* |
| *EPI_ISL_1993863* |
| *EPI_ISL_1993864* |
| *EPI_ISL_1993865* |
| *EPI_ISL_1993866* |
| *EPI_ISL_1993919* |
| *EPI_ISL_1914608* |
| *EPI_ISL_1747944* |
| *EPI_ISL_1747639* |

| |
|---|
| *EPI_ISL_1747633* |
| *EPI_ISL_1747627* |
| *EPI_ISL_2484290* |
| *EPI_ISL_2484486* |
| *EPI_ISL_2484274* |
| *EPI_ISL_2484412* |
| *EPI_ISL_2102117* |
| *EPI_ISL_2180999* |
| *EPI_ISL_1747929* |
| *EPI_ISL_1748290* |
| *EPI_ISL_1748254* |
| *EPI_ISL_1748295* |
| *EPI_ISL_1748160* |
| *EPI_ISL_1748007* |
| *EPI_ISL_1748407* |
| *EPI_ISL_1748328* |
| *EPI_ISL_1748400* |
| *EPI_ISL_1748416* |
| *EPI_ISL_1748471* |

| |
|---|
| *EPI_ISL_1748445* |
| *EPI_ISL_1748689* |
| *EPI_ISL_1748698* |
| *EPI_ISL_1748531* |
| *EPI_ISL_1748622* |
| *EPI_ISL_1748585* |
| *EPI_ISL_1748609* |
| *EPI_ISL_1748592* |
| *EPI_ISL_1748600* |
| *EPI_ISL_1748550* |
| *EPI_ISL_1748554* |
| *EPI_ISL_1748631* |
| *EPI_ISL_1973568* |
| *EPI_ISL_1833746* |
| *EPI_ISL_1993920* |
| *EPI_ISL_1833748* |
| *EPI_ISL_1993921* |
| *EPI_ISL_1993867* |
| *EPI_ISL_1993868* |

| |
|---|
| *EPI_ISL_1993922* |
| *EPI_ISL_1833751* |
| *EPI_ISL_1993923* |
| *EPI_ISL_1993869* |
| *EPI_ISL_2622151* |
| *EPI_ISL_2622152* |
| *EPI_ISL_2622153* |
| *EPI_ISL_2622154* |
| *EPI_ISL_1993870* |
| *EPI_ISL_1993871* |
| *EPI_ISL_1993872* |
| *EPI_ISL_2622155* |
| *EPI_ISL_2622156* |
| *EPI_ISL_2622157* |
| *EPI_ISL_2622158* |
| *EPI_ISL_1993873* |
| *EPI_ISL_2622159* |
| *EPI_ISL_2102119* |
| *EPI_ISL_2102120* |

| |
|---|
| *EPI_ISL_2181002* |
| *EPI_ISL_2270101* |
| *EPI_ISL_2270102* |
| *EPI_ISL_2270104* |
| *EPI_ISL_2181004* |
| *EPI_ISL_2270106* |
| *EPI_ISL_2408473* |
| *EPI_ISL_2367259* |
| *EPI_ISL_2466589* |
| *EPI_ISL_2367260* |
| *EPI_ISL_2466590* |
| *EPI_ISL_2408474* |
| *EPI_ISL_2408475* |
| *EPI_ISL_2611003* |
| *EPI_ISL_2466591* |
| *EPI_ISL_2484305* |
| *EPI_ISL_1273653* |
| *EPI_ISL_2484394* |
| *EPI_ISL_2484320* |

| |
|---|
| *EPI_ISL_2484515* |
| *EPI_ISL_2484516* |
| *EPI_ISL_2484447* |
| *EPI_ISL_1747953* |
| *EPI_ISL_1747935* |
| *EPI_ISL_1748107* |
| *EPI_ISL_1748324* |
| *EPI_ISL_1993934* |
| *EPI_ISL_1993941* |
| *EPI_ISL_2181005* |
| *EPI_ISL_2466607* |
| *EPI_ISL_2274447* |
| EPI_ISL_466927 |
| EPI_ISL_466931 |
| EPI_ISL_466939 |
| EPI_ISL_466940 |
| EPI_ISL_466941 |
| EPI_ISL_466942 |
| EPI_ISL_466944 |

| |
|---|
| EPI_ISL_466946 |
| EPI_ISL_466949 |
| EPI_ISL_466951 |
| EPI_ISL_466952 |
| EPI_ISL_466953 |
| EPI_ISL_466954 |
| EPI_ISL_466958 |
| EPI_ISL_466960 |
| EPI_ISL_466985 |
| EPI_ISL_451675 |
| EPI_ISL_451695 |
| EPI_ISL_451699 |
| EPI_ISL_451700 |
| EPI_ISL_451703 |
| EPI_ISL_451713 |
| EPI_ISL_451716 |
| EPI_ISL_451728 |
| EPI_ISL_451732 |
| EPI_ISL_574905 |

| |
|---|
| EPI_ISL_574906 |
| EPI_ISL_574907 |
| EPI_ISL_574908 |
| EPI_ISL_574909 |
| EPI_ISL_574910 |
| EPI_ISL_574911 |
| EPI_ISL_1130016 |
| EPI_ISL_574912 |
| EPI_ISL_451772 |
| EPI_ISL_451774 |
| EPI_ISL_451778 |
| EPI_ISL_451803 |
| EPI_ISL_451814 |
| EPI_ISL_451820 |
| EPI_ISL_451841 |
| EPI_ISL_486442 |
| EPI_ISL_486443 |
| EPI_ISL_486445 |
| EPI_ISL_486456 |

| |
|---|
| EPI_ISL_486473 |
| EPI_ISL_486474 |
| EPI_ISL_451874 |
| EPI_ISL_451885 |
| EPI_ISL_451895 |
| EPI_ISL_451897 |
| EPI_ISL_451899 |
| EPI_ISL_468210 |
| EPI_ISL_468213 |
| EPI_ISL_468221 |
| EPI_ISL_468225 |
| EPI_ISL_468254 |
| EPI_ISL_467001 |
| EPI_ISL_467012 |
| EPI_ISL_476108 |
| EPI_ISL_486538 |
| EPI_ISL_500941 |
| EPI_ISL_500944 |
| EPI_ISL_510739 |

| |
|---|
| EPI_ISL_510747 |
| EPI_ISL_512043 |
| EPI_ISL_516563 |
| EPI_ISL_1130040 |
| EPI_ISL_523907 |
| EPI_ISL_523925 |
| EPI_ISL_535585 |
| EPI_ISL_535587 |
| EPI_ISL_535610 |
| EPI_ISL_1130001 |
| EPI_ISL_535629 |
| EPI_ISL_535632 |
| EPI_ISL_535636 |
| EPI_ISL_535637 |
| EPI_ISL_535640 |
| EPI_ISL_535649 |
| EPI_ISL_539377 |
| EPI_ISL_539378 |
| EPI_ISL_541463 |

| |
|---|
| EPI_ISL_560470 |
| EPI_ISL_560471 |
| EPI_ISL_560472 |
| EPI_ISL_1130013 |
| EPI_ISL_560473 |
| EPI_ISL_560474 |
| EPI_ISL_603343 |
| EPI_ISL_603356 |
| EPI_ISL_603420 |
| EPI_ISL_603421 |
| EPI_ISL_603620 |
| EPI_ISL_603682 |
| EPI_ISL_603732 |
| EPI_ISL_1496513 |
| EPI_ISL_1496524 |
| EPI_ISL_1496520 |
| EPI_ISL_614899 |
| EPI_ISL_614900 |
| EPI_ISL_614901 |

| |
|---|
| EPI_ISL_614902 |
| EPI_ISL_614903 |
| EPI_ISL_614904 |
| EPI_ISL_614905 |
| EPI_ISL_614906 |
| EPI_ISL_1496515 |
| EPI_ISL_1496518 |
| EPI_ISL_1496521 |
| EPI_ISL_1496523 |
| EPI_ISL_1496516 |
| EPI_ISL_1496514 |
| EPI_ISL_1496262 |
| EPI_ISL_1496280 |
| EPI_ISL_1496522 |
| EPI_ISL_1496517 |
| EPI_ISL_2724424 |
| EPI_ISL_2724423 |
| EPI_ISL_2724426 |
| EPI_ISL_2938392 |

| |
|---|
| EPI_ISL_3296240 |
| EPI_ISL_3007559 |
| EPI_ISL_2938386 |
| EPI_ISL_3007561 |
| EPI_ISL_2938390 |
| EPI_ISL_2938394 |
| EPI_ISL_2938389 |
| EPI_ISL_2938393 |
| EPI_ISL_2938395 |
| EPI_ISL_2938388 |
| EPI_ISL_2938387 |
| EPI_ISL_2938396 |
| EPI_ISL_2938385 |
| EPI_ISL_2938391 |
| EPI_ISL_3465251 |
| EPI_ISL_3465308 |
| EPI_ISL_3465280 |
| EPI_ISL_3465269 |
| EPI_ISL_3465290 |

| |
|---|
| EPI_ISL_3296248 |
| EPI_ISL_3296292 |
| EPI_ISL_3296302 |
| EPI_ISL_3296300 |
| EPI_ISL_3296250 |
| EPI_ISL_3296209 |
| EPI_ISL_3296205 |
| EPI_ISL_3296203 |
| EPI_ISL_3296298 |
| EPI_ISL_3296194 |
| EPI_ISL_3296232 |
| EPI_ISL_3296238 |
| EPI_ISL_3296228 |
| EPI_ISL_3296304 |
| EPI_ISL_3296308 |
| EPI_ISL_3296290 |
| EPI_ISL_3296286 |
| EPI_ISL_3296224 |
| EPI_ISL_3296219 |

| |
|---|
| EPI_ISL_3296265 |
| EPI_ISL_3296260 |
| EPI_ISL_3296221 |
| EPI_ISL_3296242 |
| EPI_ISL_3296230 |
| EPI_ISL_3296207 |
| EPI_ISL_3296254 |
| EPI_ISL_3296199 |
| EPI_ISL_3296271 |
| EPI_ISL_3296246 |
| EPI_ISL_3296263 |
| EPI_ISL_3296196 |
| EPI_ISL_3296257 |
| EPI_ISL_3296269 |
| EPI_ISL_3296217 |
| EPI_ISL_3296252 |
| EPI_ISL_3296267 |
| EPI_ISL_3296296 |
| EPI_ISL_3296226 |

| |
|---|
| EPI_ISL_3296279 |
| EPI_ISL_3296275 |
| EPI_ISL_3296236 |
| EPI_ISL_3296234 |
| EPI_ISL_3296288 |
| EPI_ISL_3296201 |
| EPI_ISL_3296306 |
| EPI_ISL_3296215 |
| EPI_ISL_3296284 |
| EPI_ISL_3296273 |
| EPI_ISL_3296277 |
| EPI_ISL_3296294 |
| EPI_ISL_3296211 |
| EPI_ISL_3296310 |
| EPI_ISL_3296213 |
| EPI_ISL_3296258 |
| EPI_ISL_3296281 |
| EPI_ISL_3296244 |
| EPI_ISL_3345910 |

| |
|---|
| EPI_ISL_3345900 |
| EPI_ISL_3345889 |
| EPI_ISL_3345915 |
| EPI_ISL_3345904 |
| EPI_ISL_3345895 |
| EPI_ISL_3345909 |
| EPI_ISL_3345877 |
| EPI_ISL_3345931 |
| EPI_ISL_3345892 |
| EPI_ISL_4486606 |
| EPI_ISL_4486610 |
| EPI_ISL_4486618 |
| EPI_ISL_4486611 |
| EPI_ISL_4486617 |
| EPI_ISL_4486615 |
| EPI_ISL_3345882 |
| EPI_ISL_3345901 |
| EPI_ISL_3345876 |
| EPI_ISL_4486608 |

| |
|---|
| EPI_ISL_3345922 |
| EPI_ISL_3345914 |
| EPI_ISL_4486613 |
| EPI_ISL_3345906 |
| EPI_ISL_3345928 |
| EPI_ISL_3345881 |
| EPI_ISL_3345874 |
| EPI_ISL_3345933 |
| EPI_ISL_3345907 |
| EPI_ISL_3345879 |
| EPI_ISL_3345926 |
| EPI_ISL_3345925 |
| EPI_ISL_3345885 |
| EPI_ISL_3345923 |
| EPI_ISL_3345917 |
| EPI_ISL_3345903 |
| EPI_ISL_3345893 |
| EPI_ISL_3345912 |
| EPI_ISL_3345884 |

| |
|---|
| EPI_ISL_3345896 |
| EPI_ISL_3345898 |
| EPI_ISL_3345918 |
| EPI_ISL_3345890 |
| EPI_ISL_3345929 |
| EPI_ISL_3345887 |
| EPI_ISL_3476209 |
| EPI_ISL_3476211 |
| EPI_ISL_3345920 |
| EPI_ISL_3476202 |
| EPI_ISL_3476203 |
| EPI_ISL_3476205 |
| EPI_ISL_3476208 |
| EPI_ISL_3476207 |
| EPI_ISL_3476214 |
| EPI_ISL_3476201 |
| EPI_ISL_3476206 |
| EPI_ISL_3476204 |
| EPI_ISL_3476213 |

| |
|---|
| EPI_ISL_3476210 |
| EPI_ISL_3465283 |
| EPI_ISL_3476212 |
| EPI_ISL_3465289 |
| EPI_ISL_3465303 |
| EPI_ISL_3465294 |
| EPI_ISL_3465262 |
| EPI_ISL_3465267 |
| EPI_ISL_3465295 |
| EPI_ISL_3465271 |
| EPI_ISL_3465255 |
| EPI_ISL_3465273 |
| EPI_ISL_3465266 |
| EPI_ISL_3465243 |
| EPI_ISL_3465301 |
| EPI_ISL_3465246 |
| EPI_ISL_3465299 |
| EPI_ISL_3465244 |
| EPI_ISL_3465287 |

| | | |
|---|---|---|
| EPI_ISL_3465249 | EPI_ISL_3854236 | EPI_ISL_4233539 |
| EPI_ISL_3465292 | EPI_ISL_3854234 | EPI_ISL_4025398 |
| EPI_ISL_3465241 | EPI_ISL_3854242 | EPI_ISL_4025399 |
| EPI_ISL_3465297 | EPI_ISL_3854252 | EPI_ISL_4025400 |
| EPI_ISL_3465304 | EPI_ISL_3854262 | EPI_ISL_4025401 |
| EPI_ISL_3465253 | EPI_ISL_3854257 | EPI_ISL_3854237 |
| EPI_ISL_3465276 | EPI_ISL_4233541 | EPI_ISL_3854232 |
| EPI_ISL_3465274 | EPI_ISL_4233535 | EPI_ISL_3854250 |
| EPI_ISL_3465306 | EPI_ISL_4233536 | EPI_ISL_3854245 |
| EPI_ISL_3465285 | EPI_ISL_4233529 | EPI_ISL_3854247 |
| EPI_ISL_3465281 | EPI_ISL_4233540 | EPI_ISL_3854244 |
| EPI_ISL_3465260 | EPI_ISL_4233528 | EPI_ISL_3854253 |
| EPI_ISL_3854239 | EPI_ISL_4233538 | EPI_ISL_4318024 |
| EPI_ISL_3465258 | EPI_ISL_4233530 | EPI_ISL_4233999 |
| EPI_ISL_3465264 | EPI_ISL_4233532 | EPI_ISL_4233990 |
| EPI_ISL_3465278 | EPI_ISL_4233533 | EPI_ISL_4233992 |
| EPI_ISL_3465248 | EPI_ISL_4233531 | EPI_ISL_4318030 |
| EPI_ISL_3465256 | EPI_ISL_4233537 | EPI_ISL_4236355 |
| EPI_ISL_3854248 | EPI_ISL_4233534 | EPI_ISL_4236356 |

| | | |
|---|---|---|
| EPI_ISL_4236354 | EPI_ISL_4233984 | EPI_ISL_4233989 |
| EPI_ISL_4236340 | EPI_ISL_4233988 | EPI_ISL_4233996 |
| EPI_ISL_4236347 | EPI_ISL_4234001 | EPI_ISL_4233982 |
| EPI_ISL_4234002 | EPI_ISL_4318027 | EPI_ISL_4236352 |
| EPI_ISL_4318005 | EPI_ISL_4316074 | EPI_ISL_4316075 |
| EPI_ISL_4236349 | EPI_ISL_4236336 | EPI_ISL_4233987 |
| EPI_ISL_4236344 | EPI_ISL_4236337 | EPI_ISL_4233995 |
| EPI_ISL_4236341 | EPI_ISL_4317999 | EPI_ISL_4233983 |
| EPI_ISL_4236345 | EPI_ISL_4236343 | EPI_ISL_4316084 |
| EPI_ISL_4236348 | EPI_ISL_4317970 | EPI_ISL_4316072 |
| EPI_ISL_4236350 | EPI_ISL_4236333 | EPI_ISL_4316086 |
| EPI_ISL_4236339 | EPI_ISL_4236334 | EPI_ISL_4316067 |
| EPI_ISL_4236346 | EPI_ISL_4233997 | EPI_ISL_4316092 |
| EPI_ISL_4236338 | EPI_ISL_4233985 | EPI_ISL_4316090 |
| EPI_ISL_4236335 | EPI_ISL_4233986 | EPI_ISL_4316088 |
| EPI_ISL_4236353 | EPI_ISL_4233994 | EPI_ISL_4316070 |
| EPI_ISL_4233991 | EPI_ISL_4233993 | EPI_ISL_4339335 |
| EPI_ISL_4236342 | EPI_ISL_4233998 | EPI_ISL_4318002 |
| EPI_ISL_4234000 | EPI_ISL_4236351 | EPI_ISL_4318014 |

| | | |
|---|---|---|
| EPI_ISL_4317973 | EPI_ISL_4317985 | EPI_ISL_4791058 |
| EPI_ISL_4339331 | EPI_ISL_4487212 | EPI_ISL_728846 |
| EPI_ISL_4317988 | EPI_ISL_4487209 | EPI_ISL_693862 |
| EPI_ISL_4339333 | EPI_ISL_4487206 | EPI_ISL_693867 |
| EPI_ISL_4316077 | EPI_ISL_4487215 | EPI_ISL_693870 |
| EPI_ISL_4317976 | EPI_ISL_4581208 | EPI_ISL_693872 |
| EPI_ISL_4318008 | EPI_ISL_4588371 | EPI_ISL_693873 |
| EPI_ISL_4317996 | EPI_ISL_4631455 | EPI_ISL_693888 |
| EPI_ISL_4318018 | EPI_ISL_4631453 | EPI_ISL_1005181 |
| EPI_ISL_4318034 | EPI_ISL_4631451 | EPI_ISL_1005170 |
| EPI_ISL_4317979 | EPI_ISL_4631458 | EPI_ISL_1005142 |
| EPI_ISL_4317982 | EPI_ISL_4631456 | EPI_ISL_1005080 |
| EPI_ISL_4318011 | EPI_ISL_4631459 | EPI_ISL_1130039 |
| EPI_ISL_4316079 | EPI_ISL_4631452 | EPI_ISL_693939 |
| EPI_ISL_4318021 | EPI_ISL_4631460 | EPI_ISL_693941 |
| EPI_ISL_4316082 | EPI_ISL_4631457 | EPI_ISL_693942 |
| EPI_ISL_4317993 | EPI_ISL_4631450 | EPI_ISL_693943 |
| EPI_ISL_4317964 | EPI_ISL_4631454 | EPI_ISL_693944 |
| EPI_ISL_4317967 | EPI_ISL_4581201 | EPI_ISL_693945 |

| |
|---|
| EPI_ISL_693946 |
| EPI_ISL_693947 |
| EPI_ISL_694003 |
| EPI_ISL_728973 |
| EPI_ISL_721801 |
| EPI_ISL_721802 |
| EPI_ISL_1130045 |
| EPI_ISL_721684 |
| EPI_ISL_721803 |
| EPI_ISL_721804 |
| EPI_ISL_721805 |
| EPI_ISL_899515 |
| EPI_ISL_729081 |
| EPI_ISL_729130 |
| EPI_ISL_1130037 |
| EPI_ISL_1129997 |
| EPI_ISL_729001 |
| EPI_ISL_728774 |
| EPI_ISL_1130027 |

| |
|---|
| EPI_ISL_1130042 |
| EPI_ISL_737413 |
| EPI_ISL_737431 |
| EPI_ISL_737549 |
| EPI_ISL_1002357 |
| EPI_ISL_1130008 |
| EPI_ISL_768101 |
| EPI_ISL_1260205 |
| EPI_ISL_1260201 |
| EPI_ISL_1260200 |
| EPI_ISL_1259430 |
| EPI_ISL_1260202 |
| EPI_ISL_1259558 |
| EPI_ISL_1260195 |
| EPI_ISL_1260186 |
| EPI_ISL_1130055 |
| EPI_ISL_768100 |
| EPI_ISL_768102 |
| EPI_ISL_768104 |

| |
|---|
| EPI_ISL_1130030 |
| EPI_ISL_1130009 |
| EPI_ISL_768103 |
| EPI_ISL_796434 |
| EPI_ISL_796258 |
| EPI_ISL_796432 |
| EPI_ISL_796430 |
| EPI_ISL_796274 |
| EPI_ISL_1407052 |
| EPI_ISL_796433 |
| EPI_ISL_796429 |
| EPI_ISL_796187 |
| EPI_ISL_796238 |
| EPI_ISL_1130005 |
| EPI_ISL_1129996 |
| EPI_ISL_1130018 |
| EPI_ISL_796270 |
| EPI_ISL_1406900 |
| EPI_ISL_1130024 |

| | | |
|---|---|---|
| EPI_ISL_796198 | EPI_ISL_1004938 | EPI_ISL_1004437 |
| EPI_ISL_796431 | EPI_ISL_1407053 | EPI_ISL_1130052 |
| EPI_ISL_1130003 | EPI_ISL_1004762 | EPI_ISL_1406906 |
| EPI_ISL_796263 | EPI_ISL_1004761 | EPI_ISL_1004438 |
| EPI_ISL_796276 | EPI_ISL_1004753 | EPI_ISL_1004358 |
| EPI_ISL_1130058 | EPI_ISL_899524 | EPI_ISL_1004359 |
| EPI_ISL_899516 | EPI_ISL_899525 | EPI_ISL_1004326 |
| EPI_ISL_1130012 | EPI_ISL_899526 | EPI_ISL_1004361 |
| EPI_ISL_899517 | EPI_ISL_899527 | EPI_ISL_1004360 |
| EPI_ISL_899518 | EPI_ISL_899528 | EPI_ISL_1004357 |
| EPI_ISL_899519 | EPI_ISL_1130032 | EPI_ISL_1259544 |
| EPI_ISL_899520 | EPI_ISL_1130049 | EPI_ISL_1004253 |
| EPI_ISL_899521 | EPI_ISL_899529 | EPI_ISL_1004278 |
| EPI_ISL_899066 | EPI_ISL_1406868 | EPI_ISL_1004277 |
| EPI_ISL_899522 | EPI_ISL_1004669 | EPI_ISL_1004276 |
| EPI_ISL_899523 | EPI_ISL_1004668 | EPI_ISL_1004258 |
| EPI_ISL_1004940 | EPI_ISL_1004598 | EPI_ISL_1004245 |
| EPI_ISL_1130021 | EPI_ISL_1004573 | EPI_ISL_1406937 |
| EPI_ISL_1004939 | EPI_ISL_1004473 | EPI_ISL_899530 |

| |
|---|
| EPI_ISL_899531 |
| EPI_ISL_1002217 |
| EPI_ISL_1001702 |
| EPI_ISL_1002493 |
| EPI_ISL_1002627 |
| EPI_ISL_1002626 |
| EPI_ISL_1002629 |
| EPI_ISL_1002630 |
| EPI_ISL_1002628 |
| EPI_ISL_1002685 |
| EPI_ISL_1002807 |
| EPI_ISL_1002805 |
| EPI_ISL_1130015 |
| EPI_ISL_1130000 |
| EPI_ISL_1002806 |
| EPI_ISL_1002809 |
| EPI_ISL_1002804 |
| EPI_ISL_1002810 |
| EPI_ISL_1002802 |

| |
|---|
| EPI_ISL_1002808 |
| EPI_ISL_1002803 |
| EPI_ISL_1002858 |
| EPI_ISL_1002859 |
| EPI_ISL_1002899 |
| EPI_ISL_1002895 |
| EPI_ISL_1002898 |
| EPI_ISL_1002897 |
| EPI_ISL_1002896 |
| EPI_ISL_1002900 |
| EPI_ISL_899532 |
| EPI_ISL_899533 |
| EPI_ISL_899534 |
| EPI_ISL_1059552 |
| EPI_ISL_899041 |
| EPI_ISL_1059319 |
| EPI_ISL_1059560 |
| EPI_ISL_899535 |
| EPI_ISL_899536 |

| |
|---|
| EPI_ISL_1002273 |
| EPI_ISL_1003251 |
| EPI_ISL_1003249 |
| EPI_ISL_1003250 |
| EPI_ISL_1001986 |
| EPI_ISL_1130011 |
| EPI_ISL_1130057 |
| EPI_ISL_1001949 |
| EPI_ISL_1002009 |
| EPI_ISL_1002037 |
| EPI_ISL_1003058 |
| EPI_ISL_1003106 |
| EPI_ISL_1003313 |
| EPI_ISL_1259496 |
| EPI_ISL_1059559 |
| EPI_ISL_1130026 |
| EPI_ISL_1003164 |
| EPI_ISL_1001516 |
| EPI_ISL_1130043 |

| |
|---|
| EPI_ISL_1406952 |
| EPI_ISL_1002087 |
| EPI_ISL_1129999 |
| EPI_ISL_1003658 |
| EPI_ISL_1003493 |
| EPI_ISL_1003558 |
| EPI_ISL_1130010 |
| EPI_ISL_1003559 |
| EPI_ISL_1130029 |
| EPI_ISL_1001497 |
| EPI_ISL_1130002 |
| EPI_ISL_1003789 |
| EPI_ISL_1130056 |
| EPI_ISL_1130059 |
| EPI_ISL_1130023 |
| EPI_ISL_1059359 |
| EPI_ISL_1059555 |
| EPI_ISL_1059554 |
| EPI_ISL_1059556 |

| |
|---|
| EPI_ISL_1059550 |
| EPI_ISL_1059393 |
| EPI_ISL_1130025 |
| EPI_ISL_1001500 |
| EPI_ISL_1002319 |
| EPI_ISL_1003909 |
| EPI_ISL_1001616 |
| EPI_ISL_1130038 |
| EPI_ISL_1130022 |
| EPI_ISL_1130047 |
| EPI_ISL_1130048 |
| EPI_ISL_1130051 |
| EPI_ISL_1130046 |
| EPI_ISL_1004060 |
| EPI_ISL_1130041 |
| EPI_ISL_1130035 |
| EPI_ISL_1130054 |
| EPI_ISL_1130031 |
| EPI_ISL_1129994 |

| |
|---|
| EPI_ISL_1002428 |
| EPI_ISL_1002425 |
| EPI_ISL_1002424 |
| EPI_ISL_1002427 |
| EPI_ISL_1002426 |
| EPI_ISL_1002421 |
| EPI_ISL_1002422 |
| EPI_ISL_1002423 |
| EPI_ISL_1002248 |
| EPI_ISL_1130007 |
| EPI_ISL_1129998 |
| EPI_ISL_1130019 |
| EPI_ISL_1059553 |
| EPI_ISL_1059551 |
| EPI_ISL_1059434 |
| EPI_ISL_1059433 |
| EPI_ISL_1059557 |
| EPI_ISL_1130028 |
| EPI_ISL_1130014 |

| | | |
|---|---|---|
| EPI_ISL_1130033 | EPI_ISL_1119911 | EPI_ISL_1119266 |
| EPI_ISL_1130044 | EPI_ISL_1119225 | EPI_ISL_1119081 |
| EPI_ISL_1130004 | EPI_ISL_1119226 | EPI_ISL_1119389 |
| EPI_ISL_1130053 | EPI_ISL_1119360 | EPI_ISL_1120023 |
| EPI_ISL_1129995 | EPI_ISL_1119130 | EPI_ISL_1120065 |
| EPI_ISL_1130034 | EPI_ISL_1119398 | EPI_ISL_1119493 |
| EPI_ISL_1130036 | EPI_ISL_1118961 | EPI_ISL_1119573 |
| EPI_ISL_1130020 | EPI_ISL_1119216 | EPI_ISL_1119572 |
| EPI_ISL_1130050 | EPI_ISL_1119243 | EPI_ISL_1119665 |
| EPI_ISL_1130006 | EPI_ISL_1119242 | EPI_ISL_1119664 |
| EPI_ISL_1130017 | EPI_ISL_1119299 | EPI_ISL_1119381 |
| EPI_ISL_1059417 | EPI_ISL_1119199 | EPI_ISL_1119829 |
| EPI_ISL_1080515 | EPI_ISL_1119220 | EPI_ISL_1119900 |
| EPI_ISL_1080516 | EPI_ISL_1119325 | EPI_ISL_1194992 |
| EPI_ISL_1080500 | EPI_ISL_1119036 | EPI_ISL_1194995 |
| EPI_ISL_1080525 | EPI_ISL_1119038 | EPI_ISL_1194991 |
| EPI_ISL_1059558 | EPI_ISL_1119037 | EPI_ISL_1194993 |
| EPI_ISL_1059412 | EPI_ISL_1119454 | EPI_ISL_1194994 |
| EPI_ISL_1080524 | EPI_ISL_1119197 | EPI_ISL_1260189 |

| |
|---|
| EPI_ISL_1260188 |
| EPI_ISL_1260191 |
| EPI_ISL_1260197 |
| EPI_ISL_1260190 |
| EPI_ISL_1259794 |
| EPI_ISL_1259423 |
| EPI_ISL_1259630 |
| EPI_ISL_1260194 |
| EPI_ISL_1259417 |
| EPI_ISL_1260193 |
| EPI_ISL_1260199 |
| EPI_ISL_1260203 |
| EPI_ISL_1260192 |
| EPI_ISL_1259454 |
| EPI_ISL_1259631 |
| EPI_ISL_1408675 |
| EPI_ISL_1654180 |
| EPI_ISL_1408366 |
| EPI_ISL_1408674 |

| |
|---|
| EPI_ISL_1654041 |
| EPI_ISL_1408672 |
| EPI_ISL_1408386 |
| EPI_ISL_1654194 |
| EPI_ISL_1408671 |
| EPI_ISL_1408678 |
| EPI_ISL_1408673 |
| EPI_ISL_1407549 |
| EPI_ISL_1407481 |
| EPI_ISL_1408670 |
| EPI_ISL_1407929 |
| EPI_ISL_1408144 |
| EPI_ISL_1408142 |
| EPI_ISL_1407930 |
| EPI_ISL_1408139 |
| EPI_ISL_1407528 |
| EPI_ISL_1407529 |
| EPI_ISL_1407616 |
| EPI_ISL_1407729 |

| |
|---|
| EPI_ISL_1407928 |
| EPI_ISL_1259470 |
| EPI_ISL_1259471 |
| EPI_ISL_1259747 |
| EPI_ISL_1260204 |
| EPI_ISL_1260196 |
| EPI_ISL_1260187 |
| EPI_ISL_1260206 |
| EPI_ISL_1360859 |
| EPI_ISL_1260198 |
| EPI_ISL_1360845 |
| EPI_ISL_1360863 |
| EPI_ISL_1360848 |
| EPI_ISL_1360838 |
| EPI_ISL_1360858 |
| EPI_ISL_1360861 |
| EPI_ISL_1360846 |
| EPI_ISL_1360850 |
| EPI_ISL_1760929 |

| |
|---|
| EPI_ISL_1760915 |
| EPI_ISL_1760926 |
| EPI_ISL_1760918 |
| EPI_ISL_1760931 |
| EPI_ISL_1760928 |
| EPI_ISL_1760920 |
| EPI_ISL_1760916 |
| EPI_ISL_1760925 |
| EPI_ISL_1760927 |
| EPI_ISL_1760923 |
| EPI_ISL_1760924 |
| EPI_ISL_1760930 |
| EPI_ISL_1760932 |
| EPI_ISL_1760917 |
| EPI_ISL_1760921 |
| EPI_ISL_1360866 |
| EPI_ISL_1360865 |
| EPI_ISL_1360841 |
| EPI_ISL_1360853 |

| |
|---|
| EPI_ISL_1760922 |
| EPI_ISL_1360839 |
| EPI_ISL_1360847 |
| EPI_ISL_1360844 |
| EPI_ISL_1760919 |
| EPI_ISL_1408669 |
| EPI_ISL_1360856 |
| EPI_ISL_1360867 |
| EPI_ISL_1408668 |
| EPI_ISL_1408677 |
| EPI_ISL_1408676 |
| EPI_ISL_1360843 |
| EPI_ISL_1360860 |
| EPI_ISL_1360862 |
| EPI_ISL_1360840 |
| EPI_ISL_1360842 |
| EPI_ISL_1360854 |
| EPI_ISL_1360864 |
| EPI_ISL_1360849 |

| |
|---|
| EPI_ISL_1360851 |
| EPI_ISL_1360852 |
| EPI_ISL_1360855 |
| EPI_ISL_1360857 |
| EPI_ISL_1407927 |
| EPI_ISL_1408143 |
| EPI_ISL_1407781 |
| EPI_ISL_1407480 |
| EPI_ISL_1408371 |
| EPI_ISL_1407923 |
| EPI_ISL_1407931 |
| EPI_ISL_1407924 |
| EPI_ISL_1407932 |
| EPI_ISL_1407925 |
| EPI_ISL_1407755 |
| EPI_ISL_1408263 |
| EPI_ISL_1407503 |
| EPI_ISL_1407558 |
| EPI_ISL_1654145 |

| |
|---|
| EPI_ISL_1408356 |
| EPI_ISL_1654105 |
| EPI_ISL_1408341 |
| EPI_ISL_1654037 |
| EPI_ISL_1408404 |
| EPI_ISL_1654060 |
| EPI_ISL_1654166 |
| EPI_ISL_1407926 |
| EPI_ISL_1496519 |
| EPI_ISL_1496241 |
| EPI_ISL_1407499 |
| EPI_ISL_1407933 |
| EPI_ISL_1408141 |
| EPI_ISL_1408140 |
| EPI_ISL_1598351 |
| EPI_ISL_1598792 |
| EPI_ISL_1598793 |
| EPI_ISL_1598329 |
| EPI_ISL_1598791 |

| |
|---|
| EPI_ISL_1598341 |
| EPI_ISL_1598338 |
| EPI_ISL_1598344 |
| EPI_ISL_1598346 |
| EPI_ISL_1598637 |
| EPI_ISL_1598349 |
| EPI_ISL_1598772 |
| EPI_ISL_1598110 |
| EPI_ISL_1598331 |
| EPI_ISL_1598337 |
| EPI_ISL_1598354 |
| EPI_ISL_1598985 |
| EPI_ISL_1598986 |
| EPI_ISL_1598980 |
| EPI_ISL_1598977 |
| EPI_ISL_1598978 |
| EPI_ISL_1598979 |
| EPI_ISL_1598981 |
| EPI_ISL_1598613 |

| |
|---|
| EPI_ISL_1598054 |
| EPI_ISL_1598983 |
| EPI_ISL_1598069 |
| EPI_ISL_1598094 |
| EPI_ISL_1597875 |
| EPI_ISL_1598072 |
| EPI_ISL_1598332 |
| EPI_ISL_1598343 |
| EPI_ISL_1597876 |
| EPI_ISL_1598084 |
| EPI_ISL_1597944 |
| EPI_ISL_1598350 |
| EPI_ISL_1598347 |
| EPI_ISL_1598335 |
| EPI_ISL_1597964 |
| EPI_ISL_1598334 |
| EPI_ISL_1598685 |
| EPI_ISL_1597986 |
| EPI_ISL_1598984 |

| |
|---|
| EPI_ISL_1496036 |
| EPI_ISL_1496300 |
| EPI_ISL_1496267 |
| EPI_ISL_1496124 |
| EPI_ISL_1658369 |
| EPI_ISL_1658370 |
| EPI_ISL_1658371 |
| EPI_ISL_1658368 |
| EPI_ISL_2375198 |
| EPI_ISL_1658185 |
| EPI_ISL_1657950 |
| EPI_ISL_1657960 |
| EPI_ISL_1657962 |
| EPI_ISL_1657951 |
| EPI_ISL_1658241 |
| EPI_ISL_1657961 |
| EPI_ISL_1657958 |
| EPI_ISL_1658372 |
| EPI_ISL_1657948 |

| |
|---|
| EPI_ISL_1657955 |
| EPI_ISL_1657959 |
| EPI_ISL_1657963 |
| EPI_ISL_1682589 |
| EPI_ISL_1682586 |
| EPI_ISL_1682594 |
| EPI_ISL_1682601 |
| EPI_ISL_1682600 |
| EPI_ISL_1682585 |
| EPI_ISL_1682598 |
| EPI_ISL_1682597 |
| EPI_ISL_1682595 |
| EPI_ISL_1682592 |
| EPI_ISL_1682591 |
| EPI_ISL_1682588 |
| EPI_ISL_1598339 |
| EPI_ISL_1598636 |
| EPI_ISL_1598345 |
| EPI_ISL_1598333 |

| |
|---|
| EPI_ISL_1598799 |
| EPI_ISL_1598330 |
| EPI_ISL_1598336 |
| EPI_ISL_1598342 |
| EPI_ISL_1598638 |
| EPI_ISL_1598352 |
| EPI_ISL_1598340 |
| EPI_ISL_1598759 |
| EPI_ISL_1598348 |
| EPI_ISL_1598982 |
| EPI_ISL_1598353 |
| EPI_ISL_1914271 |
| EPI_ISL_1657956 |
| EPI_ISL_1658297 |
| EPI_ISL_1657949 |
| EPI_ISL_1658214 |
| EPI_ISL_1657953 |
| EPI_ISL_1657957 |
| EPI_ISL_1658235 |

| |
|---|
| EPI_ISL_1657954 |
| EPI_ISL_1657952 |
| EPI_ISL_1658512 |
| EPI_ISL_1659338 |
| EPI_ISL_1658458 |
| EPI_ISL_1658523 |
| EPI_ISL_1658469 |
| EPI_ISL_1658485 |
| EPI_ISL_1658652 |
| EPI_ISL_1658539 |
| EPI_ISL_1658856 |
| EPI_ISL_1658852 |
| EPI_ISL_1658862 |
| EPI_ISL_1658854 |
| EPI_ISL_1658866 |
| EPI_ISL_1658865 |
| EPI_ISL_1658554 |
| EPI_ISL_1658861 |
| EPI_ISL_1658853 |

| |
|---|
| EPI_ISL_1658857 |
| EPI_ISL_1658533 |
| EPI_ISL_1659339 |
| EPI_ISL_1659336 |
| EPI_ISL_1659337 |
| EPI_ISL_1659340 |
| EPI_ISL_1659155 |
| EPI_ISL_1659335 |
| EPI_ISL_1658851 |
| EPI_ISL_1658860 |
| EPI_ISL_1658850 |
| EPI_ISL_1658863 |
| EPI_ISL_1658859 |
| EPI_ISL_1658858 |
| EPI_ISL_1658855 |
| EPI_ISL_1658867 |
| EPI_ISL_1658864 |
| EPI_ISL_1750037 |
| EPI_ISL_1750029 |

| |
|---|
| EPI_ISL_1750040 |
| EPI_ISL_1750580 |
| EPI_ISL_1750028 |
| EPI_ISL_1749692 |
| EPI_ISL_1750025 |
| EPI_ISL_1750043 |
| EPI_ISL_1749794 |
| EPI_ISL_1750875 |
| EPI_ISL_1750770 |
| EPI_ISL_1750886 |
| EPI_ISL_1749783 |
| EPI_ISL_1750826 |
| EPI_ISL_1749750 |
| EPI_ISL_1750621 |
| EPI_ISL_1750027 |
| EPI_ISL_1750515 |
| EPI_ISL_1749974 |
| EPI_ISL_1750513 |
| EPI_ISL_1749903 |

| |
|---|
| EPI_ISL_1750569 |
| EPI_ISL_1750038 |
| EPI_ISL_1750030 |
| EPI_ISL_1750042 |
| EPI_ISL_1750419 |
| EPI_ISL_1750026 |
| EPI_ISL_1913595 |
| EPI_ISL_1750420 |
| EPI_ISL_1750516 |
| EPI_ISL_1750032 |
| EPI_ISL_1750033 |
| EPI_ISL_1750034 |
| EPI_ISL_1750041 |
| EPI_ISL_1750035 |
| EPI_ISL_1750677 |
| EPI_ISL_1913605 |
| EPI_ISL_1913601 |
| EPI_ISL_1914162 |
| EPI_ISL_1913613 |

| |
|---|
| EPI_ISL_1913616 |
| EPI_ISL_1913271 |
| EPI_ISL_1913596 |
| EPI_ISL_1913608 |
| EPI_ISL_1913619 |
| EPI_ISL_1913606 |
| EPI_ISL_1913618 |
| EPI_ISL_1913958 |
| EPI_ISL_1913918 |
| EPI_ISL_1913730 |
| EPI_ISL_1913630 |
| EPI_ISL_1914101 |
| EPI_ISL_1913870 |
| EPI_ISL_1913291 |
| EPI_ISL_1914045 |
| EPI_ISL_1913783 |
| EPI_ISL_1913335 |
| EPI_ISL_1913294 |
| EPI_ISL_1914052 |

| |
|---|
| EPI_ISL_1913311 |
| EPI_ISL_1914142 |
| EPI_ISL_1913784 |
| EPI_ISL_1913825 |
| EPI_ISL_1913740 |
| EPI_ISL_1913437 |
| EPI_ISL_1913637 |
| EPI_ISL_1913624 |
| EPI_ISL_1913612 |
| EPI_ISL_1914224 |
| EPI_ISL_1913611 |
| EPI_ISL_1913319 |
| EPI_ISL_1913593 |
| EPI_ISL_1913975 |
| EPI_ISL_1914238 |
| EPI_ISL_1913610 |
| EPI_ISL_1914134 |
| EPI_ISL_1914135 |
| EPI_ISL_1913594 |

| |
|---|
| EPI_ISL_1913625 |
| EPI_ISL_1913974 |
| EPI_ISL_1913623 |
| EPI_ISL_1913341 |
| EPI_ISL_1913970 |
| EPI_ISL_1913622 |
| EPI_ISL_1913614 |
| EPI_ISL_1913589 |
| EPI_ISL_1913301 |
| EPI_ISL_1913602 |
| EPI_ISL_1913285 |
| EPI_ISL_1913591 |
| EPI_ISL_1913971 |
| EPI_ISL_1913331 |
| EPI_ISL_1913320 |
| EPI_ISL_1913603 |
| EPI_ISL_1913604 |
| EPI_ISL_1914165 |
| EPI_ISL_1913609 |

| |
|---|
| EPI_ISL_2017012 |
| EPI_ISL_2017007 |
| EPI_ISL_2017176 |
| EPI_ISL_2017135 |
| EPI_ISL_2017002 |
| EPI_ISL_2017005 |
| EPI_ISL_2017004 |
| EPI_ISL_2017011 |
| EPI_ISL_2017006 |
| EPI_ISL_2016859 |
| EPI_ISL_2017003 |
| EPI_ISL_2017008 |
| EPI_ISL_2017234 |
| EPI_ISL_2017010 |
| EPI_ISL_2017009 |
| EPI_ISL_2017226 |
| EPI_ISL_2019300 |
| EPI_ISL_2019274 |
| EPI_ISL_2019339 |

| |
|---|
| EPI_ISL_2019122 |
| EPI_ISL_2019109 |
| EPI_ISL_2019240 |
| EPI_ISL_2019006 |
| EPI_ISL_2019310 |
| EPI_ISL_2019456 |
| EPI_ISL_2019267 |
| EPI_ISL_2018995 |
| EPI_ISL_2018968 |
| EPI_ISL_2019164 |
| EPI_ISL_2019404 |
| EPI_ISL_2019428 |
| EPI_ISL_2019493 |
| EPI_ISL_2019528 |
| EPI_ISL_2019068 |
| EPI_ISL_2019136 |
| EPI_ISL_2019377 |
| EPI_ISL_2307687 |
| EPI_ISL_2308067 |

| |
|---|
| EPI_ISL_2307897 |
| EPI_ISL_2307615 |
| EPI_ISL_2019195 |
| EPI_ISL_2307644 |
| EPI_ISL_2308015 |
| EPI_ISL_2307781 |
| EPI_ISL_2307630 |
| EPI_ISL_2019216 |
| EPI_ISL_2019420 |
| EPI_ISL_2019219 |
| EPI_ISL_2211975 |
| EPI_ISL_2212341 |
| EPI_ISL_2212435 |
| EPI_ISL_2211980 |
| EPI_ISL_2212009 |
| EPI_ISL_2212759 |
| EPI_ISL_2212734 |
| EPI_ISL_2212023 |
| EPI_ISL_2211990 |

| |
|---|
| EPI_ISL_2212000 |
| EPI_ISL_2212622 |
| EPI_ISL_2212020 |
| EPI_ISL_2212618 |
| EPI_ISL_2211613 |
| EPI_ISL_2211611 |
| EPI_ISL_2212441 |
| EPI_ISL_2212004 |
| EPI_ISL_2212443 |
| EPI_ISL_2211986 |
| EPI_ISL_2212014 |
| EPI_ISL_2211994 |
| EPI_ISL_2151962 |
| EPI_ISL_2152049 |
| EPI_ISL_2152344 |
| EPI_ISL_2152216 |
| EPI_ISL_2152052 |
| EPI_ISL_2151958 |
| EPI_ISL_2152305 |

| |
|---|
| EPI_ISL_2151944 |
| EPI_ISL_2152002 |
| EPI_ISL_2152356 |
| EPI_ISL_2152078 |
| EPI_ISL_2152226 |
| EPI_ISL_2151945 |
| EPI_ISL_2152171 |
| EPI_ISL_2151930 |
| EPI_ISL_2152209 |
| EPI_ISL_2152195 |
| EPI_ISL_2151960 |
| EPI_ISL_2152062 |
| EPI_ISL_2152249 |
| EPI_ISL_2152114 |
| EPI_ISL_2307737 |
| EPI_ISL_2307740 |
| EPI_ISL_2307734 |
| EPI_ISL_2307744 |
| EPI_ISL_2375486 |

| | | |
|---|---|---|
| EPI_ISL_2307743 | EPI_ISL_2375203 | EPI_ISL_2544390 |
| EPI_ISL_2307745 | EPI_ISL_2375205 | EPI_ISL_2544385 |
| EPI_ISL_2307749 | EPI_ISL_2375211 | EPI_ISL_2544389 |
| EPI_ISL_2307735 | EPI_ISL_2375197 | EPI_ISL_2544387 |
| EPI_ISL_2307727 | EPI_ISL_2375212 | EPI_ISL_2544384 |
| EPI_ISL_2307738 | EPI_ISL_2375204 | EPI_ISL_2544388 |
| EPI_ISL_2307779 | EPI_ISL_2375210 | EPI_ISL_2611071 |
| EPI_ISL_2307544 | EPI_ISL_2375199 | EPI_ISL_2724425 |
| EPI_ISL_2308018 | EPI_ISL_2662355 | EPI_ISL_2662356 |
| EPI_ISL_2307812 | EPI_ISL_2375194 | EPI_ISL_2662353 |
| EPI_ISL_2307953 | EPI_ISL_2375329 | EPI_ISL_2662354 |
| EPI_ISL_2308047 | EPI_ISL_2375129 | EPI_ISL_2820573 |
| EPI_ISL_2307878 | EPI_ISL_2375326 | EPI_ISL_3854240 |
| EPI_ISL_2375206 | EPI_ISL_2375351 | EPI_ISL_3854260 |
| EPI_ISL_2375208 | EPI_ISL_2375108 | EPI_ISL_3854255 |
| EPI_ISL_2375207 | EPI_ISL_2462658 | EPI_ISL_3854258 |
| EPI_ISL_2375214 | EPI_ISL_2462656 | EPI_ISL_4630567 |
| EPI_ISL_2375213 | EPI_ISL_2462659 | EPI_ISL_4634131 |
| EPI_ISL_2375201 | EPI_ISL_2544386 | EPI_ISL_1388991 |

| | | |
|---|---|---|
| EPI_ISL_910325 | EPI_ISL_1388889 | EPI_ISL_527931 |
| EPI_ISL_3344127 | EPI_ISL_1388892 | EPI_ISL_527932 |
| EPI_ISL_3344126 | EPI_ISL_1388238 | EPI_ISL_527933 |
| EPI_ISL_1388564 | EPI_ISL_1389060 | EPI_ISL_527934 |
| EPI_ISL_1388375 | EPI_ISL_1388562 | EPI_ISL_527935 |
| EPI_ISL_1388920 | EPI_ISL_1388124 | EPI_ISL_527938 |
| EPI_ISL_1388858 | EPI_ISL_1388568 | EPI_ISL_527941 |
| EPI_ISL_1388875 | EPI_ISL_1389049 | EPI_ISL_527942 |
| EPI_ISL_1388877 | EPI_ISL_2918756 | EPI_ISL_527943 |
| EPI_ISL_1388202 | EPI_ISL_527916 | EPI_ISL_527944 |
| EPI_ISL_1388854 | EPI_ISL_527920 | EPI_ISL_527951 |
| EPI_ISL_1388856 | EPI_ISL_527922 | EPI_ISL_527952 |
| EPI_ISL_1388881 | EPI_ISL_527923 | EPI_ISL_527953 |
| EPI_ISL_1388228 | EPI_ISL_527924 | EPI_ISL_527954 |
| EPI_ISL_1388251 | EPI_ISL_527925 | EPI_ISL_527955 |
| EPI_ISL_1388193 | EPI_ISL_418281 | EPI_ISL_527956 |
| EPI_ISL_1388883 | EPI_ISL_527927 | EPI_ISL_527957 |
| EPI_ISL_1388885 | EPI_ISL_527928 | EPI_ISL_527959 |
| EPI_ISL_1388887 | EPI_ISL_527930 | EPI_ISL_527961 |

| |
|---|
| EPI_ISL_527962 |
| EPI_ISL_527963 |
| EPI_ISL_527964 |
| EPI_ISL_527965 |
| EPI_ISL_527966 |
| EPI_ISL_527967 |
| EPI_ISL_527968 |
| EPI_ISL_527969 |
| EPI_ISL_527970 |
| EPI_ISL_527972 |
| EPI_ISL_527973 |
| EPI_ISL_527974 |
| EPI_ISL_527975 |
| EPI_ISL_527977 |
| EPI_ISL_527978 |
| EPI_ISL_527979 |
| EPI_ISL_527980 |
| EPI_ISL_527981 |
| EPI_ISL_527982 |

| |
|---|
| EPI_ISL_527983 |
| EPI_ISL_527985 |
| EPI_ISL_527986 |
| EPI_ISL_527987 |
| EPI_ISL_527988 |
| EPI_ISL_527989 |
| EPI_ISL_530122 |
| EPI_ISL_527991 |
| EPI_ISL_527992 |
| EPI_ISL_527995 |
| EPI_ISL_527996 |
| EPI_ISL_528001 |
| EPI_ISL_528003 |
| EPI_ISL_528004 |
| EPI_ISL_528005 |
| EPI_ISL_528007 |
| EPI_ISL_528008 |
| EPI_ISL_528009 |
| EPI_ISL_528010 |

| |
|---|
| EPI_ISL_528014 |
| EPI_ISL_528016 |
| EPI_ISL_528017 |
| EPI_ISL_528018 |
| EPI_ISL_528019 |
| EPI_ISL_528020 |
| EPI_ISL_528021 |
| EPI_ISL_528022 |
| EPI_ISL_528024 |
| EPI_ISL_528026 |
| EPI_ISL_528028 |
| EPI_ISL_528029 |
| EPI_ISL_528031 |
| EPI_ISL_528033 |
| EPI_ISL_528034 |
| EPI_ISL_528035 |
| EPI_ISL_528036 |
| EPI_ISL_528037 |
| EPI_ISL_528039 |

| |
|---|
| EPI_ISL_528041 |
| EPI_ISL_528042 |
| EPI_ISL_528044 |
| EPI_ISL_528046 |
| EPI_ISL_528047 |
| EPI_ISL_528052 |
| EPI_ISL_528054 |
| EPI_ISL_528057 |
| EPI_ISL_528058 |
| EPI_ISL_528059 |
| EPI_ISL_528060 |
| EPI_ISL_528063 |
| EPI_ISL_528066 |
| EPI_ISL_528067 |
| EPI_ISL_528068 |
| EPI_ISL_528069 |
| EPI_ISL_528070 |
| EPI_ISL_528071 |
| EPI_ISL_528074 |

| |
|---|
| EPI_ISL_528075 |
| EPI_ISL_528076 |
| EPI_ISL_528078 |
| EPI_ISL_528080 |
| EPI_ISL_528081 |
| EPI_ISL_528082 |
| EPI_ISL_528083 |
| EPI_ISL_528085 |
| EPI_ISL_528087 |
| EPI_ISL_528089 |
| EPI_ISL_528091 |
| EPI_ISL_528092 |
| EPI_ISL_528093 |
| EPI_ISL_528094 |
| EPI_ISL_528096 |
| EPI_ISL_528097 |
| EPI_ISL_528098 |
| EPI_ISL_528105 |
| EPI_ISL_528111 |

| |
|---|
| EPI_ISL_528112 |
| EPI_ISL_528114 |
| EPI_ISL_528115 |
| EPI_ISL_528116 |
| EPI_ISL_528118 |
| EPI_ISL_528119 |
| EPI_ISL_528120 |
| EPI_ISL_528121 |
| EPI_ISL_528123 |
| EPI_ISL_528124 |
| EPI_ISL_528127 |
| EPI_ISL_528128 |
| EPI_ISL_528129 |
| EPI_ISL_528132 |
| EPI_ISL_528134 |
| EPI_ISL_528136 |
| EPI_ISL_528138 |
| EPI_ISL_528140 |
| EPI_ISL_528141 |

| |
|---|
| EPI_ISL_528142 |
| EPI_ISL_528143 |
| EPI_ISL_528145 |
| EPI_ISL_528148 |
| EPI_ISL_528149 |
| EPI_ISL_528150 |
| EPI_ISL_528151 |
| EPI_ISL_528156 |
| EPI_ISL_528159 |
| EPI_ISL_528160 |
| EPI_ISL_528163 |
| EPI_ISL_528164 |
| EPI_ISL_528165 |
| EPI_ISL_528166 |
| EPI_ISL_528167 |
| EPI_ISL_528168 |
| EPI_ISL_528171 |
| EPI_ISL_528174 |
| EPI_ISL_528178 |

| |
|---|
| EPI_ISL_528181 |
| EPI_ISL_528182 |
| EPI_ISL_528183 |
| EPI_ISL_528185 |
| EPI_ISL_528186 |
| EPI_ISL_528187 |
| EPI_ISL_528189 |
| EPI_ISL_528190 |
| EPI_ISL_528191 |
| EPI_ISL_528194 |
| EPI_ISL_528195 |
| EPI_ISL_528196 |
| EPI_ISL_528198 |
| EPI_ISL_528200 |
| EPI_ISL_528202 |
| EPI_ISL_528204 |
| EPI_ISL_528205 |
| EPI_ISL_528206 |
| EPI_ISL_528207 |

| |
|---|
| EPI_ISL_528209 |
| EPI_ISL_528213 |
| EPI_ISL_528214 |
| EPI_ISL_528216 |
| EPI_ISL_528217 |
| EPI_ISL_528218 |
| EPI_ISL_528220 |
| EPI_ISL_528221 |
| EPI_ISL_528222 |
| EPI_ISL_528223 |
| EPI_ISL_528227 |
| EPI_ISL_528228 |
| EPI_ISL_528229 |
| EPI_ISL_528230 |
| EPI_ISL_528232 |
| EPI_ISL_528233 |
| EPI_ISL_528234 |
| EPI_ISL_528235 |
| EPI_ISL_528236 |

| | | |
|---|---|---|
| EPI_ISL_528237 | EPI_ISL_528256 | EPI_ISL_528278 |
| EPI_ISL_528238 | EPI_ISL_528257 | EPI_ISL_528280 |
| EPI_ISL_528239 | EPI_ISL_528258 | EPI_ISL_528281 |
| EPI_ISL_528240 | EPI_ISL_528259 | EPI_ISL_528282 |
| EPI_ISL_528241 | EPI_ISL_528260 | EPI_ISL_528283 |
| EPI_ISL_528242 | EPI_ISL_528261 | EPI_ISL_528284 |
| EPI_ISL_528243 | EPI_ISL_528262 | EPI_ISL_528285 |
| EPI_ISL_528244 | EPI_ISL_528263 | EPI_ISL_528286 |
| EPI_ISL_528245 | EPI_ISL_528264 | EPI_ISL_528287 |
| EPI_ISL_528247 | EPI_ISL_528268 | EPI_ISL_528288 |
| EPI_ISL_528248 | EPI_ISL_528269 | EPI_ISL_530123 |
| EPI_ISL_528249 | EPI_ISL_528270 | EPI_ISL_528290 |
| EPI_ISL_528250 | EPI_ISL_528271 | EPI_ISL_528291 |
| EPI_ISL_528251 | EPI_ISL_528272 | EPI_ISL_528292 |
| EPI_ISL_528252 | EPI_ISL_528273 | EPI_ISL_528293 |
| EPI_ISL_528253 | EPI_ISL_528274 | EPI_ISL_528294 |
| EPI_ISL_528254 | EPI_ISL_528275 | EPI_ISL_528295 |
| EPI_ISL_581716 | EPI_ISL_528276 | EPI_ISL_528296 |
| EPI_ISL_528255 | EPI_ISL_528277 | EPI_ISL_528297 |

| |
|---|
| EPI_ISL_528298 |
| EPI_ISL_528299 |
| EPI_ISL_528302 |
| EPI_ISL_528303 |
| EPI_ISL_528304 |
| EPI_ISL_528305 |
| EPI_ISL_528307 |
| EPI_ISL_528309 |
| EPI_ISL_528310 |
| EPI_ISL_528311 |
| EPI_ISL_528312 |
| EPI_ISL_528314 |
| EPI_ISL_528315 |
| EPI_ISL_528316 |
| EPI_ISL_528317 |
| EPI_ISL_528318 |
| EPI_ISL_581722 |
| EPI_ISL_528319 |
| EPI_ISL_528320 |

| |
|---|
| EPI_ISL_528323 |
| EPI_ISL_528325 |
| EPI_ISL_528326 |
| EPI_ISL_528327 |
| EPI_ISL_528330 |
| EPI_ISL_528331 |
| EPI_ISL_528332 |
| EPI_ISL_528333 |
| EPI_ISL_528334 |
| EPI_ISL_528335 |
| EPI_ISL_528336 |
| EPI_ISL_528337 |
| EPI_ISL_528339 |
| EPI_ISL_528340 |
| EPI_ISL_528341 |
| EPI_ISL_528342 |
| EPI_ISL_528343 |
| EPI_ISL_528344 |
| EPI_ISL_528345 |

| |
|---|
| EPI_ISL_528346 |
| EPI_ISL_528347 |
| EPI_ISL_528348 |
| EPI_ISL_528349 |
| EPI_ISL_528350 |
| EPI_ISL_528351 |
| EPI_ISL_528354 |
| EPI_ISL_528355 |
| EPI_ISL_528356 |
| EPI_ISL_528357 |
| EPI_ISL_528358 |
| EPI_ISL_528359 |
| EPI_ISL_528360 |
| EPI_ISL_528361 |
| EPI_ISL_528362 |
| EPI_ISL_528363 |
| EPI_ISL_528364 |
| EPI_ISL_528365 |
| EPI_ISL_528366 |

| |
|---|
| EPI_ISL_528367 |
| EPI_ISL_528369 |
| EPI_ISL_528370 |
| EPI_ISL_581736 |
| EPI_ISL_1388662 |
| EPI_ISL_528371 |
| EPI_ISL_1388750 |
| EPI_ISL_528372 |
| EPI_ISL_581728 |
| EPI_ISL_581729 |
| EPI_ISL_581730 |
| EPI_ISL_581731 |
| EPI_ISL_581783 |
| EPI_ISL_581737 |
| EPI_ISL_581732 |
| EPI_ISL_528373 |
| EPI_ISL_1388678 |
| EPI_ISL_581738 |
| EPI_ISL_581739 |

| |
|---|
| EPI_ISL_581740 |
| EPI_ISL_581741 |
| EPI_ISL_1388188 |
| EPI_ISL_581784 |
| EPI_ISL_581742 |
| EPI_ISL_581744 |
| EPI_ISL_581745 |
| EPI_ISL_581747 |
| EPI_ISL_581786 |
| EPI_ISL_581749 |
| EPI_ISL_581750 |
| EPI_ISL_581787 |
| EPI_ISL_581788 |
| EPI_ISL_581751 |
| EPI_ISL_581789 |
| EPI_ISL_581790 |
| EPI_ISL_581753 |
| EPI_ISL_581754 |
| EPI_ISL_581792 |

| |
|---|
| EPI_ISL_581756 |
| EPI_ISL_581757 |
| EPI_ISL_581758 |
| EPI_ISL_581760 |
| EPI_ISL_581793 |
| EPI_ISL_581761 |
| EPI_ISL_581762 |
| EPI_ISL_581763 |
| EPI_ISL_581794 |
| EPI_ISL_581795 |
| EPI_ISL_581796 |
| EPI_ISL_581764 |
| EPI_ISL_581767 |
| EPI_ISL_581769 |
| EPI_ISL_581797 |
| EPI_ISL_581772 |
| EPI_ISL_581773 |
| EPI_ISL_581774 |
| EPI_ISL_581775 |

| |
|---|
| EPI_ISL_581776 |
| EPI_ISL_581777 |
| EPI_ISL_581778 |
| EPI_ISL_581779 |
| EPI_ISL_581781 |
| EPI_ISL_1388751 |
| EPI_ISL_581799 |
| EPI_ISL_581801 |
| EPI_ISL_581803 |
| EPI_ISL_581804 |
| EPI_ISL_581805 |
| EPI_ISL_581806 |
| EPI_ISL_581807 |
| EPI_ISL_581808 |
| EPI_ISL_581809 |
| EPI_ISL_581810 |
| EPI_ISL_581811 |
| EPI_ISL_581812 |
| EPI_ISL_581668 |

| |
|---|
| EPI_ISL_581669 |
| EPI_ISL_581670 |
| EPI_ISL_581814 |
| EPI_ISL_581671 |
| EPI_ISL_581672 |
| EPI_ISL_581673 |
| EPI_ISL_581815 |
| EPI_ISL_581674 |
| EPI_ISL_581816 |
| EPI_ISL_581675 |
| EPI_ISL_581676 |
| EPI_ISL_581677 |
| EPI_ISL_581817 |
| EPI_ISL_581818 |
| EPI_ISL_581819 |
| EPI_ISL_581820 |
| EPI_ISL_581821 |
| EPI_ISL_581822 |
| EPI_ISL_581824 |

| |
|---|
| EPI_ISL_581825 |
| EPI_ISL_581828 |
| EPI_ISL_581829 |
| EPI_ISL_581830 |
| EPI_ISL_581831 |
| EPI_ISL_581832 |
| EPI_ISL_1388753 |
| EPI_ISL_581835 |
| EPI_ISL_581836 |
| EPI_ISL_581837 |
| EPI_ISL_581838 |
| EPI_ISL_581839 |
| EPI_ISL_581840 |
| EPI_ISL_581841 |
| EPI_ISL_581842 |
| EPI_ISL_581843 |
| EPI_ISL_581844 |
| EPI_ISL_581845 |
| EPI_ISL_581848 |

| |
|---|
| EPI_ISL_581849 |
| EPI_ISL_581850 |
| EPI_ISL_581851 |
| EPI_ISL_581852 |
| EPI_ISL_581853 |
| EPI_ISL_581679 |
| EPI_ISL_581854 |
| EPI_ISL_581855 |
| EPI_ISL_581856 |
| EPI_ISL_581857 |
| EPI_ISL_581858 |
| EPI_ISL_581859 |
| EPI_ISL_581860 |
| EPI_ISL_581861 |
| EPI_ISL_581862 |
| EPI_ISL_581863 |
| EPI_ISL_581864 |
| EPI_ISL_581866 |
| EPI_ISL_581867 |

| |
|---|
| EPI_ISL_581868 |
| EPI_ISL_581681 |
| EPI_ISL_581682 |
| EPI_ISL_581683 |
| EPI_ISL_581687 |
| EPI_ISL_581688 |
| EPI_ISL_581690 |
| EPI_ISL_581692 |
| EPI_ISL_581693 |
| EPI_ISL_581694 |
| EPI_ISL_581695 |
| EPI_ISL_581696 |
| EPI_ISL_581698 |
| EPI_ISL_581699 |
| EPI_ISL_581700 |
| EPI_ISL_581702 |
| EPI_ISL_581703 |
| EPI_ISL_581704 |
| EPI_ISL_581705 |

| |
|---|
| EPI_ISL_581706 |
| EPI_ISL_1388748 |
| EPI_ISL_581707 |
| EPI_ISL_581708 |
| EPI_ISL_581709 |
| EPI_ISL_581710 |
| EPI_ISL_581711 |
| EPI_ISL_581713 |
| EPI_ISL_581715 |
| EPI_ISL_581916 |
| EPI_ISL_581917 |
| EPI_ISL_581919 |
| EPI_ISL_581920 |
| EPI_ISL_1388755 |
| EPI_ISL_581921 |
| EPI_ISL_581922 |
| EPI_ISL_581923 |
| EPI_ISL_581925 |
| EPI_ISL_581926 |

| |
|---|
| EPI_ISL_581927 |
| EPI_ISL_581928 |
| EPI_ISL_581874 |
| EPI_ISL_581891 |
| EPI_ISL_581908 |
| EPI_ISL_581892 |
| EPI_ISL_581893 |
| EPI_ISL_581894 |
| EPI_ISL_581895 |
| EPI_ISL_581897 |
| EPI_ISL_581877 |
| EPI_ISL_581898 |
| EPI_ISL_581878 |
| EPI_ISL_581879 |
| EPI_ISL_581880 |
| EPI_ISL_581881 |
| EPI_ISL_581882 |
| EPI_ISL_581899 |
| EPI_ISL_581883 |

| |
|---|
| EPI_ISL_581884 |
| EPI_ISL_581900 |
| EPI_ISL_581885 |
| EPI_ISL_581901 |
| EPI_ISL_581886 |
| EPI_ISL_581902 |
| EPI_ISL_581887 |
| EPI_ISL_581888 |
| EPI_ISL_581889 |
| EPI_ISL_581909 |
| EPI_ISL_581910 |
| EPI_ISL_1388119 |
| EPI_ISL_581911 |
| EPI_ISL_581912 |
| EPI_ISL_581913 |
| EPI_ISL_581930 |
| EPI_ISL_581931 |
| EPI_ISL_581932 |
| EPI_ISL_581933 |

| |
|---|
| EPI_ISL_581934 |
| EPI_ISL_581935 |
| EPI_ISL_581936 |
| EPI_ISL_581937 |
| EPI_ISL_581940 |
| EPI_ISL_581941 |
| EPI_ISL_581942 |
| EPI_ISL_581943 |
| EPI_ISL_581944 |
| EPI_ISL_581945 |
| EPI_ISL_581946 |
| EPI_ISL_581947 |
| EPI_ISL_581948 |
| EPI_ISL_581949 |
| EPI_ISL_581950 |
| EPI_ISL_581951 |
| EPI_ISL_581952 |
| EPI_ISL_581953 |
| EPI_ISL_581954 |

| |
|---|
| EPI_ISL_581955 |
| EPI_ISL_581956 |
| EPI_ISL_581957 |
| EPI_ISL_581958 |
| EPI_ISL_581964 |
| EPI_ISL_581965 |
| EPI_ISL_581966 |
| EPI_ISL_581967 |
| EPI_ISL_581968 |
| EPI_ISL_581969 |
| EPI_ISL_581970 |
| EPI_ISL_581971 |
| EPI_ISL_581972 |
| EPI_ISL_581973 |
| EPI_ISL_581974 |
| EPI_ISL_1388257 |
| EPI_ISL_581975 |
| EPI_ISL_581976 |
| EPI_ISL_581978 |

| |
|---|
| EPI_ISL_581979 |
| EPI_ISL_581980 |
| EPI_ISL_581981 |
| EPI_ISL_581982 |
| EPI_ISL_581983 |
| EPI_ISL_581984 |
| EPI_ISL_581986 |
| EPI_ISL_581987 |
| EPI_ISL_581988 |
| EPI_ISL_581989 |
| EPI_ISL_581990 |
| EPI_ISL_581991 |
| EPI_ISL_581992 |
| EPI_ISL_581993 |
| EPI_ISL_581994 |
| EPI_ISL_581995 |
| EPI_ISL_581996 |
| EPI_ISL_581997 |
| EPI_ISL_581998 |

| |
|---|
| EPI_ISL_581999 |
| EPI_ISL_830985 |
| EPI_ISL_830740 |
| EPI_ISL_830779 |
| EPI_ISL_830780 |
| EPI_ISL_830781 |
| EPI_ISL_830782 |
| EPI_ISL_830783 |
| EPI_ISL_830784 |
| EPI_ISL_830785 |
| EPI_ISL_830786 |
| EPI_ISL_830986 |
| EPI_ISL_830787 |
| EPI_ISL_830788 |
| EPI_ISL_830789 |
| EPI_ISL_830790 |
| EPI_ISL_830791 |
| EPI_ISL_830792 |
| EPI_ISL_830793 |

| |
|---|
| EPI_ISL_830794 |
| EPI_ISL_830795 |
| EPI_ISL_830796 |
| EPI_ISL_830797 |
| EPI_ISL_830798 |
| EPI_ISL_830987 |
| EPI_ISL_830988 |
| EPI_ISL_830799 |
| EPI_ISL_830800 |
| EPI_ISL_830801 |
| EPI_ISL_830802 |
| EPI_ISL_830803 |
| EPI_ISL_830804 |
| EPI_ISL_830805 |
| EPI_ISL_830806 |
| EPI_ISL_830807 |
| EPI_ISL_830808 |
| EPI_ISL_830809 |
| EPI_ISL_830810 |
| EPI_ISL_830811 |
| EPI_ISL_830812 |
| EPI_ISL_830813 |
| EPI_ISL_830989 |
| EPI_ISL_830814 |
| EPI_ISL_830815 |
| EPI_ISL_830816 |
| EPI_ISL_830817 |
| EPI_ISL_830818 |
| EPI_ISL_830819 |
| EPI_ISL_830820 |
| EPI_ISL_830821 |
| EPI_ISL_830822 |
| EPI_ISL_830745 |
| EPI_ISL_830768 |
| EPI_ISL_830771 |
| EPI_ISL_1388116 |
| EPI_ISL_1388757 |
| EPI_ISL_1388759 |
| EPI_ISL_1388761 |
| EPI_ISL_1388765 |
| EPI_ISL_1388767 |
| EPI_ISL_1388769 |
| EPI_ISL_1388295 |
| EPI_ISL_1388771 |
| EPI_ISL_1388320 |
| EPI_ISL_1388773 |
| EPI_ISL_1388285 |
| EPI_ISL_1388300 |
| EPI_ISL_1388774 |
| EPI_ISL_1388776 |
| EPI_ISL_1388699 |
| EPI_ISL_1388778 |
| EPI_ISL_1388780 |
| EPI_ISL_1388310 |
| EPI_ISL_1388781 |
| EPI_ISL_1388783 |
| EPI_ISL_1388226 |

| | | |
|---|---|---|
| EPI_ISL_1388304 | EPI_ISL_1388234 | EPI_ISL_1388249 |
| EPI_ISL_1388787 | EPI_ISL_1388969 | EPI_ISL_1388685 |
| EPI_ISL_1388199 | EPI_ISL_1388235 | EPI_ISL_1388244 |
| EPI_ISL_1388789 | EPI_ISL_1388972 | EPI_ISL_1388928 |
| EPI_ISL_1388791 | EPI_ISL_1388691 | EPI_ISL_1388995 |
| EPI_ISL_1388795 | EPI_ISL_1388976 | EPI_ISL_1388671 |
| EPI_ISL_1388797 | EPI_ISL_1388977 | EPI_ISL_1389016 |
| EPI_ISL_1388799 | EPI_ISL_1388975 | EPI_ISL_1389014 |
| EPI_ISL_1388324 | EPI_ISL_1388973 | EPI_ISL_1388277 |
| EPI_ISL_1388326 | EPI_ISL_1388982 | EPI_ISL_1388935 |
| EPI_ISL_1388232 | EPI_ISL_1388984 | EPI_ISL_1388936 |
| EPI_ISL_1388322 | EPI_ISL_1388980 | EPI_ISL_1388997 |
| EPI_ISL_949186 | EPI_ISL_1388922 | EPI_ISL_1388214 |
| EPI_ISL_949189 | EPI_ISL_1388924 | EPI_ISL_1389006 |
| EPI_ISL_931157 | EPI_ISL_1388986 | EPI_ISL_1388868 |
| EPI_ISL_1388978 | EPI_ISL_1389121 | EPI_ISL_1388869 |
| EPI_ISL_1388110 | EPI_ISL_1388985 | EPI_ISL_1388996 |
| EPI_ISL_1388210 | EPI_ISL_1388247 | EPI_ISL_1388999 |
| EPI_ISL_1389051 | EPI_ISL_1388926 | EPI_ISL_1389011 |

| |
|---|
| EPI_ISL_1388993 |
| EPI_ISL_1388992 |
| EPI_ISL_1389007 |
| EPI_ISL_1388301 |
| EPI_ISL_1389009 |
| EPI_ISL_1389013 |
| EPI_ISL_1388941 |
| EPI_ISL_1388283 |
| EPI_ISL_1388844 |
| EPI_ISL_1388136 |
| EPI_ISL_1388821 |
| EPI_ISL_1388639 |
| EPI_ISL_1388823 |
| EPI_ISL_1388825 |
| EPI_ISL_1388827 |
| EPI_ISL_1388829 |
| EPI_ISL_1388830 |
| EPI_ISL_1388834 |
| EPI_ISL_1388836 |

| |
|---|
| EPI_ISL_1388838 |
| EPI_ISL_1388845 |
| EPI_ISL_1388850 |
| EPI_ISL_1388852 |
| EPI_ISL_1388961 |
| EPI_ISL_1388952 |
| EPI_ISL_1388959 |
| EPI_ISL_1388957 |
| EPI_ISL_1388962 |
| EPI_ISL_1388967 |
| EPI_ISL_1388966 |
| EPI_ISL_1388212 |
| EPI_ISL_1388956 |
| EPI_ISL_1388328 |
| EPI_ISL_1388222 |
| EPI_ISL_1388718 |
| EPI_ISL_1389018 |
| EPI_ISL_1388668 |
| EPI_ISL_1389024 |

| |
|---|
| EPI_ISL_1389030 |
| EPI_ISL_1388128 |
| EPI_ISL_1388642 |
| EPI_ISL_1388944 |
| EPI_ISL_1388950 |
| EPI_ISL_1388726 |
| EPI_ISL_1388958 |
| EPI_ISL_1388955 |
| EPI_ISL_1388963 |
| EPI_ISL_1388242 |
| EPI_ISL_1388126 |
| EPI_ISL_1388195 |
| EPI_ISL_1388954 |
| EPI_ISL_1388968 |
| EPI_ISL_1388953 |
| EPI_ISL_1388330 |
| EPI_ISL_1388137 |
| EPI_ISL_1388631 |
| EPI_ISL_1388629 |

| | | |
|---|---|---|
| EPI_ISL_1388965 | EPI_ISL_1388697 | EPI_ISL_1388358 |
| EPI_ISL_1388332 | EPI_ISL_1388175 | EPI_ISL_1389020 |
| EPI_ISL_1388160 | EPI_ISL_1388633 | EPI_ISL_1388141 |
| EPI_ISL_1388744 | EPI_ISL_1388177 | EPI_ISL_1388742 |
| EPI_ISL_1389123 | EPI_ISL_1388723 | EPI_ISL_1389045 |
| EPI_ISL_1389124 | EPI_ISL_1388733 | EPI_ISL_1388370 |
| EPI_ISL_1388293 | EPI_ISL_1388988 | EPI_ISL_1388154 |
| EPI_ISL_1389129 | EPI_ISL_1388989 | EPI_ISL_1389047 |
| EPI_ISL_1389131 | EPI_ISL_1388132 | EPI_ISL_1389029 |
| EPI_ISL_1388449 | EPI_ISL_1388466 | EPI_ISL_1389033 |
| EPI_ISL_1389133 | EPI_ISL_1388464 | EPI_ISL_1388471 |
| EPI_ISL_1388118 | EPI_ISL_1388652 | EPI_ISL_1388473 |
| EPI_ISL_1388149 | EPI_ISL_1388635 | EPI_ISL_1388183 |
| EPI_ISL_1388722 | EPI_ISL_1389023 | EPI_ISL_1389035 |
| EPI_ISL_1389061 | EPI_ISL_1388469 | EPI_ISL_1388291 |
| EPI_ISL_1388139 | EPI_ISL_1388550 | EPI_ISL_1388366 |
| EPI_ISL_1388179 | EPI_ISL_1389022 | EPI_ISL_1388552 |
| EPI_ISL_1388729 | EPI_ISL_1388460 | EPI_ISL_1388604 |
| EPI_ISL_1388731 | EPI_ISL_1389027 | EPI_ISL_1388740 |

| | | |
|---|---|---|
| EPI_ISL_1389034 | EPI_ISL_1388576 | EPI_ISL_1388683 |
| EPI_ISL_1389037 | EPI_ISL_1388645 | EPI_ISL_1388281 |
| EPI_ISL_1389039 | EPI_ISL_1388388 | EPI_ISL_1389070 |
| EPI_ISL_1388476 | EPI_ISL_1389074 | EPI_ISL_1388306 |
| EPI_ISL_1388474 | EPI_ISL_1388644 | EPI_ISL_1388488 |
| EPI_ISL_1388259 | EPI_ISL_1388344 | EPI_ISL_1388312 |
| EPI_ISL_1389036 | EPI_ISL_1388156 | EPI_ISL_1388574 |
| EPI_ISL_1388532 | EPI_ISL_1388204 | EPI_ISL_1389065 |
| EPI_ISL_1389031 | EPI_ISL_1388390 | EPI_ISL_1388490 |
| EPI_ISL_1389032 | EPI_ISL_1389081 | EPI_ISL_1388267 |
| EPI_ISL_1388679 | EPI_ISL_1388346 | EPI_ISL_1389137 |
| EPI_ISL_1388579 | EPI_ISL_1389078 | EPI_ISL_1388145 |
| EPI_ISL_1389091 | EPI_ISL_1388504 | EPI_ISL_1388518 |
| EPI_ISL_1388501 | EPI_ISL_1389083 | EPI_ISL_1388158 |
| EPI_ISL_1388494 | EPI_ISL_1389087 | EPI_ISL_1389114 |
| EPI_ISL_1388499 | EPI_ISL_1388650 | EPI_ISL_1388318 |
| EPI_ISL_1388580 | EPI_ISL_1388396 | EPI_ISL_1389099 |
| EPI_ISL_1389089 | EPI_ISL_1388394 | EPI_ISL_1388455 |
| EPI_ISL_1388492 | EPI_ISL_1388392 | EPI_ISL_1388406 |

| |
|---|
| EPI_ISL_1388397 |
| EPI_ISL_1388507 |
| EPI_ISL_1388520 |
| EPI_ISL_1388522 |
| EPI_ISL_1388400 |
| EPI_ISL_1388524 |
| EPI_ISL_1388514 |
| EPI_ISL_1388585 |
| EPI_ISL_1388509 |
| EPI_ISL_1388511 |
| EPI_ISL_1388526 |
| EPI_ISL_1389085 |
| EPI_ISL_1388143 |
| EPI_ISL_1388402 |
| EPI_ISL_1388583 |
| EPI_ISL_1389101 |
| EPI_ISL_1388530 |
| EPI_ISL_1389103 |
| EPI_ISL_1388408 |

| |
|---|
| EPI_ISL_1388410 |
| EPI_ISL_1388412 |
| EPI_ISL_1389105 |
| EPI_ISL_1389107 |
| EPI_ISL_1389111 |
| EPI_ISL_1389113 |
| EPI_ISL_1389093 |
| EPI_ISL_1747768 |
| EPI_ISL_1748314 |
| EPI_ISL_1748067 |
| EPI_ISL_1748321 |
| EPI_ISL_1748355 |
| EPI_ISL_1748434 |
| EPI_ISL_1748653 |
| EPI_ISL_1748263 |
| EPI_ISL_1747864 |
| EPI_ISL_2270110 |
| EPI_ISL_2652251 |
| EPI_ISL_2610991 |

| |
|---|
| EPI_ISL_2610970 |
| EPI_ISL_2610992 |
| EPI_ISL_2610993 |
| EPI_ISL_2610977 |
| EPI_ISL_2610995 |
| EPI_ISL_2610996 |
| EPI_ISL_2610997 |
| EPI_ISL_2652252 |
| EPI_ISL_2610998 |
| EPI_ISL_2610999 |
| EPI_ISL_2622137 |
| EPI_ISL_2611001 |
| EPI_ISL_2622139 |
| EPI_ISL_2622140 |
| EPI_ISL_2652253 |
| EPI_ISL_2652254 |
| EPI_ISL_2622141 |
| EPI_ISL_2652255 |
| EPI_ISL_2622143 |

| |
|---|
| EPI_ISL_2622144 |
| EPI_ISL_2622145 |
| EPI_ISL_2622147 |
| EPI_ISL_2622148 |
| EPI_ISL_2622149 |
| EPI_ISL_2622150 |
| EPI_ISL_2652256 |
| EPI_ISL_2652257 |
| EPI_ISL_2652258 |
| EPI_ISL_2652262 |
| EPI_ISL_2652263 |
| EPI_ISL_2652264 |
| EPI_ISL_2768037 |
| EPI_ISL_2768038 |
| EPI_ISL_2768039 |
| EPI_ISL_2768040 |
| EPI_ISL_2768041 |
| EPI_ISL_2768082 |
| EPI_ISL_2768083 |

| |
|---|
| EPI_ISL_2768084 |
| EPI_ISL_2768085 |
| EPI_ISL_2768086 |
| EPI_ISL_2768093 |
| EPI_ISL_2768094 |
| EPI_ISL_2833668 |
| EPI_ISL_2858589 |
| EPI_ISL_2858590 |
| EPI_ISL_3086263 |
| EPI_ISL_3086265 |
| EPI_ISL_3086267 |
| EPI_ISL_3086268 |
| EPI_ISL_3086269 |
| EPI_ISL_3086273 |
| EPI_ISL_3086274 |
| EPI_ISL_3086277 |
| EPI_ISL_3030102 |
| EPI_ISL_3030104 |
| EPI_ISL_3030106 |

| |
|---|
| EPI_ISL_3030109 |
| EPI_ISL_3030111 |
| EPI_ISL_3030113 |
| EPI_ISL_3030117 |
| EPI_ISL_3030119 |
| EPI_ISL_3030123 |
| EPI_ISL_3030127 |
| EPI_ISL_3030129 |
| EPI_ISL_3030131 |
| EPI_ISL_3030133 |
| EPI_ISL_3030135 |
| EPI_ISL_3086280 |
| EPI_ISL_3086282 |
| EPI_ISL_3086284 |
| EPI_ISL_3086285 |
| EPI_ISL_3086287 |
| EPI_ISL_3086289 |
| EPI_ISL_3086290 |
| EPI_ISL_3184341 |

| |
|---|
| EPI_ISL_3184342 |
| EPI_ISL_3184343 |
| EPI_ISL_3184344 |
| EPI_ISL_3184345 |
| EPI_ISL_3184346 |
| EPI_ISL_3184347 |
| EPI_ISL_3086294 |
| EPI_ISL_3086295 |
| EPI_ISL_3086297 |
| EPI_ISL_3086302 |
| EPI_ISL_3086303 |
| EPI_ISL_3086305 |
| EPI_ISL_3086306 |
| EPI_ISL_3086307 |
| EPI_ISL_3247285 |
| EPI_ISL_3247286 |
| EPI_ISL_3247287 |
| EPI_ISL_3247288 |
| EPI_ISL_3247289 |

| |
|---|
| EPI_ISL_3247290 |
| EPI_ISL_3247291 |
| EPI_ISL_3247292 |
| EPI_ISL_3247293 |
| EPI_ISL_3247294 |
| EPI_ISL_3247295 |
| EPI_ISL_3247296 |
| EPI_ISL_3247297 |
| EPI_ISL_3247298 |
| EPI_ISL_3247299 |
| EPI_ISL_3247300 |
| EPI_ISL_3247301 |
| EPI_ISL_3086314 |
| EPI_ISL_3247302 |
| EPI_ISL_3247303 |
| EPI_ISL_3247304 |
| EPI_ISL_3247305 |
| EPI_ISL_3247306 |
| EPI_ISL_3247307 |

| |
|---|
| EPI_ISL_3247308 |
| EPI_ISL_3247309 |
| EPI_ISL_3247310 |
| EPI_ISL_3247313 |
| EPI_ISL_3247314 |
| EPI_ISL_3184348 |
| EPI_ISL_3247315 |
| EPI_ISL_3184349 |
| EPI_ISL_3229188 |
| EPI_ISL_3229189 |
| EPI_ISL_3229190 |
| EPI_ISL_3229191 |
| EPI_ISL_3229192 |
| EPI_ISL_3229193 |
| EPI_ISL_3229194 |
| EPI_ISL_3184351 |
| EPI_ISL_3086316 |
| EPI_ISL_3086317 |
| EPI_ISL_3184352 |

| | | |
|---|---|---|
| EPI_ISL_3184353 | EPI_ISL_3229207 | EPI_ISL_3229231 |
| EPI_ISL_3229195 | EPI_ISL_3229208 | EPI_ISL_3229241 |
| EPI_ISL_3184355 | EPI_ISL_3229209 | EPI_ISL_3229245 |
| EPI_ISL_3184356 | EPI_ISL_3229210 | EPI_ISL_3229246 |
| EPI_ISL_3184367 | EPI_ISL_3229211 | EPI_ISL_3229247 |
| EPI_ISL_3184357 | EPI_ISL_3229213 | EPI_ISL_3229248 |
| EPI_ISL_3184363 | EPI_ISL_3229214 | EPI_ISL_3229249 |
| EPI_ISL_3229196 | EPI_ISL_3229215 | EPI_ISL_3229250 |
| EPI_ISL_3184366 | EPI_ISL_3229217 | EPI_ISL_3229253 |
| EPI_ISL_3229198 | EPI_ISL_3229218 | EPI_ISL_3229254 |
| EPI_ISL_3229199 | EPI_ISL_3229219 | EPI_ISL_3229255 |
| EPI_ISL_3229200 | EPI_ISL_3229220 | EPI_ISL_3303731 |
| EPI_ISL_3229201 | EPI_ISL_3229221 | EPI_ISL_3303732 |
| EPI_ISL_3229202 | EPI_ISL_3229222 | EPI_ISL_3303737 |
| EPI_ISL_3184368 | EPI_ISL_3229223 | EPI_ISL_3303740 |
| EPI_ISL_3184369 | EPI_ISL_3229224 | EPI_ISL_3303743 |
| EPI_ISL_3229204 | EPI_ISL_3229225 | EPI_ISL_3303744 |
| EPI_ISL_3229205 | EPI_ISL_3229227 | EPI_ISL_3303745 |
| EPI_ISL_3229206 | EPI_ISL_3229230 | EPI_ISL_3303746 |

| |
|---|
| EPI_ISL_3303747 |
| EPI_ISL_3303749 |
| EPI_ISL_3303750 |
| EPI_ISL_3303751 |
| EPI_ISL_3344074 |
| EPI_ISL_3344075 |
| EPI_ISL_3247324 |
| EPI_ISL_3247325 |
| EPI_ISL_3247326 |
| EPI_ISL_3247327 |
| EPI_ISL_3247328 |
| EPI_ISL_3247329 |
| EPI_ISL_3247331 |
| EPI_ISL_3303754 |
| EPI_ISL_3303755 |
| EPI_ISL_3303756 |
| EPI_ISL_3303757 |
| EPI_ISL_3247336 |
| EPI_ISL_3247349 |

| |
|---|
| EPI_ISL_3303758 |
| EPI_ISL_3303760 |
| EPI_ISL_3303762 |
| EPI_ISL_3303763 |
| EPI_ISL_3303764 |
| EPI_ISL_3344076 |
| EPI_ISL_3344077 |
| EPI_ISL_3344079 |
| EPI_ISL_3344083 |
| EPI_ISL_3344084 |
| EPI_ISL_3344086 |
| EPI_ISL_3344087 |
| EPI_ISL_3451670 |
| EPI_ISL_3451671 |
| EPI_ISL_3451673 |
| EPI_ISL_3451674 |
| EPI_ISL_3451676 |
| EPI_ISL_3344088 |
| EPI_ISL_3451677 |

| |
|---|
| EPI_ISL_3451678 |
| EPI_ISL_3451679 |
| EPI_ISL_3451680 |
| EPI_ISL_3344091 |
| EPI_ISL_3344092 |
| EPI_ISL_3344093 |
| EPI_ISL_3344094 |
| EPI_ISL_3344095 |
| EPI_ISL_3344096 |
| EPI_ISL_3344097 |
| EPI_ISL_3451681 |
| EPI_ISL_3344098 |
| EPI_ISL_3451682 |
| EPI_ISL_3451683 |
| EPI_ISL_3344100 |
| EPI_ISL_3451684 |
| EPI_ISL_3344101 |
| EPI_ISL_3451685 |
| EPI_ISL_3344102 |

| | | |
|---|---|---|
| EPI_ISL_3344103 | EPI_ISL_3530129 | EPI_ISL_3530171 |
| EPI_ISL_3344104 | EPI_ISL_3530131 | EPI_ISL_3530173 |
| EPI_ISL_3344106 | EPI_ISL_3530133 | EPI_ISL_3530179 |
| EPI_ISL_3344107 | EPI_ISL_3530135 | EPI_ISL_3530181 |
| EPI_ISL_3344109 | EPI_ISL_3530137 | EPI_ISL_4485752 |
| EPI_ISL_3344110 | EPI_ISL_3344122 | EPI_ISL_4251188 |
| EPI_ISL_3344111 | EPI_ISL_3530141 | EPI_ISL_4251189 |
| EPI_ISL_3344112 | EPI_ISL_3530143 | EPI_ISL_4485754 |
| EPI_ISL_3344113 | EPI_ISL_3530144 | EPI_ISL_4251190 |
| EPI_ISL_3344115 | EPI_ISL_3530146 | EPI_ISL_4485755 |
| EPI_ISL_3344116 | EPI_ISL_3530148 | EPI_ISL_4251192 |
| EPI_ISL_3344117 | EPI_ISL_3530150 | EPI_ISL_3451688 |
| EPI_ISL_3344118 | EPI_ISL_3530152 | EPI_ISL_3425797 |
| EPI_ISL_3530118 | EPI_ISL_3530159 | EPI_ISL_3425800 |
| EPI_ISL_3530120 | EPI_ISL_3530161 | EPI_ISL_4485757 |
| EPI_ISL_3530122 | EPI_ISL_3530163 | EPI_ISL_4485759 |
| EPI_ISL_3530124 | EPI_ISL_3530165 | EPI_ISL_4485760 |
| EPI_ISL_3530125 | EPI_ISL_3530167 | EPI_ISL_3710246 |
| EPI_ISL_3530127 | EPI_ISL_3530169 | EPI_ISL_3710247 |

| |
|---|
| EPI_ISL_3710248 |
| EPI_ISL_3710249 |
| EPI_ISL_3710250 |
| EPI_ISL_3425805 |
| EPI_ISL_3710251 |
| EPI_ISL_3710252 |
| EPI_ISL_3425806 |
| EPI_ISL_3710253 |
| EPI_ISL_3530754 |
| EPI_ISL_3530756 |
| EPI_ISL_3530758 |
| EPI_ISL_3451695 |
| EPI_ISL_3451698 |
| EPI_ISL_3530760 |
| EPI_ISL_3530762 |
| EPI_ISL_3530764 |
| EPI_ISL_3530766 |
| EPI_ISL_3530768 |
| EPI_ISL_3303767 |

| |
|---|
| EPI_ISL_3303768 |
| EPI_ISL_3303769 |
| EPI_ISL_3303770 |
| EPI_ISL_3303771 |
| EPI_ISL_3303794 |
| EPI_ISL_3303772 |
| EPI_ISL_3303773 |
| EPI_ISL_3303774 |
| EPI_ISL_3303775 |
| EPI_ISL_3303776 |
| EPI_ISL_3303777 |
| EPI_ISL_3303778 |
| EPI_ISL_3303779 |
| EPI_ISL_3303780 |
| EPI_ISL_3303781 |
| EPI_ISL_3303782 |
| EPI_ISL_3303783 |
| EPI_ISL_3303784 |
| EPI_ISL_3303785 |

| |
|---|
| EPI_ISL_3303786 |
| EPI_ISL_3303787 |
| EPI_ISL_3303788 |
| EPI_ISL_3303789 |
| EPI_ISL_3303790 |
| EPI_ISL_3303791 |
| EPI_ISL_3303792 |
| EPI_ISL_3303793 |
| EPI_ISL_3530770 |
| EPI_ISL_3530772 |
| EPI_ISL_3530774 |
| EPI_ISL_3530776 |
| EPI_ISL_4251194 |
| EPI_ISL_3451719 |
| EPI_ISL_3451723 |
| EPI_ISL_3451732 |
| EPI_ISL_3451733 |
| EPI_ISL_4251197 |
| EPI_ISL_3824224 |

| | | |
|---|---|---|
| EPI_ISL_3530779 | EPI_ISL_3570092 | EPI_ISL_3570110 |
| EPI_ISL_3530783 | EPI_ISL_3570093 | EPI_ISL_3570113 |
| EPI_ISL_3530785 | EPI_ISL_3570094 | EPI_ISL_3570114 |
| EPI_ISL_3530791 | EPI_ISL_3570095 | EPI_ISL_3570115 |
| EPI_ISL_3530792 | EPI_ISL_4392856 | EPI_ISL_3570116 |
| EPI_ISL_3530796 | EPI_ISL_3570098 | EPI_ISL_3570117 |
| EPI_ISL_3530798 | EPI_ISL_3570099 | EPI_ISL_3570118 |
| EPI_ISL_3530800 | EPI_ISL_3710254 | EPI_ISL_3570119 |
| EPI_ISL_3530802 | EPI_ISL_3570100 | EPI_ISL_3570120 |
| EPI_ISL_3530804 | EPI_ISL_3570101 | EPI_ISL_3570121 |
| EPI_ISL_3530806 | EPI_ISL_3570102 | EPI_ISL_3570122 |
| EPI_ISL_3530808 | EPI_ISL_3570103 | EPI_ISL_3570124 |
| EPI_ISL_3570084 | EPI_ISL_3570104 | EPI_ISL_3570125 |
| EPI_ISL_3570085 | EPI_ISL_3570105 | EPI_ISL_3570126 |
| EPI_ISL_3570086 | EPI_ISL_3570106 | EPI_ISL_3570127 |
| EPI_ISL_3570087 | EPI_ISL_3570107 | EPI_ISL_3530829 |
| EPI_ISL_3570088 | EPI_ISL_3570108 | EPI_ISL_3530831 |
| EPI_ISL_3570090 | EPI_ISL_3570109 | EPI_ISL_3570128 |
| EPI_ISL_3570091 | EPI_ISL_3710255 | EPI_ISL_3570129 |

| |
|---|
| EPI_ISL_3570130 |
| EPI_ISL_3570131 |
| EPI_ISL_3570132 |
| EPI_ISL_3570133 |
| EPI_ISL_3570134 |
| EPI_ISL_3570135 |
| EPI_ISL_3570136 |
| EPI_ISL_3570137 |
| EPI_ISL_3570138 |
| EPI_ISL_3570139 |
| EPI_ISL_3570140 |
| EPI_ISL_3570141 |
| EPI_ISL_3570142 |
| EPI_ISL_4392857 |
| EPI_ISL_4392858 |
| EPI_ISL_4392859 |
| EPI_ISL_4392860 |
| EPI_ISL_4392862 |
| EPI_ISL_4392863 |

| |
|---|
| EPI_ISL_4395035 |
| EPI_ISL_3824226 |
| EPI_ISL_4392865 |
| EPI_ISL_3824227 |
| EPI_ISL_4392866 |
| EPI_ISL_4392867 |
| EPI_ISL_4392868 |
| EPI_ISL_4392869 |
| EPI_ISL_4392870 |
| EPI_ISL_4392871 |
| EPI_ISL_3824228 |
| EPI_ISL_3824229 |
| EPI_ISL_4392872 |
| EPI_ISL_4392873 |
| EPI_ISL_4392874 |
| EPI_ISL_4392875 |
| EPI_ISL_4485762 |
| EPI_ISL_4392877 |
| EPI_ISL_3824230 |

| |
|---|
| EPI_ISL_4392878 |
| EPI_ISL_4392879 |
| EPI_ISL_4485763 |
| EPI_ISL_4485765 |
| EPI_ISL_3710264 |
| EPI_ISL_4392880 |
| EPI_ISL_4485766 |
| EPI_ISL_4485768 |
| EPI_ISL_4485770 |
| EPI_ISL_4485771 |
| EPI_ISL_4485773 |
| EPI_ISL_4485776 |
| EPI_ISL_4485778 |
| EPI_ISL_4485781 |
| EPI_ISL_4485783 |
| EPI_ISL_4485784 |
| EPI_ISL_4485786 |
| EPI_ISL_4485788 |
| EPI_ISL_4485789 |

| |
|---|
| EPI_ISL_4485791 |
| EPI_ISL_4485793 |
| EPI_ISL_4485794 |
| EPI_ISL_4485796 |
| EPI_ISL_4251198 |
| EPI_ISL_4251199 |
| EPI_ISL_4251200 |
| EPI_ISL_4251203 |
| EPI_ISL_4485798 |
| EPI_ISL_4251204 |
| EPI_ISL_4251206 |
| EPI_ISL_4485799 |
| EPI_ISL_4485801 |
| EPI_ISL_4485803 |
| EPI_ISL_4485804 |
| EPI_ISL_4485806 |
| EPI_ISL_4485807 |
| EPI_ISL_4485809 |
| EPI_ISL_4485811 |

| |
|---|
| EPI_ISL_4485812 |
| EPI_ISL_4485814 |
| EPI_ISL_4485815 |
| EPI_ISL_4485817 |
| EPI_ISL_4485819 |
| EPI_ISL_4485820 |
| EPI_ISL_4485822 |
| EPI_ISL_4485825 |
| EPI_ISL_4450796 |
| EPI_ISL_3730669 |
| EPI_ISL_3730670 |
| EPI_ISL_3730690 |
| EPI_ISL_3730693 |
| EPI_ISL_3824245 |
| EPI_ISL_3824257 |
| EPI_ISL_3824261 |
| EPI_ISL_3824269 |
| EPI_ISL_4395150 |
| EPI_ISL_3824270 |

| |
|---|
| EPI_ISL_4395085 |
| EPI_ISL_4395151 |
| EPI_ISL_4410796 |
| EPI_ISL_4395152 |
| EPI_ISL_4251219 |
| EPI_ISL_4251221 |
| EPI_ISL_4251223 |
| EPI_ISL_4251231 |
| EPI_ISL_4251235 |
| EPI_ISL_4251236 |
| EPI_ISL_4251237 |
| EPI_ISL_4251238 |
| EPI_ISL_4395153 |
| EPI_ISL_4395154 |
| EPI_ISL_4395155 |
| EPI_ISL_4251248 |
| EPI_ISL_4251249 |
| EPI_ISL_4485827 |
| EPI_ISL_4395156 |

| |
|---|
| EPI_ISL_4395157 |
| EPI_ISL_4395087 |
| EPI_ISL_4395158 |
| EPI_ISL_4395159 |
| EPI_ISL_4395160 |
| EPI_ISL_4395042 |
| EPI_ISL_4395043 |
| EPI_ISL_4395044 |
| EPI_ISL_4395045 |
| EPI_ISL_4395046 |
| EPI_ISL_4395059 |
| EPI_ISL_4395061 |
| EPI_ISL_4395062 |
| EPI_ISL_4395063 |
| EPI_ISL_4395064 |
| EPI_ISL_4395088 |
| EPI_ISL_4395089 |
| EPI_ISL_4395065 |
| EPI_ISL_4395066 |

| |
|---|
| EPI_ISL_4395067 |
| EPI_ISL_4395068 |
| EPI_ISL_4395161 |
| EPI_ISL_4395069 |
| EPI_ISL_4395070 |
| EPI_ISL_4395072 |
| EPI_ISL_4395162 |
| EPI_ISL_4395091 |
| EPI_ISL_4450797 |
| EPI_ISL_4450798 |
| EPI_ISL_4450799 |
| EPI_ISL_4450800 |
| EPI_ISL_4450802 |
| EPI_ISL_4450803 |
| EPI_ISL_4450804 |
| EPI_ISL_4450805 |
| EPI_ISL_4450806 |
| EPI_ISL_4450807 |
| EPI_ISL_4450808 |

| |
|---|
| EPI_ISL_4450809 |
| EPI_ISL_4395102 |
| EPI_ISL_4450810 |
| EPI_ISL_4395103 |
| EPI_ISL_4395104 |
| EPI_ISL_4395105 |
| EPI_ISL_4395107 |
| EPI_ISL_4395109 |
| EPI_ISL_4395110 |
| EPI_ISL_4395113 |
| EPI_ISL_4395114 |
| EPI_ISL_4395138 |
| EPI_ISL_4395163 |
| EPI_ISL_4395164 |
| EPI_ISL_4395165 |
| EPI_ISL_4410804 |
| EPI_ISL_4395166 |
| EPI_ISL_4410806 |
| EPI_ISL_4450855 |

| | | |
|---|---|---|
| EPI_ISL_4395167 | EPI_ISL_4395176 | EPI_ISL_4450891 |
| EPI_ISL_4450859 | EPI_ISL_4395177 | EPI_ISL_4450892 |
| EPI_ISL_4450860 | EPI_ISL_4450873 | EPI_ISL_4450893 |
| EPI_ISL_4450861 | EPI_ISL_4450874 | EPI_ISL_4450894 |
| EPI_ISL_4450862 | EPI_ISL_4450875 | EPI_ISL_4450896 |
| EPI_ISL_4450863 | EPI_ISL_4450876 | EPI_ISL_4450897 |
| EPI_ISL_4395168 | EPI_ISL_4450877 | EPI_ISL_4450829 |
| EPI_ISL_4395169 | EPI_ISL_4450879 | EPI_ISL_4450830 |
| EPI_ISL_4395170 | EPI_ISL_4450880 | EPI_ISL_4450831 |
| EPI_ISL_4395171 | EPI_ISL_4450881 | EPI_ISL_4450832 |
| EPI_ISL_4450867 | EPI_ISL_4450882 | EPI_ISL_4450833 |
| EPI_ISL_4450868 | EPI_ISL_4450883 | EPI_ISL_4450898 |
| EPI_ISL_4450869 | EPI_ISL_4450884 | EPI_ISL_4450834 |
| EPI_ISL_4395172 | EPI_ISL_4450885 | EPI_ISL_4450835 |
| EPI_ISL_4395173 | EPI_ISL_4450887 | EPI_ISL_4450836 |
| EPI_ISL_4395174 | EPI_ISL_4450888 | EPI_ISL_4450900 |
| EPI_ISL_4450870 | EPI_ISL_4450828 | EPI_ISL_4450901 |
| EPI_ISL_4395175 | EPI_ISL_4450889 | EPI_ISL_4450902 |
| EPI_ISL_4450871 | EPI_ISL_4450890 | EPI_ISL_4450903 |

| | | |
|---|---|---|
| EPI_ISL_4450837 | EPI_ISL_4395288 | EPI_ISL_4410833 |
| EPI_ISL_4450838 | EPI_ISL_4395290 | EPI_ISL_4410835 |
| EPI_ISL_4450839 | EPI_ISL_4395292 | EPI_ISL_4410837 |
| EPI_ISL_4450840 | EPI_ISL_4395293 | EPI_ISL_4410838 |
| EPI_ISL_4450841 | EPI_ISL_4395295 | EPI_ISL_4410840 |
| EPI_ISL_4450904 | EPI_ISL_4395297 | EPI_ISL_4410844 |
| EPI_ISL_4450905 | EPI_ISL_4395299 | EPI_ISL_4410845 |
| EPI_ISL_4395229 | EPI_ISL_4395301 | EPI_ISL_4410862 |
| EPI_ISL_4395267 | EPI_ISL_4395303 | EPI_ISL_4410864 |
| EPI_ISL_4395269 | EPI_ISL_4395304 | EPI_ISL_4410866 |
| EPI_ISL_4410809 | EPI_ISL_4395306 | EPI_ISL_4410867 |
| EPI_ISL_4395271 | EPI_ISL_4395308 | EPI_ISL_4410869 |
| EPI_ISL_4395273 | EPI_ISL_4395310 | EPI_ISL_4410871 |
| EPI_ISL_4395275 | EPI_ISL_4395313 | EPI_ISL_4410884 |
| EPI_ISL_4395277 | EPI_ISL_4395315 | EPI_ISL_4410886 |
| EPI_ISL_4395279 | EPI_ISL_4410811 | EPI_ISL_4410888 |
| EPI_ISL_4395281 | EPI_ISL_4410815 | EPI_ISL_4410890 |
| EPI_ISL_4395284 | EPI_ISL_4410816 | EPI_ISL_4542095 |
| EPI_ISL_4395286 | EPI_ISL_4410827 | EPI_ISL_4542097 |

| | | |
|---|---|---|
| EPI_ISL_4542099 | EPI_ISL_4542200 | EPI_ISL_4814050 |
| EPI_ISL_4542102 | EPI_ISL_4542202 | EPI_ISL_4814051 |
| EPI_ISL_4542104 | EPI_ISL_4542205 | EPI_ISL_4814067 |
| EPI_ISL_4542107 | EPI_ISL_4542208 | EPI_ISL_4814069 |
| EPI_ISL_4542110 | EPI_ISL_4542209 | EPI_ISL_4814087 |
| EPI_ISL_4542112 | EPI_ISL_4542212 | EPI_ISL_4814088 |
| EPI_ISL_4542114 | EPI_ISL_4571407 | EPI_ISL_4814097 |
| EPI_ISL_4571399 | EPI_ISL_4571409 | EPI_ISL_4814115 |
| EPI_ISL_4542117 | EPI_ISL_4571410 | EPI_ISL_4814116 |
| EPI_ISL_4542122 | EPI_ISL_4571411 | EPI_ISL_4814121 |
| EPI_ISL_4542155 | EPI_ISL_4571412 | EPI_ISL_4814122 |
| EPI_ISL_4542175 | EPI_ISL_4571420 | EPI_ISL_4814133 |
| EPI_ISL_4542177 | EPI_ISL_4571422 | EPI_ISL_528375 |
| EPI_ISL_4542180 | EPI_ISL_4571423 | EPI_ISL_528376 |
| EPI_ISL_4542185 | EPI_ISL_4571424 | EPI_ISL_528377 |
| EPI_ISL_4542188 | EPI_ISL_4571425 | EPI_ISL_528378 |
| EPI_ISL_4542190 | EPI_ISL_4571427 | EPI_ISL_528381 |
| EPI_ISL_4542194 | EPI_ISL_4814041 | EPI_ISL_581959 |
| EPI_ISL_4542197 | EPI_ISL_4814044 | EPI_ISL_581960 |

| |
|---|
| EPI_ISL_581890 |
| EPI_ISL_581903 |
| EPI_ISL_581904 |
| EPI_ISL_581906 |
| EPI_ISL_581961 |
| EPI_ISL_581963 |
| EPI_ISL_582000 |
| EPI_ISL_582001 |
| EPI_ISL_582002 |
| EPI_ISL_1389057 |
| EPI_ISL_1388983 |
| EPI_ISL_1388971 |
| EPI_ISL_1388218 |
| EPI_ISL_1389010 |
| EPI_ISL_1388279 |
| EPI_ISL_1388681 |
| EPI_ISL_1388840 |
| EPI_ISL_1388987 |
| EPI_ISL_1388727 |

| |
|---|
| EPI_ISL_1388447 |
| EPI_ISL_1388162 |
| EPI_ISL_1388444 |
| EPI_ISL_1388362 |
| EPI_ISL_1388462 |
| EPI_ISL_1388152 |
| EPI_ISL_1388534 |
| EPI_ISL_1388134 |
| EPI_ISL_1388191 |
| EPI_ISL_1388130 |
| EPI_ISL_1388478 |
| EPI_ISL_1388570 |
| EPI_ISL_1388482 |
| EPI_ISL_1388480 |
| EPI_ISL_1388497 |
| EPI_ISL_1388648 |
| EPI_ISL_1388572 |
| EPI_ISL_1388269 |
| EPI_ISL_1388486 |

| |
|---|
| EPI_ISL_1388669 |
| EPI_ISL_1388484 |
| EPI_ISL_1388265 |
| EPI_ISL_1388404 |
| EPI_ISL_1388516 |
| EPI_ISL_1389095 |
| EPI_ISL_2611004 |
| EPI_ISL_2611005 |
| EPI_ISL_2653075 |
| EPI_ISL_3030142 |
| EPI_ISL_3247356 |
| EPI_ISL_3229259 |
| EPI_ISL_3303795 |
| EPI_ISL_3451737 |
| EPI_ISL_3344125 |
| EPI_ISL_3530197 |
| EPI_ISL_3710273 |
| EPI_ISL_3530840 |
| EPI_ISL_3570143 |

| |
|---|
| EPI_ISL_3570144 |
| EPI_ISL_3570148 |
| EPI_ISL_4297849 |
| EPI_ISL_4297850 |
| EPI_ISL_4395084 |
| EPI_ISL_4450850 |
| EPI_ISL_4450851 |
| EPI_ISL_4450852 |
| EPI_ISL_4450916 |
| EPI_ISL_4450921 |
| EPI_ISL_4450922 |

| |
|---|
| EPI_ISL_4450923 |
| EPI_ISL_4450924 |
| EPI_ISL_4395323 |
| EPI_ISL_4395324 |
| EPI_ISL_4410895 |
| EPI_ISL_4410897 |
| EPI_ISL_4542235 |
| EPI_ISL_4542237 |
| EPI_ISL_4814134 |
| EPI_ISL_4814145 |
| EPI_ISL_1388964 |

| |
|---|
| EPI_ISL_1389017 |
| EPI_ISL_1388334 |
| EPI_ISL_2611006 |
| EPI_ISL_2833669 |
| EPI_ISL_2833670 |
| EPI_ISL_3530842 |
| EPI_ISL_4450926 |
| EPI_ISL_1388817 |
| EPI_ISL_1388289 |
| EPI_ISL_1388308 |

**Table S3**. Odds ratios in defined educational institutions (daycare [0-3 yo], kindergarten [4-5 yo], elementary school [6-11 yo], secondary school level I [12-15 yo], secondary school level II [16-19 yo]) compared to adults ( >19 yo) for prevalent lineages from 01.10.2020 to 31.05.2021. Significant results to the level of 0.05 and confidence intervals (CI) not crossing 1 in boldface.

| comparison | OR | 2.5% CI | 97.5% CI | p |
|---|---|---|---|---|
| B.1.1.39_Daycare_vs_adults | 0.000 | NA | 2.00E+13 | 0.9858 |
| B.1.1.39_Kindergarten_vs_adults | 0.000 | NA | 2.40E+24 | 0.9889 |
| B.1.1.39_Elementary_vs_adults | 1.002 | 0.299 | 2.500 | 0.9977 |
| B.1.1.39_secondary_I_vs_adults | 0.946 | 0.283 | 2.356 | 0.9159 |
| B.1.1.39_secondary_II_vs_adults | 1.268 | 0.520 | 2.643 | 0.5615 |
| **B.1.1.7_Daycare_vs_adults** | **4.327** | **1.834** | **11.322** | **0.0013** |
| **B.1.1.7_Kindergarten_vs_adults** | **3.407** | **1.171** | **11.142** | **0.0287** |
| **B.1.1.7_Elementary_vs_adults** | **2.975** | **1.840** | **4.887** | **0.0000** |
| **B.1.1.7_secondary_I_vs_adults** | **1.995** | **1.255** | **3.179** | **0.0035** |
| **B.1.1.7_secondary_II_vs_adults** | **1.521** | **1.008** | **2.283** | **0.0436** |
| B.1.1.70_Daycare_vs_adults | 0.000 | NA | 6.81E+25 | 0.9914 |
| B.1.1.70_Kindergarten_vs_adults | 3.528 | 0.191 | 18.607 | 0.2316 |
| B.1.1.70_Elementary_vs_adults | 0.000 | 0.000 | 1.40E+10 | 0.9847 |
| B.1.1.70_secondary_I_vs_adults | 1.885 | 0.445 | 5.451 | 0.3045 |
| B.1.1.70_secondary_II_vs_adults | 0.459 | 0.026 | 2.174 | 0.4456 |
| B.1.160_Daycare_vs_adults | 0.271 | 0.015 | 1.300 | 0.2028 |
| B.1.160_Kindergarten_vs_adults | 0.000 | NA | 2.22E+05 | 0.9717 |
| B.1.160_Elementary_vs_adults | 0.542 | 0.208 | 1.168 | 0.1571 |
| B.1.160_secondary_I_vs_adults | 0.511 | 0.196 | 1.099 | 0.1201 |
| B.1.160_secondary_II_vs_adults | 0.881 | 0.462 | 1.551 | 0.6782 |
| B.1.177_Daycare_vs_adults | 0.513 | 0.082 | 1.766 | 0.3700 |
| B.1.177_Kindergarten_vs_adults | 0.000 | NA | 2.01E+05 | 0.9715 |
| B.1.177_Elementary_vs_adults | 0.317 | 0.096 | 0.776 | 0.0271 |
| B.1.177_secondary_I_vs_adults | 0.634 | 0.278 | 1.261 | 0.2319 |
| B.1.177_secondary_II_vs_adults | 0.796 | 0.418 | 1.401 | 0.4570 |
| B.1.214.2_Daycare_vs_adults | 4.687 | 0.728 | 17.070 | 0.0432 |
| **B.1.214.2_Kindergarten_vs_adults** | **8.202** | **1.240** | **31.963** | **0.0075** |
| B.1.214.2_Elementary_vs_adults | 1.406 | 0.224 | 4.808 | 0.6460 |
| B.1.214.2_secondary_I_vs_adults | 0.656 | 0.037 | 3.139 | 0.6807 |
| B.1.214.2_secondary_II_vs_adults | 1.507 | 0.356 | 4.348 | 0.5061 |
| B.1.617.2_Daycare_vs_adults | 0.000 | NA | 1.84E+112 | 0.9967 |
| B.1.617.2_Kindergarten_vs_adults | 0.000 | NA | 6.22E+145 | 0.9974 |
| **B.1.617.2_Elementary_vs_adults** | **6.749** | **1.473** | **23.202** | **0.0049** |
| B.1.617.2_secondary_I_vs_adults | 2.070 | 0.112 | 11.225 | 0.4929 |

| | | | | |
|---|---|---|---|---|
| B.1.617.2_secondary_II_vs_adults | 0.000 | NA | 1.12E+50 | 0.9930 |

**Table S4**. Odds ratios in defined educational institutions (daycare [0-3 yo], elementary school [6-11 yo], secondary school level I [12-15 yo], secondary school level II [16-19 yo]) compared to adults ( >19 years old) for prevalent lineages from 01.10.2020 to 31.12.2020. No cases in the age group of kindergarten [4-5 yo]. Significant results to the level of 0.05 and confidence intervals (CI) not crossing 1 in boldface.

| comparison | OR | 2.5% CI | 97.5% CI | p |
|---|---|---|---|---|
| B.1.1.39_Daycare_vs_adults | 0.000 | NA | 3.41E+23 | 0.9836 |
| B.1.1.39_Elementary_vs_adults | 2.491 | 0.692 | 7.164 | 0.1160 |
| B.1.1.39_secondary_I_vs_adults | 1.006 | 0.236 | 2.950 | 0.9924 |
| B.1.1.39_secondary_II_vs_adults | 1.649 | 0.654 | 3.628 | 0.2454 |
| B.1.1.7_Daycare_vs_adults | 0.000 | NA | 1.29E+282 | 0.9984 |
| B.1.1.7_Elementary_vs_adults | 8.059 | 0.409 | 53.675 | 0.0631 |
| B.1.1.7_secondary_I_vs_adults | 0.000 | NA | 6.88E+140 | 0.9962 |
| B.1.1.7_secondary_II_vs_adults | 0.000 | NA | 1.15E+113 | 0.9953 |
| B.1.160_Daycare_vs_adults | 0.765 | 0.039 | 5.213 | 0.8110 |
| B.1.160_Elementary_vs_adults | 1.176 | 0.373 | 3.169 | 0.7607 |
| B.1.160_secondary_I_vs_adults | 0.637 | 0.212 | 1.567 | 0.3670 |
| B.1.160_secondary_II_vs_adults | 1.020 | 0.483 | 2.001 | 0.9569 |
| B.1.177_Daycare_vs_adults | 1.806 | 0.237 | 10.984 | 0.5189 |
| B.1.177_Elementary_vs_adults | 0.339 | 0.053 | 1.205 | 0.1515 |
| B.1.177_secondary_I_vs_adults | 1.032 | 0.423 | 2.285 | 0.9403 |
| B.1.177_secondary_II_vs_adults | 0.903 | 0.428 | 1.770 | 0.7765 |
| **B.1.258_Daycare_vs_adults** | **9.121** | **1.180** | **56.333** | **0.0169** |
| B.1.258_Elementary_vs_adults | 0.805 | 0.044 | 4.053 | 0.8346 |
| B.1.258_secondary_I_vs_adults | 1.013 | 0.160 | 3.526 | 0.9858 |
| B.1.258_secondary_II_vs_adults | 1.001 | 0.236 | 2.890 | 0.9987 |

**Table S5**. Odds ratios in defined educational institutions (daycare [0-3 yo], kindergarten [4-5 yo], elementary school [6-11 yo], secondary school level I [12-15 yo], secondary school level II [16-19 yo]) compared to adults ( >19 years old) for prevalent lineages from 01.01.2021 to 29.02.2021. Significant results to the level of 0.05 and confidence intervals (CI) not crossing 1 in boldface.

| comparison | OR | 2.5% CI | 97.5% CI | p |
|---|---|---|---|---|
| B.1.1.7_Daycare_vs_adults | 1.24E+07 | 0.000 | NA | 0.9800 |
| B.1.1.7_Kindergarten_vs_adults | 1.076 | 0.050 | 11.393 | 0.9526 |
| **B.1.1.7_Elementary_vs_adults** | **3.945** | **1.448** | **11.808** | **0.0090** |
| B.1.1.7_secondary_I_vs_adults | 2.152 | 0.583 | 7.942 | 0.2362 |
| B.1.1.7_secondary_II_vs_adults | 0.478 | 0.072 | 1.909 | 0.3525 |
| B.1.1.70_Daycare_vs_adults | 0.000 | NA | 2.4E+124 | 0.9957 |
| B.1.1.70_Kindergarten_vs_adults | 7.281 | 0.328 | 80.061 | 0.1127 |
| B.1.1.70_Elementary_vs_adults | 0.000 | NA | 3.6E+36 | 0.9920 |
| B.1.1.70_secondary_I_vs_adults | 0.000 | NA | 5.2E+61 | 0.9939 |
| B.1.1.70_secondary_II_vs_adults | 1.456 | 0.077 | 8.350 | 0.7279 |
| B.1.177_Daycare_vs_adults | 0.000 | NA | 8.1E+72 | 0.9929 |
| B.1.177_Kindergarten_vs_adults | 0.000 | NA | 3.4E+122 | 0.9945 |
| B.1.177_Elementary_vs_adults | 0.843 | 0.129 | 3.169 | 0.8258 |
| B.1.177_secondary_I_vs_adults | 0.000 | NA | 9.4E+34 | 0.9900 |
| B.1.177_secondary_II_vs_adults | 1.405 | 0.208 | 5.748 | 0.6719 |
| B.1.214.2_Daycare_vs_adults | 0.000 | NA | 1.3E+207 | 0.9974 |
| B.1.214.2_Kindergarten_vs_adults | 20.250 | 0.873 | 242.850 | 0.0200 |
| B.1.214.2_Elementary_vs_adults | 0.000 | NA | 1.0E+89 | 0.9951 |
| B.1.214.2_secondary_I_vs_adults | 0.000 | NA | 1.4E+118 | 0.9963 |
| **B.1.214.2_secondary_II_vs_adults** | **9.000** | **1.208** | **46.005** | **0.0130** |

**Table S6**. Odds ratios in defined educational institutions (daycare [0-3 yo], kindergarten [4-5 yo], elementary school [6-11 yo], secondary school level I [12-15 yo], secondary school level II [16-19 yo]) compared to adults ( >19 yo) for prevalent lineages from 01.03.2021 to 31.05.2021. Significant results to the level of 0.05 and confidence intervals (CI) not crossing 1 in boldface.

| comparison | OR | 2.5% CI | 97.5% CI | p |
|---|---|---|---|---|
| B.1.1.7_Daycare_vs_adults | 0.889 | 0.232 | 5.8313 | 0.8805 |
| B.1.1.7_Kindergarten_vs_adults | 0.431 | 0.121 | 2.0066 | 0.2228 |
| B.1.1.7_Elementary_vs_adults | 1.035 | 0.422 | 3.1113 | 0.9453 |
| B.1.1.7_secondary_I_vs_adults | 1.833 | 0.635 | 7.7635 | 0.3262 |
| B.1.1.7_secondary_II_vs_adults | 2.318 | 0.814 | 9.7540 | 0.1696 |
| B.1.214.2_Daycare_vs_adults | 3.678 | 0.548 | 14.8086 | 0.1032 |
| B.1.214.2_Kindergarten_vs_adults | 2.023 | 0.108 | 11.2891 | 0.5108 |
| B.1.214.2_Elementary_vs_adults | 1.156 | 0.181 | 4.1482 | 0.8487 |
| B.1.214.2_secondary_I_vs_adults | 0.562 | 0.031 | 2.7992 | 0.5783 |
| B.1.214.2_secondary_II_vs_adults | 0.449 | 0.025 | 2.2201 | 0.4395 |
| B.1.617.2_Daycare_vs_adults | 0.000 | NA | 9.18E+87 | 0.9958 |
| B.1.617.2_Kindergarten_vs_adults | 0.000 | NA | 1.65E+96 | 0.9962 |
| B.1.617.2_Elementary_vs_adults | 4.490 | 0.963 | 15.8680 | 0.0295 |
| B.1.617.2_secondary_I_vs_adults | 1.414 | 0.076 | 7.8355 | 0.7459 |
| B.1.617.2_secondary_II_vs_adults | 0.000 | NA | 3.81E+43 | 0.9921 |

**Table S7**. Summary of SARS-CoV-2 lineages and cases per lineage detected in different educational institutions from 01.10.2020 to 31.05.2021.

| PANGO | Daycare | Kindergarten | Elementary | secondary_I | secondary_II | adults | Total |
|---|---|---|---|---|---|---|---|
| B | | | | | | 1 | 1 |
| B.1 | | | | 2 | 2 | 18 | 22 |
| B.1.1 | | | | 1 | | 11 | 12 |
| B.1.1.1 | | | | | | 4 | 4 |
| B.1.1.117 | | | | | | 1 | 1 |
| B.1.1.130 | | | | | | 1 | 1 |
| B.1.1.135 | | | | | | 1 | 1 |
| B.1.1.141 | | | | | | 1 | 1 |
| B.1.1.144 | | | | | | 1 | 1 |
| B.1.1.153 | | | | | | 4 | 4 |
| B.1.1.170 | | | | | 1 | | 1 |
| B.1.1.186 | | | | | | 1 | 1 |
| B.1.1.189 | | | 2 | | 1 | 6 | 9 |
| B.1.1.194 | | | | | | 1 | 1 |
| B.1.1.220 | | | | | | 1 | 1 |
| B.1.1.247 | | | | | | 1 | 1 |
| B.1.1.250 | | | | | | 2 | 2 |
| B.1.1.269 | | | | | 1 | 1 | 2 |
| B.1.1.277 | | | | | | 2 | 2 |
| B.1.1.288 | | | | | | 6 | 6 |
| B.1.1.29 | | | 1 | | | 9 | 10 |
| B.1.1.297 | | | | | | 2 | 2 |
| B.1.1.318 | | | | | 1 | | 1 |
| B.1.1.348 | | | | | | 1 | 1 |
| B.1.1.37 | | | | | | 2 | 2 |
| B.1.1.39 | | | 4 | 4 | 7 | 78 | 93 |
| B.1.1.445 | | | | | | 1 | 1 |
| B.1.1.47 | | | | 1 | | 5 | 6 |
| B.1.1.519 | | | | | | 2 | 2 |
| B.1.1.54 | | | | | | 1 | 1 |
| B.1.1.7 | 16 | 9 | 44 | 39 | 45 | 486 | 639 |
| B.1.1.70 | | 1 | | 3 | 1 | 30 | 35 |
| B.1.1.84 | | | | | | 2 | 2 |

| | | | | | | | |
|---|---|---|---|---|---|---|---|
| **B.1.1.99** | | | | | 1 | 3 | 4 |
| **B.1.146** | | | | | | 1 | 1 |
| **B.1.160** | 1 | | 6 | 6 | 13 | 202 | 228 |
| **B.1.160.10** | | | | | | 1 | 1 |
| **B.1.160.12** | | | | | | 1 | 1 |
| **B.1.160.14** | | | | | | 1 | 1 |
| **B.1.160.16** | | | | 1 | 1 | 1 | 3 |
| **B.1.160.19** | | | | | | 1 | 1 |
| **B.1.160.20** | | | | | | 11 | 11 |
| **B.1.160.30** | | | | | | 2 | 2 |
| **B.1.160.32** | | | | | | 1 | 1 |
| **B.1.177** | 2 | | 4 | 8 | 13 | 220 | 247 |
| **B.1.177.23** | | | | 1 | 1 | 11 | 13 |
| **B.1.177.43** | | | | | | 2 | 2 |
| **B.1.177.44** | | | | | | 2 | 2 |
| **B.1.177.86** | | | | | | 2 | 2 |
| **B.1.213** | | | | | | 1 | 1 |
| **B.1.214** | | | 1 | | | 4 | 5 |
| **B.1.214.2** | 2 | 2 | 2 | 1 | 3 | 28 | 38 |
| **B.1.221** | | | 1 | 1 | 3 | 47 | 52 |
| **B.1.236** | | | | | | 5 | 5 |
| **B.1.247** | | | | | | 1 | 1 |
| **B.1.258** | 2 | | 1 | 2 | 3 | 59 | 67 |
| **B.1.258.14** | | | | | | 5 | 5 |
| **B.1.258.17** | | | | 1 | 1 | 10 | 12 |
| **B.1.351** | | 2 | | | | 4 | 6 |
| **B.1.36** | | | | | 1 | 10 | 11 |
| **B.1.36.1** | | | 1 | | 1 | 10 | 12 |
| **B.1.36.17** | | | | | | 11 | 11 |
| **B.1.36.35** | | | | | | 6 | 6 |
| **B.1.367** | | | | 1 | | | 1 |
| **B.1.400** | | | 1 | | | | 1 |
| **B.1.416** | | | | | | 2 | 2 |
| **B.1.416.1** | | | | 2 | | 12 | 14 |
| **B.1.474** | | | | | | 1 | 1 |
| **B.1.480** | | | | 1 | | 2 | 3 |

| | | | | | | | |
|---|---|---|---|---|---|---|---|
| **B.1.509** | | | | | | 5 | 5 |
| **B.1.525** | | | | | | 1 | 1 |
| **B.1.617.2** | | | 3 | 1 | | 9 | 13 |
| **B.1.620** | | | | | | 1 | 1 |
| **B.1.8** | | | | | | 1 | 1 |
| **B.1.88** | | | | | 1 | 14 | 15 |
| **B.1.98** | | | | | | 1 | 1 |
| **C.16** | | | | | | 2 | 2 |
| **C.4** | | | | | | 1 | 1 |
| **P.1** | | | | | | 4 | 4 |
| **P.1.2** | | | | | | 2 | 2 |
| **P.2** | | | 1 | | | 2 | 3 |
| **Total** | 23 | 14 | 72 | 76 | 101 | 1406 | 1692 |

**Table S8**. Summary of SARS-CoV-2 lineages and cases per lineage detected in different educational institutions from 01.10.2020 to 31.12.2020.

| | Daycare | Kindergarten | Elementary | secondary_I | secondary_II | adults | Total |
|---|---|---|---|---|---|---|---|
| **B** | | | | | | 1 | 1 |
| **B.1** | | | | 1 | 2 | 12 | 15 |
| **B.1.1** | | | | 1 | | 5 | 6 |
| **B.1.1.1** | | | | | | 4 | 4 |
| **B.1.1.144** | | | | | | 1 | 1 |
| **B.1.1.153** | | | | | | 4 | 4 |
| **B.1.1.170** | | | | | 1 | | 1 |
| **B.1.1.186** | | | | | | 1 | 1 |
| **B.1.1.189** | | | 2 | | 1 | 6 | 9 |
| **B.1.1.194** | | | | | | 1 | 1 |
| **B.1.1.220** | | | | | | 1 | 1 |
| **B.1.1.247** | | | | | | 1 | 1 |
| **B.1.1.250** | | | | | | 1 | 1 |
| **B.1.1.269** | | | | | 1 | 1 | 2 |
| **B.1.1.277** | | | | | | 2 | 2 |
| **B.1.1.288** | | | | | | 5 | 5 |
| **B.1.1.29** | | | | | | 9 | 9 |
| **B.1.1.297** | | | | | | 2 | 2 |

| | | | | | | | |
|---|---|---|---|---|---|---|---|
| **B.1.1.37** | | | | | | 2 | 2 |
| **B.1.1.39** | | | 4 | 3 | 7 | 71 | 85 |
| **B.1.1.47** | | | | | | 4 | 4 |
| **B.1.1.54** | | | | | | 1 | 1 |
| **B.1.1.7** | | | 1 | | | 5 | 6 |
| **B.1.1.70** | | | | 3 | | 13 | 16 |
| **B.1.1.84** | | | | | | 1 | 1 |
| **B.1.1.99** | | | | | 1 | 3 | 4 |
| **B.1.160** | 1 | | 5 | 5 | 11 | 170 | 192 |
| **B.1.160.10** | | | | | | 1 | 1 |
| **B.1.160.12** | | | | | | 1 | 1 |
| **B.1.160.16** | | | | | 1 | | 1 |
| **B.1.160.20** | | | | | | 9 | 9 |
| **B.1.160.30** | | | | | | 1 | 1 |
| **B.1.160.32** | | | | | | 1 | 1 |
| **B.1.177** | 2 | | 2 | 8 | 11 | 186 | 209 |
| **B.1.177.23** | | | | 1 | 1 | 11 | 13 |
| **B.1.177.43** | | | | | | 1 | 1 |
| **B.1.177.44** | | | | | | 1 | 1 |
| **B.1.213** | | | | | | 1 | 1 |
| **B.1.221** | | | 1 | 1 | 2 | 30 | 34 |
| **B.1.236** | | | | | | 4 | 4 |
| **B.1.247** | | | | | | 1 | 1 |
| **B.1.258** | 2 | | 1 | 2 | 3 | 47 | 55 |
| **B.1.258.14** | | | | | | 4 | 4 |
| **B.1.258.17** | | | | | | 1 | 1 |
| **B.1.36** | | | | | | 7 | 7 |
| **B.1.36.1** | | | 1 | | 1 | 7 | 9 |
| **B.1.36.17** | | | | | | 11 | 11 |
| **B.1.367** | | | | 1 | | | 1 |
| **B.1.400** | | | 1 | | | | 1 |
| **B.1.416** | | | | | | 2 | 2 |
| **B.1.416.1** | | | | 2 | | 12 | 14 |
| **B.1.474** | | | | | | 1 | 1 |
| **B.1.480** | | | | 1 | | 2 | 3 |
| **B.1.509** | | | | | | 5 | 5 |

| | | | | | | | |
|---|---|---|---|---|---|---|---|
| **B.1.88** | | | | | 1 | 14 | 15 |
| **B.1.98** | | | | | | 1 | 1 |
| **P.2** | | | | | | 1 | 1 |
| **Total** | 5 | 0 | 18 | 29 | 44 | 690 | 786 |

**Table S9.** Summary of SARS-CoV-2 lineages and cases per lineage detected in different educational institutions from 01.01.2021 to 28.02.2021.

| | Daycare | Kindergarten | Elementary | secondary_I | secondary_II | adults | Total |
|---|---|---|---|---|---|---|---|
| **B.1** | | | | 1 | | 3 | 4 |
| **B.1.1** | | | | | | 2 | 2 |
| **B.1.1.117** | | | | | | 1 | 1 |
| **B.1.1.130** | | | | | | 1 | 1 |
| **B.1.1.135** | | | | | | 1 | 1 |
| **B.1.1.141** | | | | | | 1 | 1 |
| **B.1.1.250** | | | | | | 1 | 1 |
| **B.1.1.288** | | | | | | 1 | 1 |
| **B.1.1.29** | | | 1 | | | | 1 |
| **B.1.1.348** | | | | | | 1 | 1 |
| **B.1.1.39** | | | | 1 | | 6 | 7 |
| **B.1.1.445** | | | | | | 1 | 1 |
| **B.1.1.47** | | | | 1 | | 1 | 2 |
| **B.1.1.7** | 5 | 1 | 11 | 5 | 2 | 79 | 103 |
| **B.1.1.70** | | 1 | | | 1 | 16 | 18 |
| **B.1.1.84** | | | | | | 1 | 1 |
| **B.1.146** | | | | | | 1 | 1 |
| **B.1.160** | | | 1 | 1 | 2 | 31 | 35 |
| **B.1.160.14** | | | | | | 1 | 1 |
| **B.1.160.16** | | | | 1 | | 1 | 2 |
| **B.1.160.19** | | | | | | 1 | 1 |
| **B.1.160.20** | | | | | | 2 | 2 |
| **B.1.160.30** | | | | | | 1 | 1 |
| **B.1.177** | | | 2 | | 2 | 34 | 38 |
| **B.1.177.43** | | | | | | 1 | 1 |
| **B.1.177.44** | | | | | | 1 | 1 |
| **B.1.214** | | | 1 | | | 4 | 5 |

| | | | | | | | |
|---|---|---|---|---|---|---|---|
| **B.1.214.2** | | 1 | | | 2 | 6 | 9 |
| **B.1.221** | | | | | 1 | 17 | 18 |
| **B.1.236** | | | | | | 1 | 1 |
| **B.1.258** | | | | | | 11 | 11 |
| **B.1.258.14** | | | | | | 1 | 1 |
| **B.1.258.17** | | | | | | 3 | 3 |
| **B.1.351** | | | | | | 2 | 2 |
| **B.1.36** | | | | | 1 | 3 | 4 |
| **B.1.36.1** | | | | | | 3 | 3 |
| **B.1.36.35** | | | | | | 3 | 3 |
| **B.1.8** | | | | | | 1 | 1 |
| **C.16** | | | | | | 2 | 2 |
| **C.4** | | | | | | 1 | 1 |
| **P.2** | | | 1 | | | 1 | 2 |
| **Total** | 5 | 3 | 17 | 10 | 11 | 249 | 295 |

**Table S10**. Summary of SARS-CoV-2 lineages and cases per lineage detected in different educational institutions from 01.03.2021 to 31.05.2021.

| | Daycare | Kindergarten | Elementary | secondary_I | secondary_II | adults | Total |
|---|---|---|---|---|---|---|---|
| **B.1** | | | | | | 3 | 3 |
| **B.1.1** | | | | | | 4 | 4 |
| **B.1.1.318** | | | | | 1 | | 1 |
| **B.1.1.39** | | | | | | 1 | 1 |
| **B.1.1.519** | | | | | | 2 | 2 |
| **B.1.1.7** | 11 | 8 | 32 | 34 | 43 | 402 | 530 |
| **B.1.1.70** | | | | | | 1 | 1 |
| **B.1.160** | | | | | | 1 | 1 |
| **B.1.177.86** | | | | | | 2 | 2 |
| **B.1.214.2** | 2 | 1 | 2 | 1 | 1 | 22 | 29 |
| **B.1.258** | | | | | | 1 | 1 |
| **B.1.258.17** | | | | 1 | 1 | 6 | 8 |
| **B.1.351** | | 2 | | | | 2 | 4 |
| **B.1.36.35** | | | | | | 3 | 3 |
| **B.1.525** | | | | | | 1 | 1 |

| | | | | | | | |
|---|---|---|---|---|---|---|---|
| **B.1.617.2** | | | 3 | 1 | | 9 | 13 |
| **B.1.620** | | | | | | 1 | 1 |
| **P.1** | | | | | | 4 | 4 |
| **P.1.2** | | | | | | 2 | 2 |
| **Total** | 13 | 11 | 37 | 37 | 46 | 467 | 611 |
